# Gut-Brain Nexus: Mapping Multi-Modal Links to Neurodegeneration at Biobank Scale

**DOI:** 10.1101/2024.09.12.24313490

**Authors:** Mohammad Shafieinouri, Samantha Hong, Artur Schuh, Mary B. Makarious, Rodrigo Sandon, Paul Suhwan Lee, Emily Simmonds, Hirotaka Iwaki, Gracelyn Hill, Cornelis Blauwendraat, Valentina Escott-Price, Yue A. Qi, Alastair J. Noyce, Armando Reyes-Palomares, Hampton L. Leonard, Malu Tansey, Andrew Singleton, Mike A. Nalls, Kristin S Levine, Sara Bandres-Ciga

## Abstract

Alzheimer’s disease (AD) and Parkinson’s disease (PD) are influenced by genetic and environmental factors. Using data from UK Biobank, SAIL Biobank, and FinnGen, we conducted an unbiased, population-scale study to: 1) Investigate how 155 endocrine, nutritional, metabolic, and digestive system disorders are associated with AD and PD risk prior to their diagnosis, considering known genetic influences; 2) Assess plasma biomarkers’ specificity for AD or PD in individuals with these conditions; 3) Develop a multi-modal classification model integrating genetics, proteomics, and clinical data relevant to conditions affecting the gut-brain axis. Our findings show that certain disorders elevate AD and PD risk before AD and PD diagnosis including: insulin and non-insulin dependent diabetes mellitus, noninfective gastro-enteritis and colitis, functional intestinal disorders, and bacterial intestinal infections, among others. Polygenic risk scores revealed lower genetic predisposition to AD and PD in individuals with co-occurring disorders in the study categories, underscoring the importance of regulating the gut-brain axis to potentially prevent or delay the onset of neurodegenerative diseases. The proteomic profile of AD/PD cases was influenced by comorbid endocrine, nutritional, metabolic, and digestive systems conditions. Importantly, we developed multi-modal prediction models integrating clinical, genetic, proteomic and demographic data, the combination of which performs better than any single paradigm approach in disease classification. This work aims to illuminate the intricate interplay between various physiological factors involved in the gut-brain axis and the development of AD and PD, providing a multifactorial systemic understanding that goes beyond traditional approaches. Further, we have developed an interactive resource for the scientific community [https://gut-brain-nexus.streamlit.app/] where researchers can investigate components of the predictive model and can investigate feature effects on a sample level.

**Teaser:** Co-occurring disorders of the gut-brain axis combined with genetic and proteomic data can better predict neurodegenerative risk.

## Introduction

Alzheimer’s disease (AD) and Parkinson’s disease (PD) are the two most common neurodegenerative disorders (1,2) and cumulatively affect over 400 million individuals worldwide (3,4). Though significant genetic risk factors for AD and PD have been identified, sporadic and late-onset forms are thought to be caused by a complex interplay between genetic (5,6) and environmental (7,8) factors. This interplay underscores the imperative to explore a multitude of variables across bodily systems to comprehend their contributions to the etiology of AD/PD (9,10).

Increasingly, research in neurodegeneration emphasizes the role of gut-brain axis health in neurodegeneration risk (11,12). The gut-brain axis is a complex communication network that links the gastrointestinal tract and the central nervous system. This bidirectional system, including neural pathways, hormonal signaling, and immune mechanisms, facilitates constant interactions between the brain, digestive, endocrine, metabolic systems and nutritional status. Conditions that impact the gut-brain axis include digestive system disorders (13), endocrine pathway disorders (14,15), nutritional deficiencies (16,17), and metabolic traits (18).

Endocrine disorders, such as thyroid hormone imbalances, have been linked to AD and PD (15,19), with conditions like hypothyroidism and subclinical hyperthyroidism being associated with dementia risk (20) and both hypo-and hyperthyroidism being associated with increasing PD risk (15). Metabolic disorders, particularly diabetes, are also related to neurodegenerative disease risk. An increased severity of diabetes is associated with a higher risk of PD (21,22) and type 2 diabetes is a recognized risk factor for AD (23).

Consequently, antidiabetic medications are being explored as potential treatments for AD and PD (24). Additionally, nutritional deficiencies such as low vitamin D levels are more prevalent in AD and PD patients (25). Digestive disorders have been observed to precede PD (26) or be significantly associated with an increased risk for dementia (27). These are just a few examples of factors contributing to the gut-brain axis health and their influence on neurodegeneration.

Understanding the connection between disorders of the gut-brain axis and neurodegeneration can provide useful insights into therapeutic interventions, with major implications for prevention and disease prognosis. In this study, we perform a large biobank-scale characterization of the impact that disorders affecting the gut-brain axis and related to endocrine, nutritional, metabolic, and digestive systems have on the risk of AD and PD. Utilizing data from the UK Biobank, SAIL, and FinnGen, we conducted a population-scale, unbiased assessment that aimed to: 1) Investigate the association between 155 diagnoses related to endocrine, nutritional, metabolic, and digestive system disorders and the risk of AD and PD, while also accounting for established genetic factors known to influence the development of these conditions; 2) Evaluate the specificity of plasma biomarkers associated with AD or PD when individuals have these co-occurring conditions; 3) Develop a multi-modal classification model combining all these data modalities; 4) Analyze the interpretability of ML models and deploy an open-access cloud platform to ensure reproducibility and transparency.

## Results

### A prior diagnosis of certain endocrine, nutritional, metabolic, and digestive system related disorders is associated with increased risk for Alzheimer’s disease and Parkinson’s disease

Cox regression models unraveled a total of 16 ICD-10 diagnoses to be significantly associated with the risk of AD (table S1 and fig. S1) in the UKB after correction for multiple comparisons (discovery cohort) that were also found to be significant in either SAIL, FinnGen biobanks, or both (Table 1). A diagnosis of hemorrhoids and perianal venous thrombosis was found to have a HR<1 for AD in all three datasets. This observation could potentially be due to the fact that a hospitalized diagnosis of hemorrhoids and perianal venous thrombosis could be an indication of other, more serious conditions linked to a high mortality rate, thus explaining the protective observed effect (28). The 15 other ICD-10 codes that were found to be significant have a HR>1, suggesting that being diagnosed with these conditions increases the risk for AD; these include amyloidosis; diseases of pulp and periapical tissues; disorders of lipoprotein metabolism and other lipidemias; disorders of mineral metabolism; gastritis and duodenitis; insulin-dependent, non-insulin-dependent, and unspecified diabetes mellitus; oesophagitis; other bacterial intestinal infections; other disorders of fluid, electrolyte and acid-base balance; other functional intestinal disorders; other noninfective gastro-enteritis and colitis; and volume depletion.

**Table 1.**
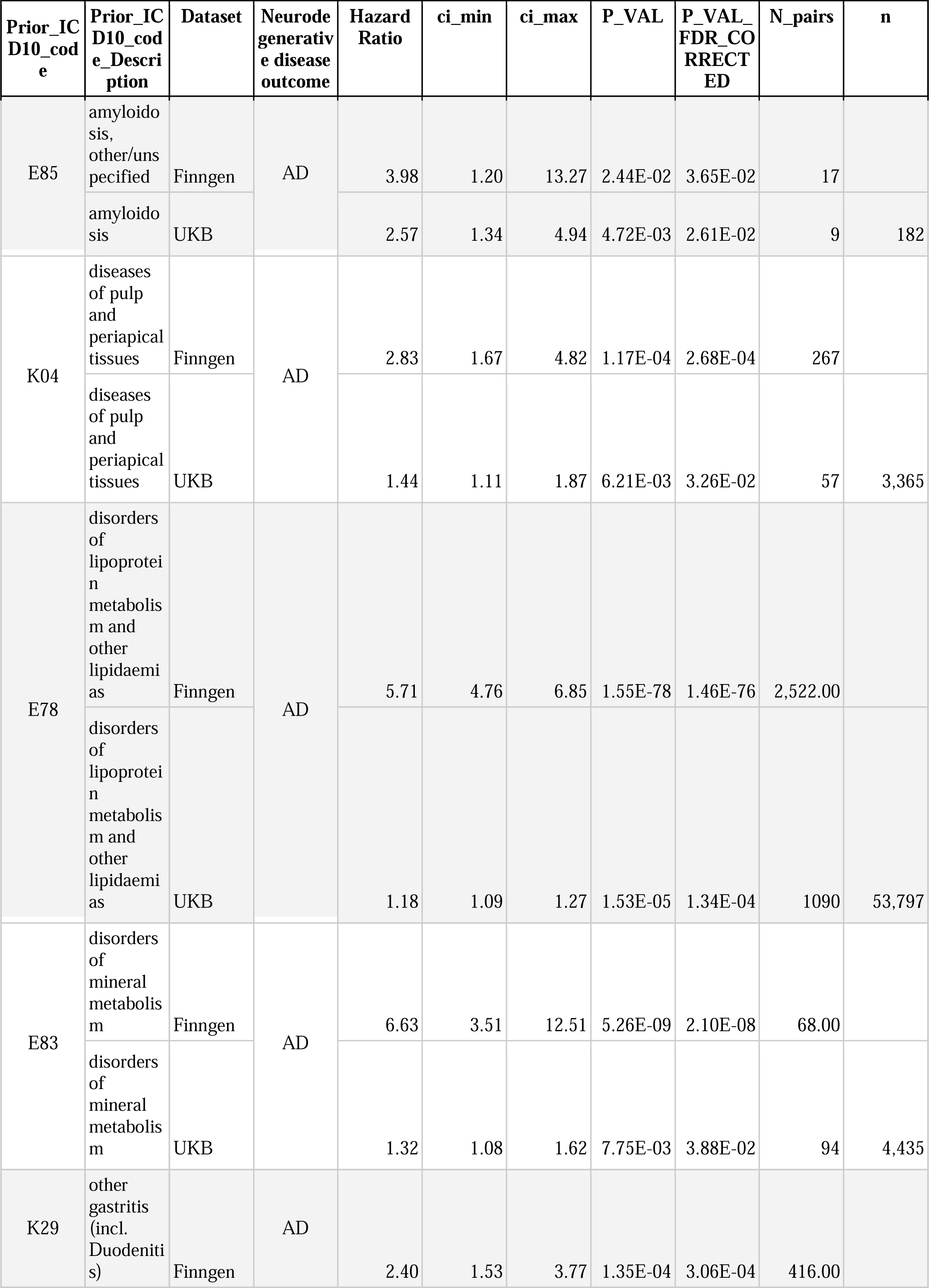
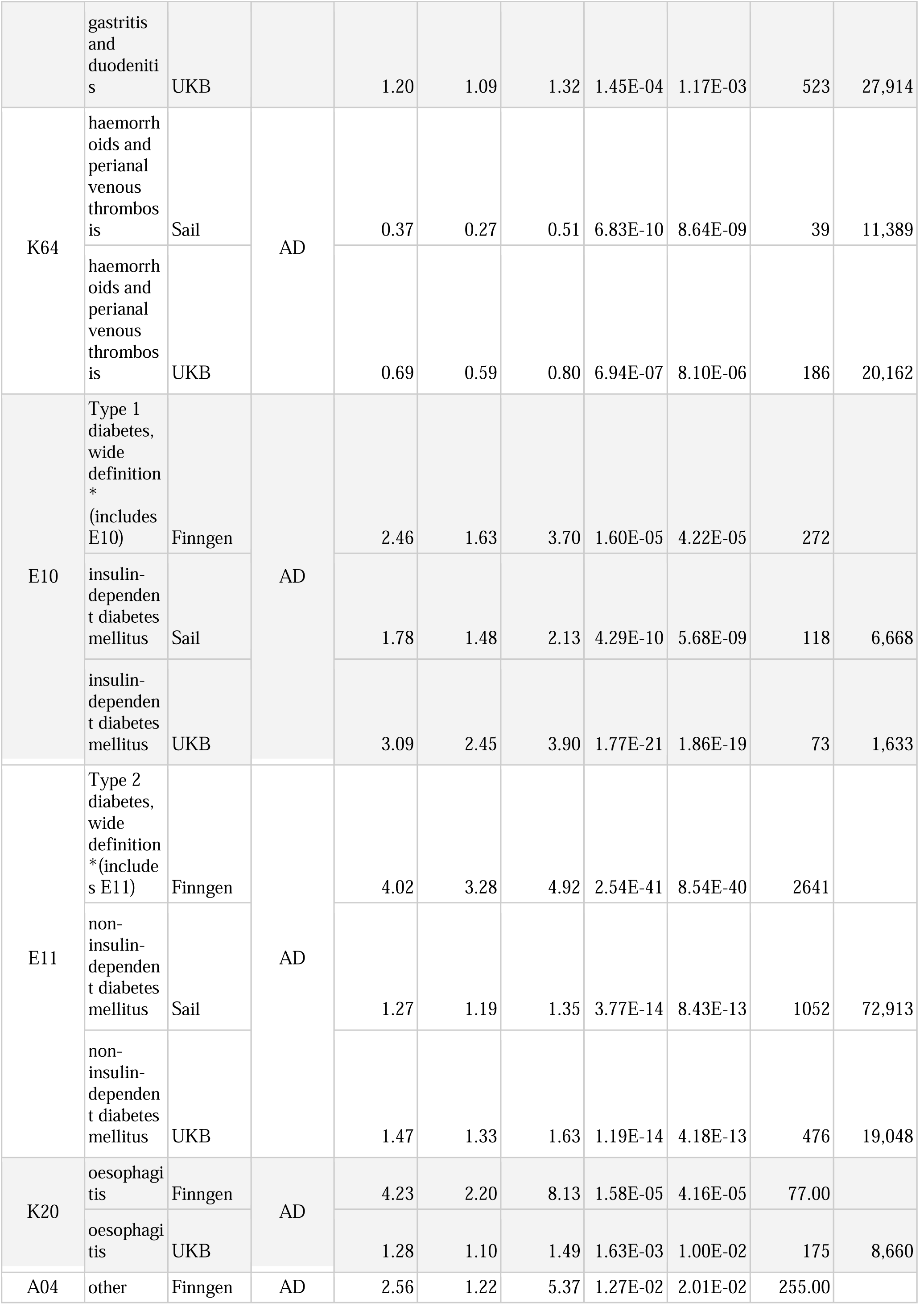
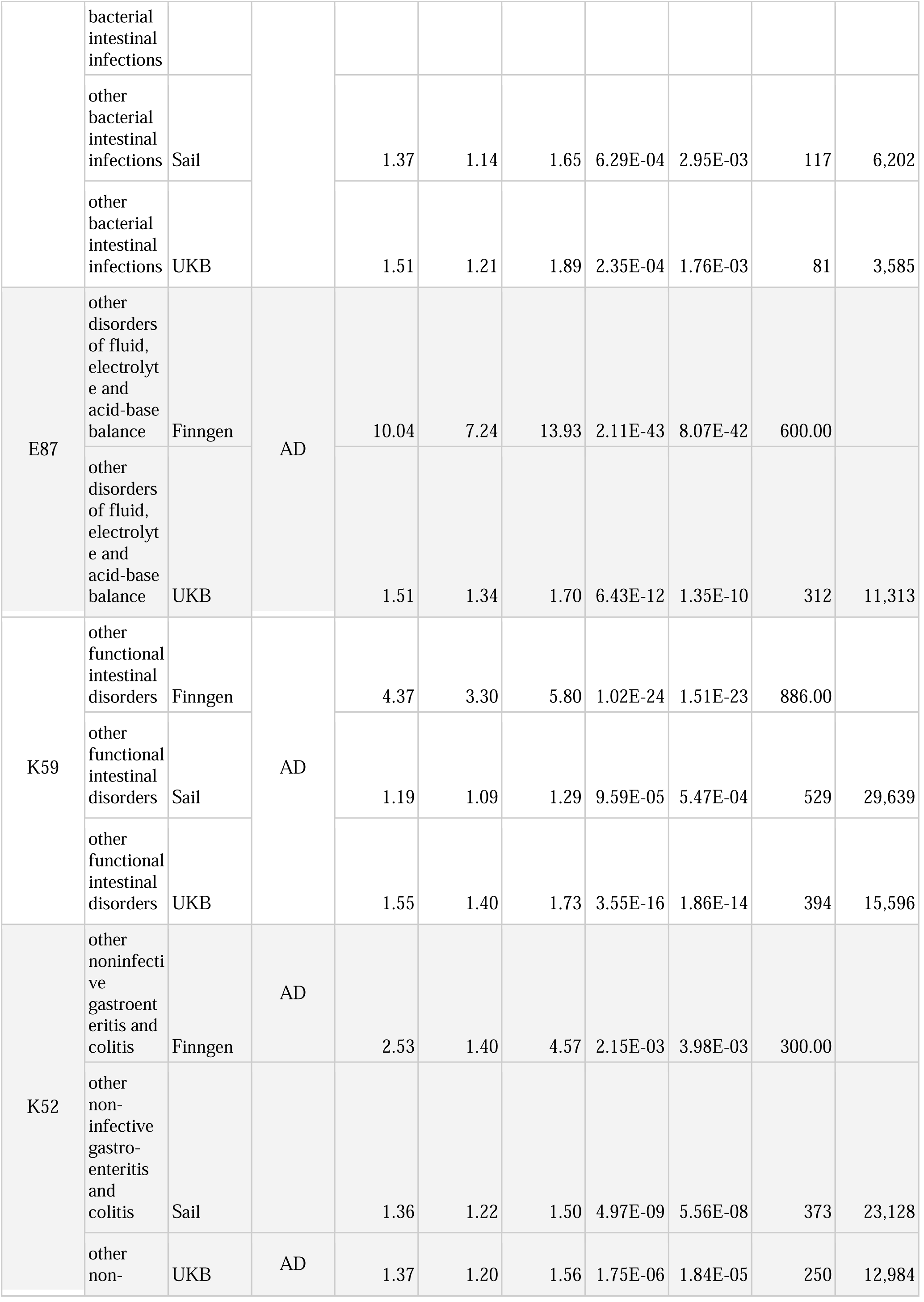
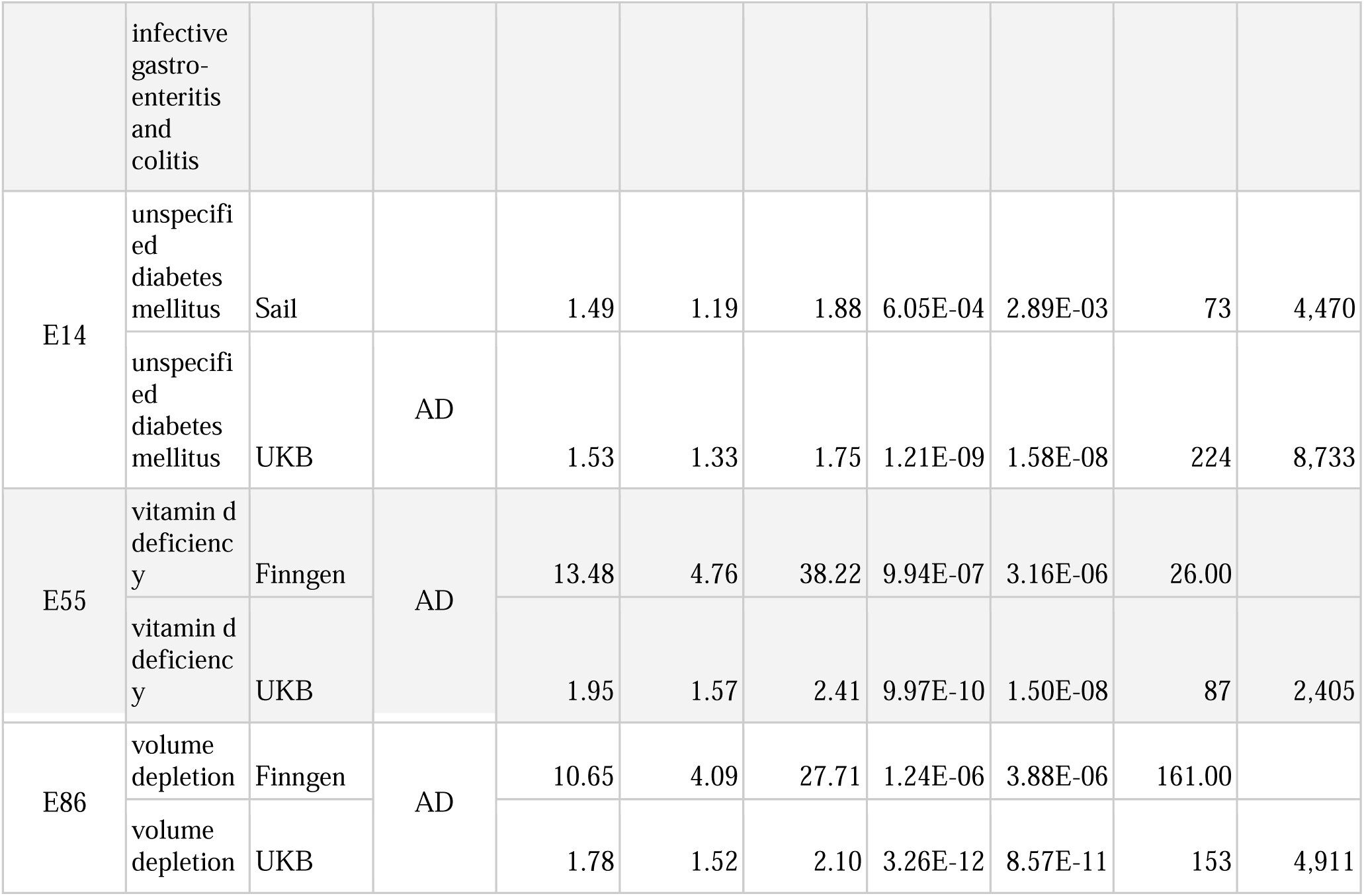
Replicated diagnoses in endocrine, nutritional, metabolic, and digestive systems associated with Alzheimer’s Disease (AD) risk. UKB: UK Biobank. SAIL: Secure Anonymised Information Linkage Databank. Prior_ICD10_code: initial diagnosis of endocrine, metabolic, digestive system, and nutritional disorders. ci_min: Confidence interval minimum. ci_max: Confidence interval maximum. P_VAL: p-value. P_VAL_FDR_CORRECTED: p-value after False Discovery Rate corrected. N_pairs: Number of individuals identified with both ICD-10 code and neurodegenerative disease outcome. n: Number of individuals identified with ICD-10 code.

Of note, our analyses indicate significant associations between 7 disorders and the risk for PD (table S2 and fig. S2) that were replicated in either SAIL or FinnGen biobanks (Table 2). For PD, diverticular disease of the intestine, other diseases of the intestine, and other disorders of the peritoneum have HR<1 replicated in two datasets. Similarly to AD, individuals diagnosed with these conditions are not representative of the entire population due to the severity of these conditions and differential survival rates (29–32). The other 7 ICD-10 codes that had a HR>1 include dyspepsia; insulin-dependent and non-insulin-dependent diabetes mellitus; and other functional intestinal disorders.

**Table 2.** Replicated diagnoses in endocrine, nutritional, metabolic, and digestive systems associated with Parkinson’s Disease (PD) risk. UKB: UK Biobank. SAIL: Secure Anonymised Information Linkage Databank. Prior_ICD10_code: initial diagnosis of endocrine, metabolic, digestive system, and nutritional disorders. ci_min: Confidence interval minimum. ci_max: Confidence interval maximum. P_VAL: p-value. P_VAL_FDR_CORRECTED: p-value after False Discovery Rate corrected. N_pairs: Number of individuals identified with both ICD-10 code and neurodegenerative disease outcome. n: Number of individuals identified with ICD10_code.

| Prior_I<br>CD10_c<br>ode | Prior_I<br>CD10_c<br>ode_Des<br>cription | Dataset | Neurode<br>generati<br>ve<br>disease<br>outcome | Hazard<br>Ratio | ci_min | ci_max | P_VAL | P_VAL_<br>FDR_C<br>ORREC<br>TED | N_pairs | n |
| --- | --- | --- | --- | --- | --- | --- | --- | --- | --- | --- |
| K57 | diverticu<br>lar<br>disease<br>of<br>intestine | Sail | PD | 0.75 | 0.69 | 0.81 | 1.74E-11 | 2.82E-10 | 555.00 | 70,198.0<br>0 |
|  | diverticu<br>lar<br>disease<br>of<br>intestine | UKB |  | 0.69 | 0.62 | 0.77 | 6.24E-11 | 2.04E-09 | 354.00 | 34,801.0<br>0 |
| K30 | functiona<br>l<br>dyspepsi<br>a | Finngen | PD | 2.72 | 1.96 | 3.76 | 1.95E-09 | 1.21E-08 | 86.00 |  |
|  | dyspepsi<br>a | UKB |  | 1.34 | 1.13 | 1.60 | 8.51E-04 | 9.26E-03 | 135.00 | 8,811.00 |
| E10 | insulin-<br>depende<br>nt<br>diabetes<br>mellitus | Sail | PD | 1.59 | 1.31 | 1.93 | 3.36E-06 | 2.48E-05 | 101.00 | 6,668.00 |
|  | insulin-<br>depende<br>nt<br>diabetes<br>mellitus | UKB |  | 2.65 | 2.02 | 3.48 | 2.44E-12 | 1.20E-10 | 53.00 | 1,608.00 |
| E11 | Type 2<br>diabetes,<br>wide<br>definitio<br>n*(Inclu<br>des E11) | Finngen | PD | 2.04 | 1.75 | 2.39 | 2.06E-19 | 3.38E-18 | 421.00 |  |
|  | non-insulin-<br>depende<br>nt<br>diabetes<br>mellitus | UKB |  | 1.21 | 1.08 | 1.36 | 1.36E-03 | 1.33E-02 | 330.00 | 18,815.0<br>0 |
| K63 | other<br>diseases<br>of | Sail | PD | 0.84 | 0.73 | 0.97 | 1.51E-02 | 4.71E-02 | 205.00 | 25,877.0<br>0 |
|  | intestine |  |  |  |  |  |  |  |  |  |
|  | other diseases of intestine | UKB |  | 0.69 | 0.59 | 0.80 | 1.34E-06 | 2.20E-05 | 183.00 | 19,139.00 |
| K66 | other disorders of peritoneum | Sail | PD | 0.70 | 0.53 | 0.93 | 1.32E-02 | 4.21E-02 | 48.00 | 7,406.00 |
|  | other disorders of peritoneum | UKB |  | 0.63 | 0.45 | 0.87 | 5.81E-03 | 4.42E-02 | 35.00 | 4,612.00 |
| K59 | other functional intestinal disorders | Finngen | PD | 3.19 | 2.46 | 4.14 | 2.02E-18 | 3.02E-17 | 178.00 |  |
|  | other functional intestinal disorders | UKB |  | 1.56 | 1.38 | 1.76 | 4.81E-13 | 4.71E-11 | 301.00 | 15,431.00 |

Additionally, we conducted a time-stratified cox regression analysis to evaluate whether the timing of diagnosis for the ICD-10 codes under study impacts HR values for AD or PD. We split the samples from UKB into three strata: 1-5 years, 5-10 years, and 10-15 years prior to AD/PD diagnosis. We then re-evaluated the HRs for AD and PD for the significant ICD-10 codes identified in the previous analysis (Table 3 and Table 4). The values and directions of the HRs in the stratified analysis show that the risk associated with each ICD-10 code is consistent across the three strata.

**Table 3.** Time-Stratified Cox Regression model for Alzheimer’s Disease. ICD10_code: initial diagnosis of endocrine, metabolic, digestive system, and nutritional disorders. ICD10_diagnosis_range: Number of years from the initial diagnosis of endocrine, metabolic, digestive system, and nutritional disorders (ICD-10 code) to the occurrence of neurodegenerative disease outcome. ci_min: Confidence interval minimum. ci_max: Confidence interval maximum. P_VAL: p-value. N_pairs: Number of individuals identified with both ICD-10 code and neurodegenerative disease outcome. n: Number of individuals identified with ICD10_code. P_VAL_FDR_CORRECTED: p-value after False Discovery Rate corrected.

| ICD10_code | ICD10_code_description | ICD10_diagnosis_range | Neurodegenerative disease outcome | Hazard Ratio | ci_min | ci_max | P_VAL | N_pairs | n | P_VAL_FDR_CORRECTED |
| --- | --- | --- | --- | --- | --- | --- | --- | --- | --- | --- |
| A04 | Other Bacterial Intestinal Infections | 1-5 | AD | 2.12 | 1.28 | 3.53 | 3.63E-03 | 15 | 434 | 6.60E-03 |
| A04 | Other Bacterial Intestinal Infections | All | AD | 1.51 | 1.21 | 1.89 | 2.35E-04 | 81 | 3585 | 3.35E-04 |
| E10 | Insulin-Dependent Diabetes Mellitus | 1-5 | AD | 3.13 | 1.85 | 5.29 | 2.05E-05 | 14 | 325 | 5.11E-05 |
| E10 | Insulin-Dependent Diabetes Mellitus | 5-10 | AD | 3.08 | 1.94 | 4.90 | 1.96E-06 | 18 | 398 | 9.78E-06 |
| E10 | Insulin-Dependent Diabetes Mellitus | 10-15 | AD | 3.68 | 2.32 | 5.85 | 3.55E-08 | 18 | 318 | 1.60E-07 |
| E10 | Insulin-Dependent Diabetes Mellitus | All | AD | 3.09 | 2.45 | 3.90 | 1.77E-21 | 73 | 1633 | 3.55E-20 |
| E11 | Non-Insulin-Dependent Diabetes Mellitus | 5-10 | AD | 1.32 | 1.11 | 1.58 | 1.91E-03 | 128 | 5496 | 4.78E-03 |
| E11 | Non-Insulin-Dependent Diabetes Mellitus | 10-15 | AD | 1.82 | 1.53 | 2.17 | 1.59E-11 | 133 | 4023 | 2.86E-10 |
| E11 | Non-Insulin-Dependent Diabetes Mellitus | All | AD | 1.47 | 1.33 | 1.63 | 1.19E-14 | 476 | 19048 | 7.96E-14 |
| E14 | Unspecifi<br>ed<br>Diabetes<br>Mellitus | 5-10 | AD | 1.79 | 1.13 | 2.85 | 1.36E-02 | 18 | 827 | 2.47E-02 |
| E14 | Unspecifi<br>ed<br>Diabetes<br>Mellitus | 10-15 | AD | 2.28 | 1.76 | 2.94 | 4.16E-10 | 59 | 1803 | 3.74E-09 |
| E14 | Unspecifi<br>ed<br>Diabetes<br>Mellitus | All | AD | 1.53 | 1.33 | 1.75 | 1.21E-09 | 224 | 8733 | 3.02E-09 |
| E16 | Other<br>Disorders<br>Of<br>Pancreati<br>c Internal<br>Secretion | 1-5 | AD | 3.52 | 2.41 | 5.15 | 7.75E-11 | 27 | 374 | 1.55E-09 |
| E16 | Other<br>Disorders<br>Of<br>Pancreati<br>c Internal<br>Secretion | 5-10 | AD | 1.87 | 1.21 | 2.91 | 5.12E-03 | 20 | 525 | 1.02E-02 |
| E16 | Other<br>Disorders<br>Of<br>Pancreati<br>c Internal<br>Secretion | 10-15 | AD | 2.69 | 1.34 | 5.38 | 5.23E-03 | 8 | 180 | 1.18E-02 |
| E16 | Other<br>Disorders<br>Of<br>Pancreati<br>c Internal<br>Secretion | All | AD | 2.23 | 1.75 | 2.83 | 5.27E-11 | 69 | 1637 | 1.76E-10 |
| E53 | Deficienc<br>y Of<br>Other B<br>Group<br>Vitamins | 1-5 | AD | 2.33 | 1.66 | 3.27 | 9.65E-07 | 34 | 700 | 3.86E-06 |
| E53 | Deficienc<br>y Of<br>Other B<br>Group<br>Vitamins | 5-10 | AD | 1.74 | 1.08 | 2.80 | 2.29E-02 | 17 | 509 | 3.81E-02 |
| E53 | Deficienc<br>y Of<br>Other B<br>Group<br>Vitamins | All | AD | 1.78 | 1.40 | 2.27 | 2.61E-06 | 68 | 1939 | 4.75E-06 |
| E55 | Vitamin<br>D<br>Deficienc<br>y | 1-5 | AD | 2.35 | 1.80 | 3.06 | 2.47E-10 | 56 | 1198 | 2.47E-09 |
| E55 | Vitamin | All | AD | 1.95 | 1.57 | 2.41 | 9.97E-10 | 87 | 2405 | 2.85E-09 |
|  | D<br>Deficienc<br>y |  |  |  |  |  |  |  |  |  |
| E66 | Obesity | All | AD | 0.85 | 0.74 | 0.96 | 9.29E-03 | 260 | 22619 | 9.29E-03 |
| E78 | Disorders<br>Of<br>Lipoprot<br>ein<br>Metaboli<br>sm And<br>Other<br>Lipidaem<br>ias | 5-10 | AD | 1.48 | 1.29 | 1.70 | 3.33E-08 | 212 | 8983 | 3.33E-07 |
| E78 | Disorders<br>Of<br>Lipoprot<br>ein<br>Metaboli<br>sm And<br>Other<br>Lipidaem<br>ias | 10-15 | AD | 1.44 | 1.27 | 1.63 | 4.43E-09 | 288 | 12012 | 2.66E-08 |
| E78 | Disorders<br>Of<br>Lipoprot<br>ein<br>Metaboli<br>sm And<br>Other<br>Lipidaem<br>ias | All | AD | 1.18 | 1.09 | 1.27 | 1.53E-05 | 1090 | 53797 | 2.56E-05 |
| E83 | Disorders<br>Of<br>Mineral<br>Metaboli<br>sm | 1-5 | AD | 1.64 | 1.22 | 2.21 | 9.34E-04 | 45 | 1522 | 2.08E-03 |
| E83 | Disorders<br>Of<br>Mineral<br>Metaboli<br>sm | All | AD | 1.32 | 1.08 | 1.62 | 7.75E-03 | 94 | 4435 | 8.16E-03 |
| E85 | Amyloid<br>osis | 1-5 | AD | 3.59 | 1.61 | 8.00 | 1.75E-03 | 6 | 81 | 3.50E-03 |
| E85 | Amyloid<br>osis | All | AD | 2.57 | 1.34 | 4.94 | 4.72E-03 | 9 | 182 | 5.55E-03 |
| E86 | Volume<br>Depletio<br>n | 1-5 | AD | 1.90 | 1.48 | 2.44 | 5.01E-07 | 63 | 1678 | 2.50E-06 |
| E86 | Volume<br>Depletio<br>n | 5-10 | AD | 1.89 | 1.32 | 2.71 | 5.28E-04 | 30 | 795 | 1.56E-03 |
| E86 | Volume<br>Depletio<br>n | 10-15 | AD | 2.09 | 1.28 | 3.41 | 3.30E-03 | 16 | 374 | 9.91E-03 |
| E86 | Volume | All | AD | 1.78 | 1.52 | 2.10 | 3.26E-12 | 153 | 4911 | 1.63E-11 |
|  | Depletion |  |  |  |  |  |  |  |  |  |
| E87 | Other Disorders Of Fluid, Electrolyte And Acid-Base Balance | 1-5 | AD | 1.52 | 1.27 | 1.81 | 3.99E-06 | 128 | 4059 | 1.14E-05 |
| E87 | Other Disorders Of Fluid, Electrolyte And Acid-Base Balance | 5-10 | AD | 1.72 | 1.37 | 2.16 | 3.78E-06 | 75 | 1965 | 1.51E-05 |
| E87 | Other Disorders Of Fluid, Electrolyte And Acid-Base Balance | 10-15 | AD | 1.92 | 1.34 | 2.75 | 3.70E-04 | 30 | 786 | 1.33E-03 |
| E87 | Other Disorders Of Fluid, Electrolyte And Acid-Base Balance | All | AD | 1.51 | 1.34 | 1.70 | 6.43E-12 | 312 | 11313 | 2.57E-11 |
| K04 | Diseases Of Pulp And Periapical Tissues | 1-5 | AD | 2.41 | 1.20 | 4.82 | 1.31E-02 | 8 | 305 | 1.87E-02 |
| K04 | Diseases Of Pulp And Periapical Tissues | All | AD | 1.44 | 1.11 | 1.87 | 6.21E-03 | 57 | 3365 | 6.90E-03 |
| K20 | Oesophagitis | 1-5 | AD | 1.62 | 1.16 | 2.28 | 4.88E-03 | 34 | 1399 | 8.14E-03 |
| K20 | Oesophagitis | 5-10 | AD | 1.51 | 1.15 | 1.99 | 3.18E-03 | 52 | 2290 | 7.06E-03 |
| K20 | Oesophagitis | All | AD | 1.28 | 1.10 | 1.49 | 1.63E-03 | 175 | 8660 | 2.03E-03 |
| K29 | Gastritis And Duodenitis | 5-10 | AD | 1.32 | 1.13 | 1.55 | 5.47E-04 | 160 | 7742 | 1.56E-03 |
| K29 | Gastritis And | 10-15 | AD | 1.31 | 1.09 | 1.57 | 3.89E-03 | 120 | 5837 | 1.00E-02 |
|  | Duodenitis |  |  |  |  |  |  |  |  |  |
| K29 | Gastritis And Duodenitis | All | AD | 1.20 | 1.09 | 1.32 | 1.45E-04 | 523 | 27914 | 2.23E-04 |
| K52 | Other Non-Infective Gastro-Enteritis And Colitis | 5-10 | AD | 2.31 | 1.82 | 2.93 | 6.20E-12 | 69 | 2086 | 1.24E-10 |
| K52 | Other Non-Infective Gastro-Enteritis And Colitis | 10-15 | AD | 1.36 | 1.08 | 1.70 | 7.92E-03 | 77 | 3697 | 1.43E-02 |
| K52 | Other Non-Infective Gastro-Enteritis And Colitis | All | AD | 1.37 | 1.20 | 1.56 | 1.75E-06 | 250 | 12984 | 3.50E-06 |
| K59 | Other Functional Intestinal Disorders | 1-5 | AD | 1.55 | 1.30 | 1.85 | 1.27E-06 | 127 | 4476 | 4.22E-06 |
| K59 | Other Functional Intestinal Disorders | 5-10 | AD | 1.70 | 1.40 | 2.07 | 8.75E-08 | 105 | 3464 | 5.83E-07 |
| K59 | Other Functional Intestinal Disorders | 10-15 | AD | 1.41 | 1.10 | 1.82 | 7.32E-03 | 61 | 2512 | 1.43E-02 |
| K59 | Other Functional Intestinal Disorders | All | AD | 1.55 | 1.40 | 1.73 | 3.55E-16 | 394 | 15596 | 3.55E-15 |
| K64 | Haemorrhoids And Perianal Venous Thrombosis | 1-5 | AD | 0.41 | 0.29 | 0.58 | 2.60E-07 | 34 | 5880 | 1.73E-06 |
| K64 | Haemorrhoids | All | AD | 0.69 | 0.59 | 0.80 | 6.94E-07 | 186 | 20162 | 1.54E-06 |
|  | And Perianal Venous Thrombosis |  |  |  |  |  |  |  |  |  |
| K66 | Other Disorders Of Peritoneum | 1-5 | AD | 0.49 | 0.28 | 0.86 | 1.28E-02 | 12 | 1668 | 1.87E-02 |
| K66 | Other Disorders Of Peritoneum | All | AD | 0.59 | 0.44 | 0.81 | 8.77E-04 | 41 | 4636 | 1.17E-03 |

**Table 4.** Time-Stratified Cox Regression model for Parkinson’s Disease. ICD10_code: initial diagnosis of endocrine, metabolic, digestive system, and nutritional disorders. ICD10_diagnosis_range: Number of years from the initial diagnosis of endocrine, metabolic, digestive system, and nutritional disorders (ICD-10 code) to the occurrence of neurodegenerative disease outcome. ci_min: Confidence interval minimum. ci_max: Confidence interval maximum. P_VAL: p-value. N_pairs: Number of individuals identified with both ICD-10 code and neurodegenerative disease outcome. n: Number of individuals identified with ICD10_code. P_VAL_FDR_CORRECTED: p-value after False Discovery Rate corrected.

| ICD10_c<br>ode | ICD10_c<br>ode_desc<br>ription | ICD10_d<br>iagnosis_<br>range | Neurode<br>generati<br>ve<br>disease<br>outcome | Hazard<br>Ratio | ci_min | ci_max | P_VAL | N_pairs | n | P_VAL_<br>FDR_C<br>ORREC<br>TED |
| --- | --- | --- | --- | --- | --- | --- | --- | --- | --- | --- |
| E10 | insulin-<br>dependen<br>t diabetes<br>mellitus | 5-10 | PD | 3.08 | 1.89 | 5.04 | 7.15E-06 | 16 | 396 | 4.29E-05 |
| E10 | insulin-<br>dependen<br>t diabetes<br>mellitus | 10-15 | PD | 2.53 | 1.36 | 4.71 | 3.39E-03 | 10 | 310 | 1.02E-02 |
| E10 | insulin-<br>dependen<br>t diabetes<br>mellitus | All | PD | 2.65 | 2.02 | 3.48 | 2.44E-12 | 53 | 1608 | 1.46E-11 |
| E11 | non-insulin-<br>dependen<br>t diabetes<br>mellitus | 5-10 | PD | 1.41 | 1.16 | 1.70 | 4.19E-04 | 112 | 5480 | 8.38E-04 |
| E11 | non-insulin-<br>dependen<br>t diabetes<br>mellitus | 10-15 | PD | 1.40 | 1.13 | 1.74 | 2.03E-03 | 86 | 3976 | 8.10E-03 |
| E11 | non-insulin-<br>dependen<br>t diabetes<br>mellitus | All | PD | 1.21 | 1.08 | 1.36 | 1.36E-03 | 330 | 18815 | 1.63E-03 |
| E14 | unspecifi<br>ed<br>diabetes<br>mellitus | 1-5 | PD | 1.94 | 1.17 | 3.22 | 1.06E-02 | 15 | 623 | 1.58E-02 |
| E14 | unspecifi<br>ed<br>diabetes<br>mellitus | 5-10 | PD | 2.66 | 1.76 | 4.01 | 3.04E-06 | 23 | 832 | 3.65E-05 |
| E14 | unspecifi<br>ed<br>diabetes<br>mellitus | 10-15 | PD | 1.94 | 1.45 | 2.60 | 8.17E-06 | 46 | 1790 | 9.80E-05 |
| E14 | unspecifi<br>ed<br>diabetes<br>mellitus | All | PD | 1.61 | 1.39 | 1.86 | 1.81E-10 | 197 | 8658 | 5.44E-10 |
| E16 | other<br>disorders<br>of<br>pancreati<br>c internal secretion | 1-5 | PD | 1.89 | 1.04 | 3.42 | 3.54E-02 | 11 | 358 | 4.73E-02 |
| E16 | other disorders of pancreatic internal secretion | 5-10 | PD | 1.94 | 1.17 | 3.22 | 1.06E-02 | 15 | 520 | 1.81E-02 |
| E16 | other disorders of pancreatic internal secretion | All | PD | 1.84 | 1.36 | 2.47 | 6.76E-05 | 44 | 1602 | 1.16E-04 |
| E53 | deficiency of other b group vitamins | 1-5 | PD | 2.04 | 1.34 | 3.10 | 8.81E-04 | 22 | 688 | 2.64E-03 |
| E53 | deficiency of other b group vitamins | All | PD | 1.72 | 1.30 | 2.29 | 1.88E-04 | 48 | 1907 | 2.81E-04 |
| E66 | obesity | 1-5 | PD | 0.63 | 0.47 | 0.83 | 1.45E-03 | 47 | 6561 | 2.95E-03 |
| E66 | obesity | All | PD | 0.82 | 0.71 | 0.94 | 5.86E-03 | 211 | 22453 | 5.86E-03 |
| K30 | dyspepsia | 5-10 | PD | 1.84 | 1.39 | 2.44 | 1.90E-05 | 50 | 2480 | 7.61E-05 |
| K30 | dyspepsia | All | PD | 1.34 | 1.13 | 1.60 | 8.51E-04 | 135 | 8811 | 1.13E-03 |
| K57 | diverticular disease of intestine | 1-5 | PD | 0.60 | 0.48 | 0.75 | 8.54E-06 | 77 | 10132 | 5.12E-05 |
| K57 | diverticular disease of intestine | 5-10 | PD | 0.68 | 0.56 | 0.83 | 1.19E-04 | 104 | 10867 | 2.85E-04 |
| K57 | diverticular disease of intestine | All | PD | 0.69 | 0.62 | 0.77 | 6.24E-11 | 354 | 34801 | 2.50E-10 |
| K59 | other functional intestinal disorders | 1-5 | PD | 1.51 | 1.23 | 1.86 | 8.27E-05 | 94 | 4443 | 3.31E-04 |
| K59 | other functional intestinal disorders | 5-10 | PD | 1.62 | 1.29 | 2.03 | 3.70E-05 | 76 | 3435 | 1.11E-04 |
| K59 | other functional l<br>intestinal<br>disorders | 10-15 | PD | 1.61 | 1.22 | 2.12 | 7.90E-04 | 51 | 2502 | 4.74E-03 |
| K59 | other<br>functiona<br>l<br>intestinal<br>disorders | All | PD | 1.56 | 1.38 | 1.76 | 4.81E-13 | 301 | 15431 | 5.77E-12 |
| K63 | other<br>diseases<br>of<br>intestine | 1-5 | PD | 0.61 | 0.45 | 0.83 | 1.47E-03 | 42 | 5726 | 2.95E-03 |
| K63 | other<br>diseases<br>of<br>intestine | 5-10 | PD | 0.74 | 0.57 | 0.95 | 1.67E-02 | 62 | 6182 | 2.23E-02 |
| K63 | other<br>diseases<br>of<br>intestine | 10-15 | PD | 0.70 | 0.52 | 0.94 | 1.84E-02 | 44 | 4008 | 4.40E-02 |
| K63 | other<br>diseases<br>of<br>intestine | All | PD | 0.69 | 0.59 | 0.80 | 1.34E-06 | 183 | 19139 | 2.69E-06 |
| K64 | haemorrh<br>oids and<br>perianal<br>venous<br>thrombos<br>is | 1-5 | PD | 0.39 | 0.27 | 0.57 | 1.01E-06 | 27 | 5873 | 1.22E-05 |
| K64 | haemorrh<br>oids and<br>perianal<br>venous<br>thrombos<br>is | 5-10 | PD | 0.66 | 0.47 | 0.92 | 1.55E-02 | 34 | 4640 | 2.23E-02 |
| K64 | haemorrh<br>oids and<br>perianal<br>venous<br>thrombos<br>is | All | PD | 0.59 | 0.50 | 0.71 | 3.62E-09 | 136 | 20006 | 8.69E-09 |
| K66 | other<br>disorders<br>of<br>peritoneu<br>m | 1-5 | PD | 0.34 | 0.16 | 0.72 | 4.79E-03 | 7 | 1663 | 8.21E-03 |
| K66 | other<br>disorders<br>of<br>peritoneu<br>m | All | PD | 0.63 | 0.45 | 0.87 | 5.81E-03 | 35 | 4612 | 5.86E-03 |

### Survival analysis indicates increased Alzheimer’s disease and Parkinson’s disease incidence in individuals with significant diagnosis of endocrine, nutritional, metabolic, and digestive system related disorders

Using UKB data, we conducted survival analyses to explore the probabilities of an AD and PD diagnosis at a certain time interval. We generated Kaplan-Meier plots for the significantly associated ICD-10 codes as depicted in fig. S3 and fig. S4, respectively. At the beginning of the observation period, the survival probability starts at 1.0, indicating that all individuals diagnosed with an ICD-10 code are initially free from AD and PD. Over time, this probability diminishes as more individuals are diagnosed with AD or PD. The curves demonstrate the impact of specific ICD-10 code diagnosis on the likelihood of developing these neurodegenerative diseases. Notably, individuals with the significantly associated ICD-10 code diagnoses exhibited a higher incidence of AD and PD.

### Genetic susceptibility for Alzheimer’s disease and Parkinson’s disease is higher in isolated cases compared to those with co-occurring endocrine, nutritional, metabolic, and digestive disorders

We compared the distribution of PRS in individuals diagnosed only with AD or PD versus those having concurrent AD/PD and any ICD-10 code diagnosis under study for samples from UKB only. Of note, some significant ICD-10 codes associated with AD included those pertaining to diabetes mellitus; other disorders of fluid, electrolyte, and acid-base balance; and obesity. Similarly, there were significant associations between diabetes mellitus, disorders of the peritoneum, and vitamin B group deficiencies, and PD. A comprehensive summary of these results is shown in fig. S5 and fig. S6.

For all significant t-tests, a lower average PRS was observed in individuals with co-occurring AD and another condition compared to individuals with only AD. For instance, in AD patients with a non-insulin-dependent diabetes mellitus diagnosis, our analysis revealed lower average PRS scores compared to individuals with only AD, either including or excluding APOE (t-test = -4.26, P = 2.3e-5 and t-test = -2.88, P = 4.01e-3, respectively). Similar trends were observed in individuals with AD and other bacterial intestinal infections (including APOE t-test = -2.31, P = 2.24e-02; excluding APOE t-test = -2.26, P = 2.54e-02), other disorders of pancreatic internal secretion (including APOE t-test = -2.17, P = 3.12e-02), oesophagitis (including APOE t-test = -3.08, P = 2.33e-02; excluding APOE t-test = -2.39, P = 1.78e-02), as well as gastritis and duodenitis (including APOE t-test = -2.83, P = 4.76e-03).

Of note, lower average PRS was observed for all significant t-tests in individuals with co-occurring PD and another condition compared to individuals with only PD. For instance, PD patients with other disorders of the peritoneum had a lower average PRS score than those with only PD (t-test = -3.00, P = 3.70e-03). Additionally, individuals with PD and other functional intestinal disorders (t-test = -2.08, P = 3.80e-02), insulin-dependent diabetes (t-test = -2.55, P = 1.29e-02), non-insulin dependent diabetes (t-test = -3.68, P = 2.52e-4), or a deficiency of other B group vitamins (t-test = -2.54, P = 1.23e-02) also had lower average PRS scores than individuals with only PD.

This result must be interpreted with caution, as there is potential for collider bias in our analysis. By selecting individuals with AD/PD, we are conditioning on these disorders, which are influenced by both the PRS for AD/PD and other risk factors, such as co-occurring endocrine, nutritional, metabolic, and digestive disorders. This conditioning can induce a spurious association between the PRS for AD/PD and these other risk factors, leading to the appearance of a lower average PRS in the group with AD/PD and co-occurring disorders. Essentially, this bias might make it seem as though lower genetic risk is linked to AD/PD in the presence of ICD-10 diagnoses, when in fact the relationship may be driven by the complex interplay of risk factors.

### The interplay between AD and PD cumulative known genetic effects and the risk for endocrine, nutritional, metabolic, and digestive systems disorders did not display any synergistic impact

We did not identify significant synergistic interaction terms at a nominal P<0.05 and exhibiting an OR>1 in either AD (see table S3 and table S4) or PD (see table S5). This suggests that the combined occurrence of endocrine, nutritional, metabolic, or digestive system related disorders does not lead to a synergic effect on AD/PD risk that surpasses the sum of their individual impacts.

### The proteomic profile of individuals diagnosed with Alzheimer’s disease and Parkinson’s disease is influenced by comorbid endocrine, nutritional, metabolic, and digestive systems conditions

Our analysis showed 22 proteomic biomarkers with notable differences in AD cases versus controls, alongside 156 proteins exhibiting significant distinctions in PD cases compared to controls after controlling for multiple comparisons (table S6 and table S7, respectively). We delved deeper into the levels of proteomic biomarkers, applying a t-test to juxtapose average levels between standalone cases of AD/PD and cases co-occurring with specific ICD-10 codes. After implementing FDR corrections, we found significant differences in proteomic biomarker levels in individuals with only AD/PD compared to individuals with AD/PD and co-occurring ICD-10 conditions. Specifically, we observed that 37 biomarkers displayed increased levels in AD cases with co-occurring ICD-10 diagnosis compared to isolated AD cases (Table 5), while a total of five biomarkers demonstrated significant elevated levels in PD cases with accompanying conditions compared to standalone PD cases (Table 6). These findings hint at distinct associations between specific biomarkers and AD/PD, while others may be influenced by concurrent comorbidities.

**Table 5.**
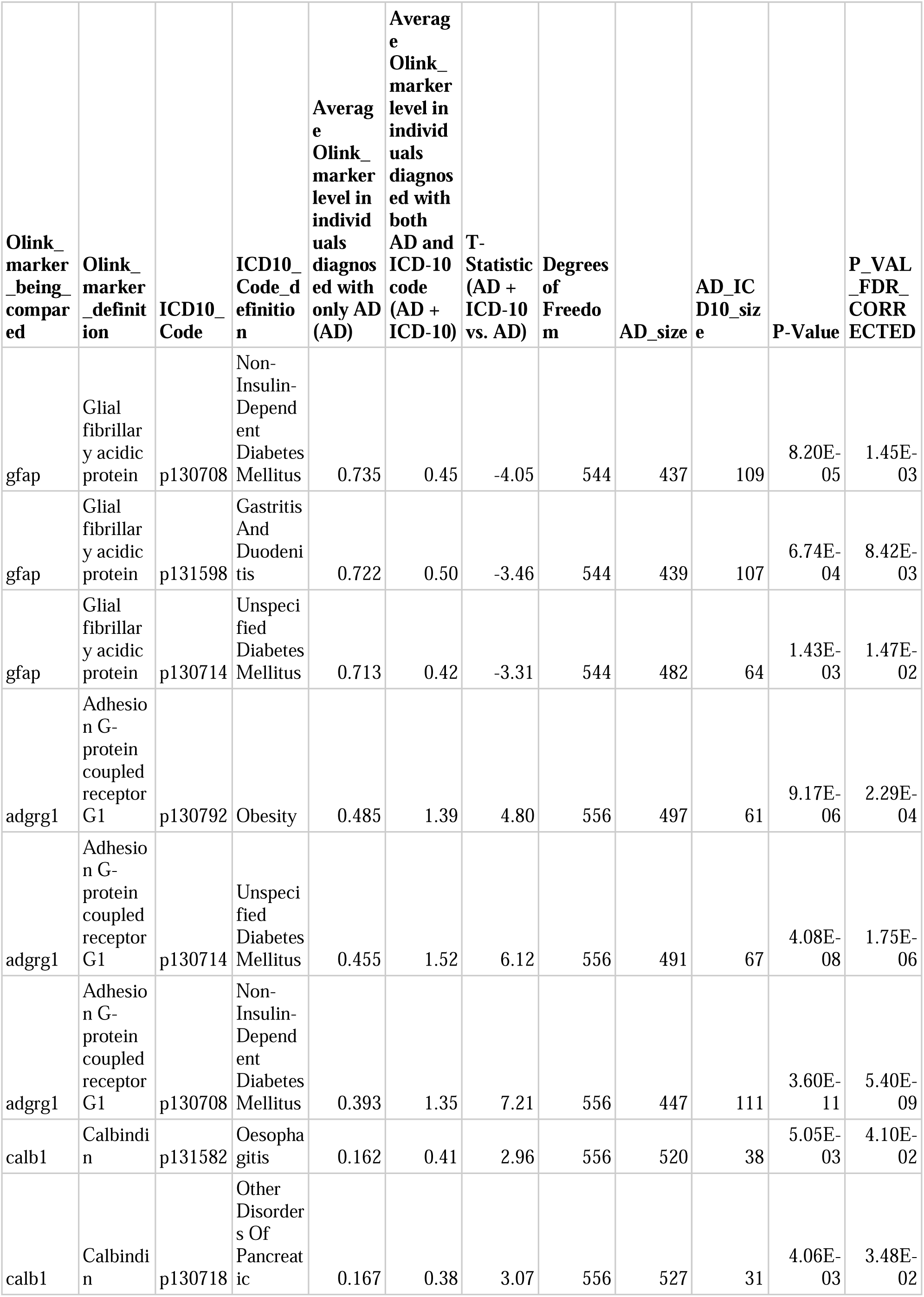
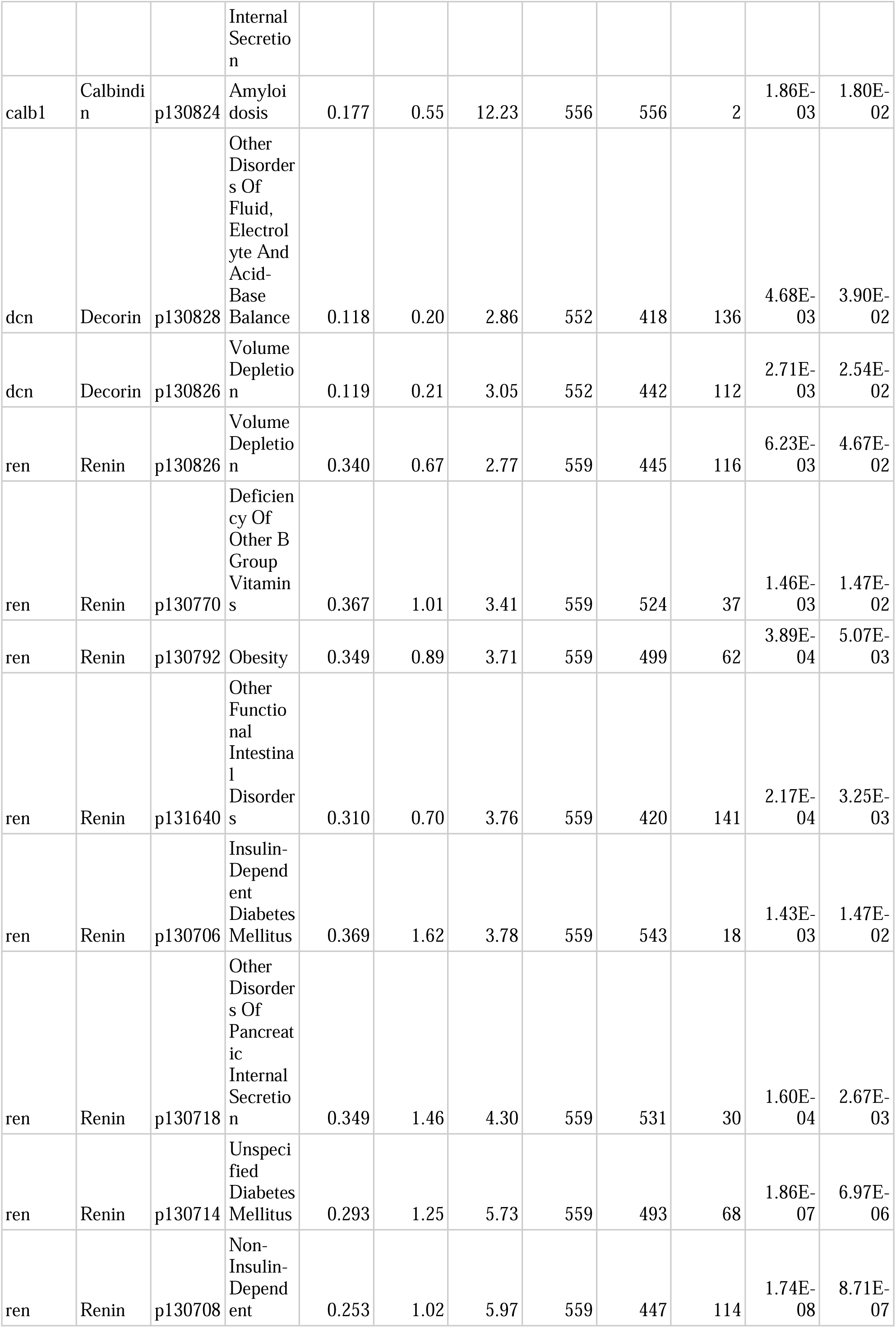
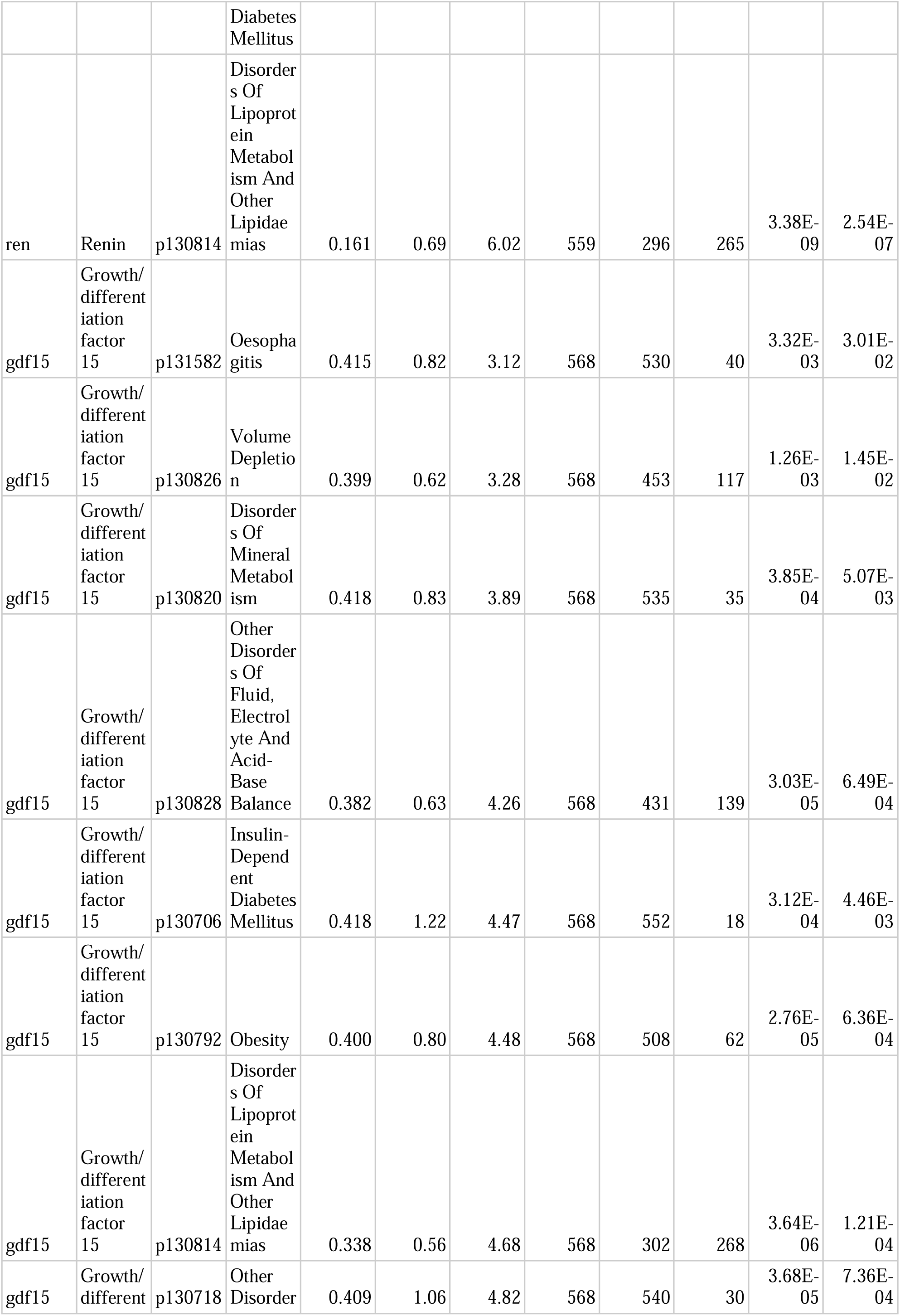
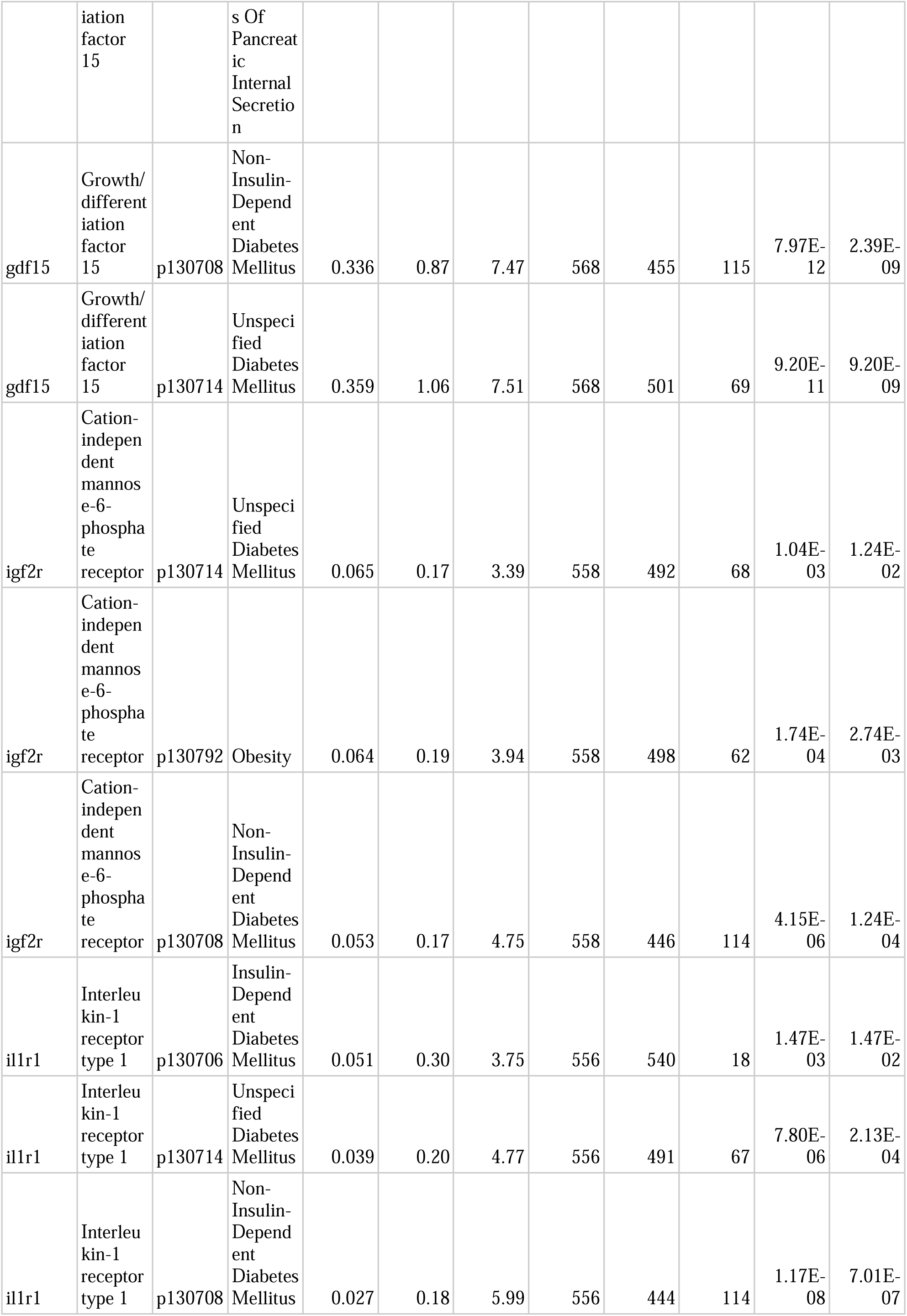
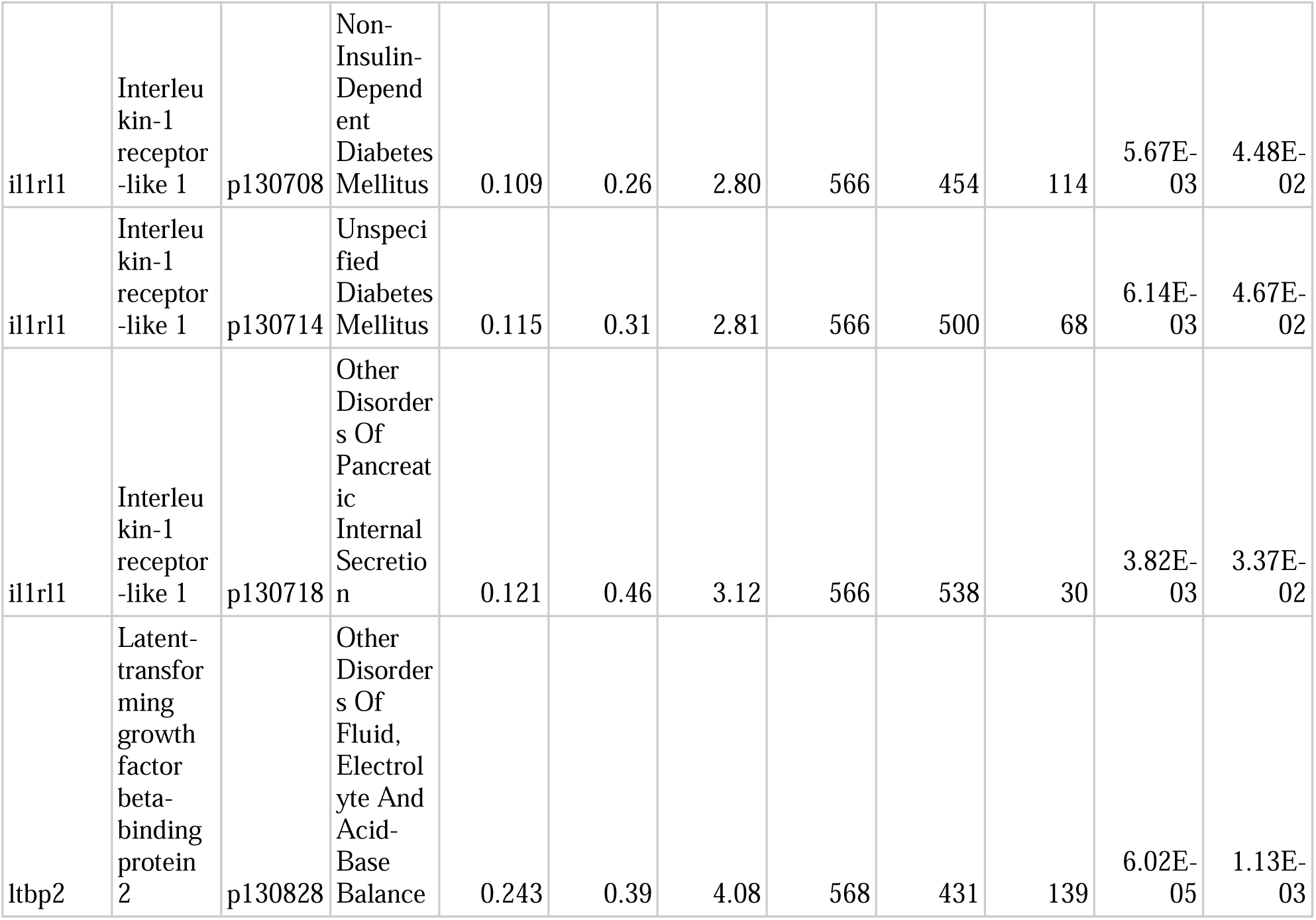
Proteomic biomarker comparison in isolated Alzheimer’s Disease (AD) cases vs. cases with digestive, endocrine, metabolic, and nutritional conditions. P_VAL_FDR_CORRECTED: p-value after False Discovery Rate (FDR) corrected.

**Table 6.**
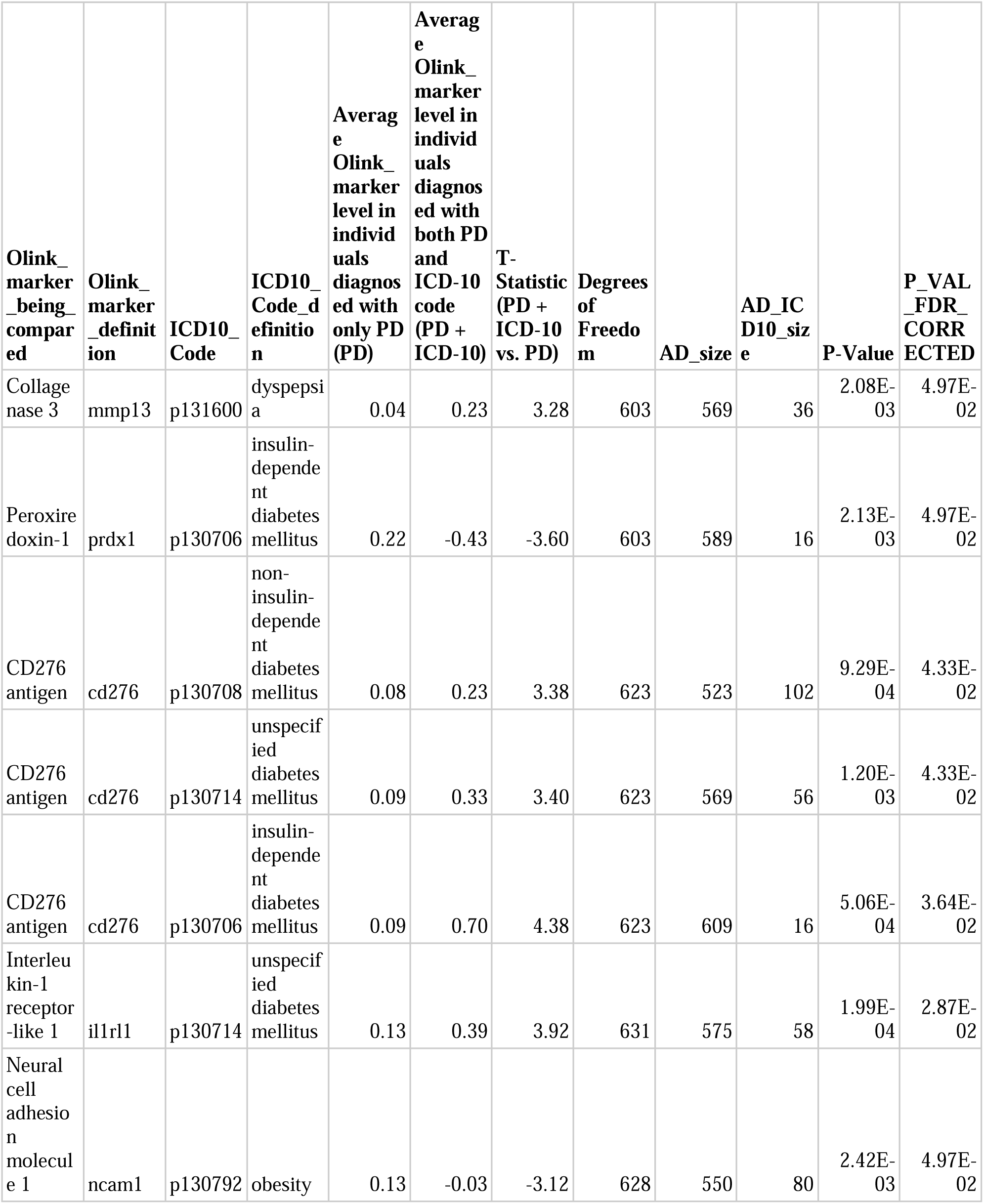
Proteomic biomarker comparison in isolated Parkinson’s Disease (PD) cases vs. cases with digestive, endocrine, metabolic, and nutritional conditions. P_VAL_FDR_CORRECTED: p-value after False Discovery Rate (FDR) corrected.

### Multi omics integration models based on clinical, genetic, and proteomic data improve accuracy to predict Alzheimer’s disease and Parkinson’s disease risk versus a single paradigm

In our analysis, the combination of genetic, clinical, proteomic (controlling for multiple comparisons using Bonferroni correction), and demographic factors (including age at recruitment and gender) exhibited superior predictive performance for AD risk when compared to the individual data sets, with a test AUC of 0.9 [95% CI 0.88-0.92] and test balanced accuracy (BA) of 0.83 [0.80-0.86]. Curiously, the model combining genetic, proteomic, and demographic features (i.e. no clinical information) had a very similar test AUC of 0.89 [0.87-0.91] and a test BA of 0.82 [0.78-0.86], suggesting that the risk information captured by clinical features may be encompassed within the information provided by genetic, proteomic, and demographic features. The best classifier using only one data attribute was the model that used only proteomic data for prediction with a test AUC of 0.87 [0.85-0.89] and test BA of 0.79 [0.76-0.82]. The performance of each predictive model is shown in fig. 1 and the metrics are listed in table S8.

**Fig. 1.**
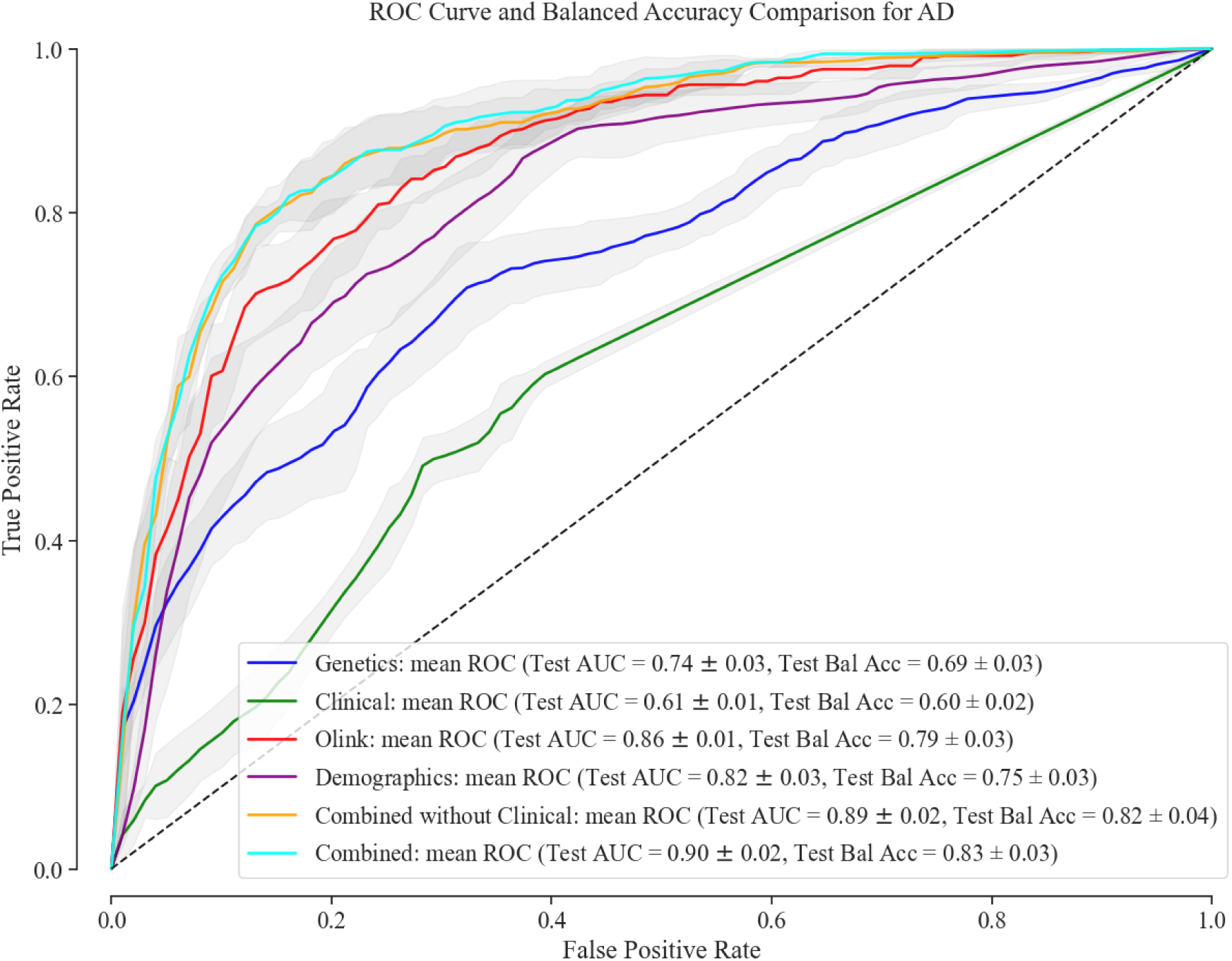
Receiver Operating Characteristic Curve and Balanced Accuracy Comparison for AD. Performance evaluation of multi-omics integration models using clinical, genetic, proteomic, and demographic data for AD.

For PD, the integration of genetic, proteomic and demographic factors showcased heightened predictive efficacy with a test AUC of 0.78 [0.74-0.82] and test BA of 0.70 [0.68-0.72]. For PD, addition of the clinical data did not improve the AUC and test BA of the model. Clinical features do not contain any useful information and predicting solely on clinical features perform no better than a random classifier, with a test AUC of 0.52 [0.49-0.55]. The best classifier using only one data attribute was the model that used demographics data (Age at recruitment, Sex, and Townsend_deprivation_index), with a test AUC of 0.77 [0.74-0.80] and a test BA of 0.72 [0.68-.76]. The performance of each model for PD is shown in fig. 2 and table S9.

**Fig. 2.**
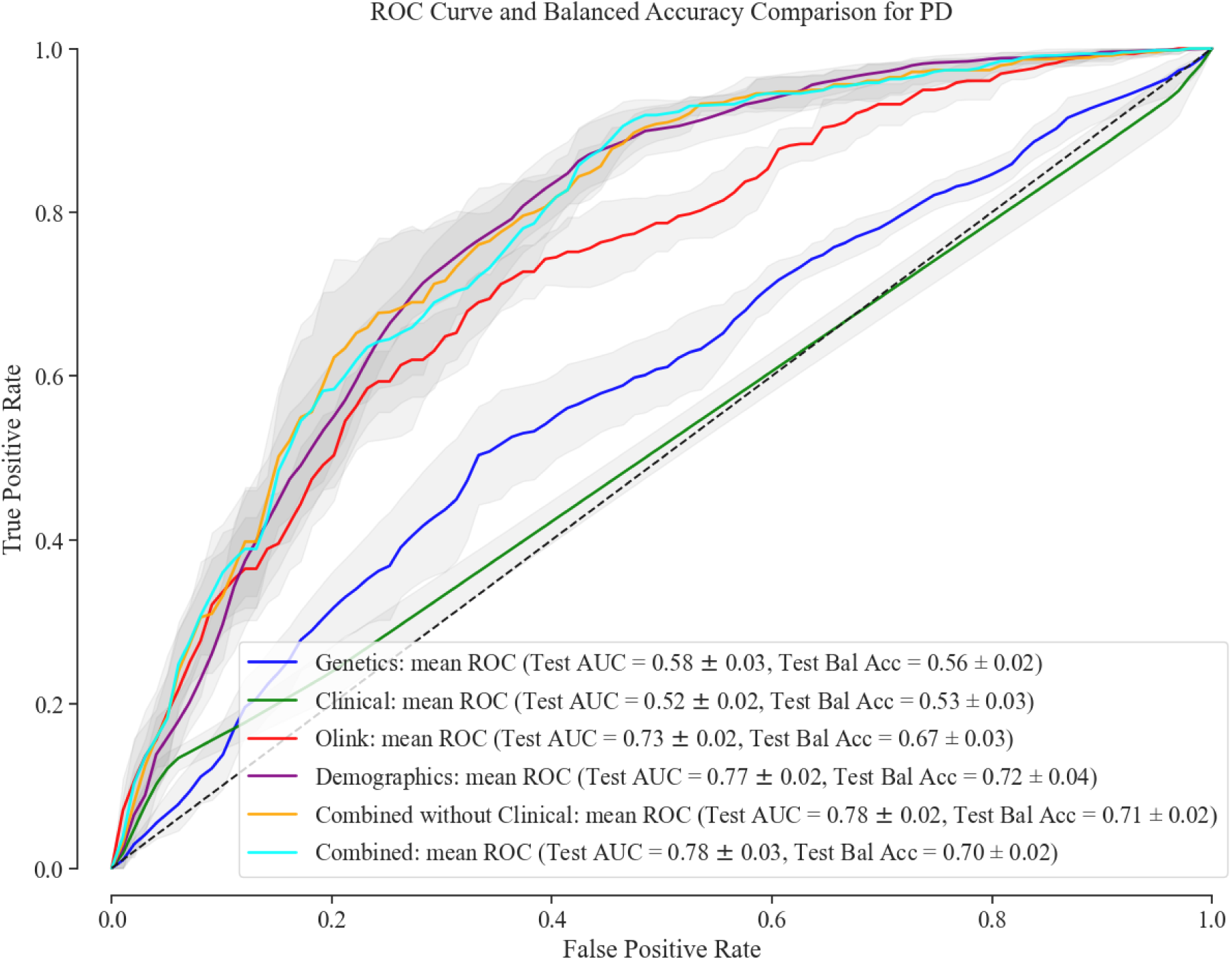
Receiver Operating Characteristic Curve and Balanced Accuracy Comparison for PD. Performance evaluation of multi-omics integration models using clinical, genetic, proteomic, and demographic data for PD.

SHAP values provide an in-depth analysis of machine learning classifiers by highlighting the top discriminating features of AD/PD compared to healthy controls. The biological plausibility of the models is supported by feature importance plots, which emphasize known risk factors for AD and PD, such as age and PRS. Among the proteomic features, neurofilament light polypeptide (NLF) emerges as a significant factor for both AD and PD, suggesting its potential as a biomarker for multiple forms of neurodegeneration. In AD, other top features include glial fibrillary acidic protein (GFAP) and growth differentiation factor (GDF) from proteomics data. From clinical features, only gastritis and duodenitis appear among top features (fig. 3A). Proteomic factors, including Adhesion G-protein coupled receptor G2 and Integrin alpha-M and Interleukin receptor, help distinguish PD from controls (fig. 3B). The interactive website (https://gut-brain-nexus.streamlit.app/) was developed as an open-access and cloud-based platform for researchers to investigate the top features of the machine learning models developed and how these may influence the AD/PD risk scores. Finally, the screenshots of our cloud-based interpretability analysis and multimodal prediction tool are shown in fig. S7-S10.

**Fig. 3.**
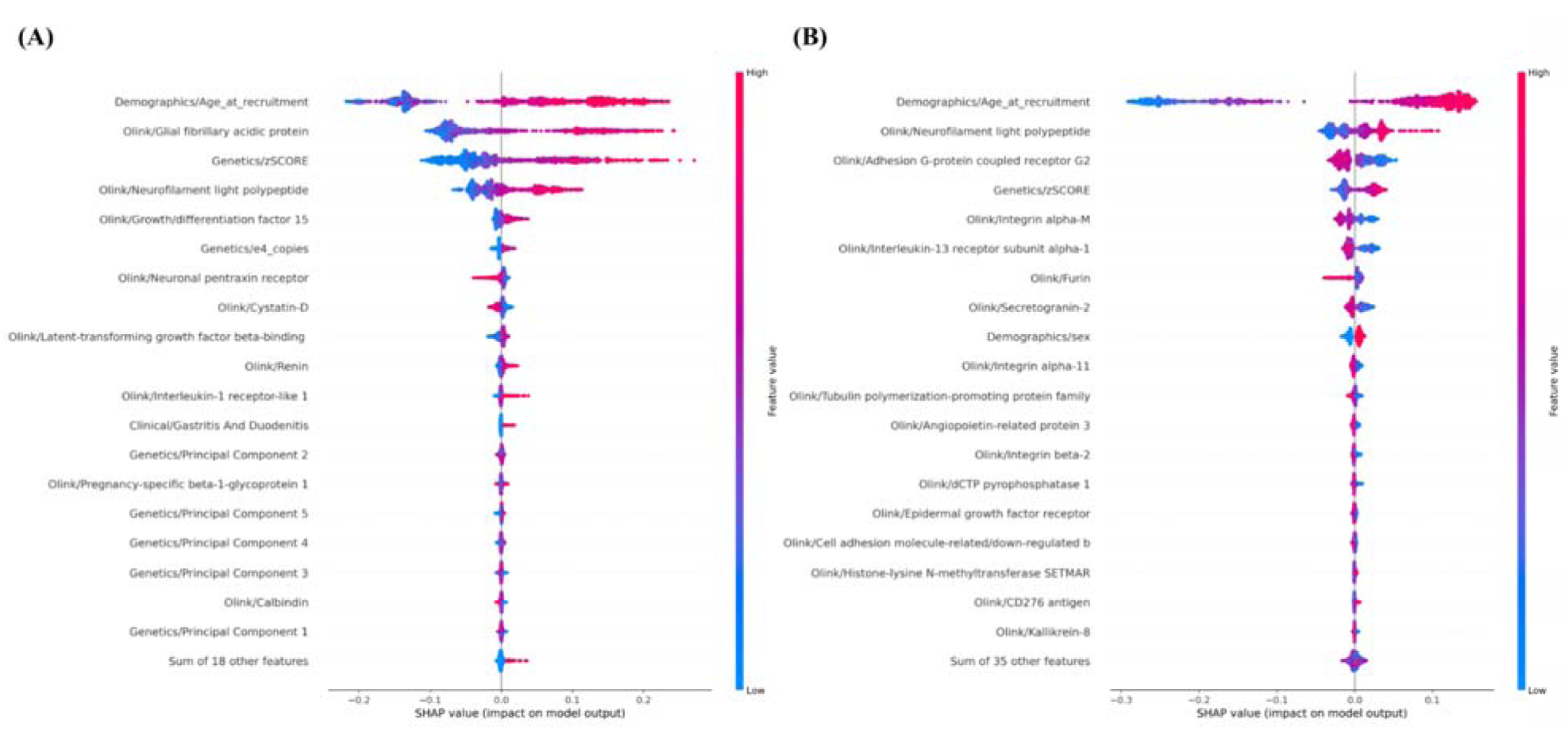
Feature Importance Plots. **(A)** Distribution of the top 20 features that had the most substantial effect on the Alzheimer’s Disease risk estimates. Each point represents a patient and the amount of effect on model output for each feature depends on its Shapley Additive Explanations (SHAP) value. For example, the effect of the “Demographics/Age_at_recruitment” feature on model output is large and positive (indicating a higher risk) when the patient has high values for “Demographics/Age_at_recruitment” (more red points are on the right side). Similarly, **(B)** shows the top features for Parkinson’s Disease risk estimates.

## Discussion

With the increasing prevalence of AD (33) and PD (34), it is imperative to enhance our understanding of the determinants that increase the risk for these common neurodegenerative diseases and most importantly, develop improved prediction models for early detection. Here, we have undertaken the most extensive biobank-scale omics study to date to assess the influence of main biological system disorders implicated in the gut-brain axis (including endocrine, nutritional, metabolic, and digestive-related conditions) preceding the diagnosis of AD and PD. The culmination of which is a multi-modal classification model that combines clinical, genetics, and proteomics data enhancing the prediction accuracy of AD and PD.

In a large-scale and data driven manner, we demonstrate that certain endocrine, nutritional, metabolic, and digestive system related disorders are significantly associated with an increased risk of AD and/or PD prior to diagnosis. Of note, individuals with other non-infective gastroenteritis and colitis; oesophagitis; gastritis and duodenitis; disorders of fluid, electrolyte, and acid-base balance; pancreatic internal secretion disorders; and other functional intestinal disorders showed a higher likelihood of developing AD later in life.

Recent literature has suggested the amplification of AD risk from disorders affecting the gut-brain axis, such as gastritis (35). Our results corroborate these findings and reveal additional, novel potential disorders of interest for further study, with replication across multiple datasets. In regards to PD, significant diagnoses associated with increased PD risk include other functional intestinal disorders, disorders of pancreatic internal secretion, and deficiency of other B group vitamins. The correlation between the deficiency of other B group vitamins and PD expands upon previous studies that have looked at other vitamin deficiencies, such as vitamin D (25), and warrants more research on the impact of nutritional deficiencies on neurodegenerative diseases.

Our study robustly demonstrates that the probability of developing AD/PD increases with certain co-occurring endocrine, metabolic, digestive, or nutritional conditions, corroborating the hypothesis that a diagnosis affecting the gut-brain axis elevates the risk of AD/PD. We also show that risk of neurodegeneration persists up to 15 years before AD/PD onset with a co-occurring diagnosis of an endocrine, metabolic, digestive or nutritional disorder or trait. For example, having a diagnosis of other functional intestinal disorders results in an increased HR for both AD and PD in the periods 1-5, 5-10, and 10-15 years prior to AD/PD diagnosis.

In an effort to investigate genetic distinctions and potential etiological subtypes of AD and PD, we compared polygenic risk for AD and PD in individuals diagnosed only with AD or PD and individuals with AD or PD co-occurring with other endocrine, nutritional, metabolic, and digestive-related disorders. Of note, our study confirmed that individuals diagnosed with any type of diabetes mellitus in addition to AD or PD are shown to have a significant different PRS than individuals with AD or PD alone, in concordance with previous studies showing that diabetes is a risk factor for both AD and PD (36–38). For both AD and PD, comorbidity with any of the significant disorders affecting the gut-brain axis showed lower average PRS scores compared to those with only AD or PD. Our findings suggest that systemic health risk factors can more prominently account for one’s disease risk in the absence of other genetic risk factors in patients with AD or PD, highlighting the importance of considering both genetic as well as other health factors in assessing the overall risk of developing AD and PD. Using diabetes as a positive control in our study, we found that individuals diagnosed with AD along with another disorder affecting the gut-brain axis, such as other bacterial intestinal infections and other functional intestinal disorders, result in a significantly lower average PRS score compared to individuals with only AD. This suggests that severe digestive conditions such as infections affecting the gut microbiome, independent of genetic risk, could increase the risk for AD as previously described (39). For PD, there existed a significant difference in PRS in individuals without vs. with other functional intestinal disorders and other disorders of the peritoneum, in concordance with previous studies suggesting an influence of disorders involving the gut-brain axis on PD (40).

In addition, our analysis revealed that the relationship between the genetic risk for AD/PD and many disorders of the endocrine, nutritional, metabolic and digestive systems resulted in a combined impact not significantly greater than the sum of their individual effects. These interactions are not synergistic, confirming the notion that known genetic risk factors included in the models under study for both AD and PD are independent from gastrointestinal and metabolic disorders, which highlights the importance of environmental factors in the development of both AD and PD. While both genetic and systemic health disorders independently influence the risk of AD and PD, many disorders may not interact in a way that significantly amplifies this risk when combined.

Through our exploration of proteomics and AD/PD risk, we identified several promising candidates for AD/PD diagnosis. For instance, we found that for AD, the proteomic biomarker with the largest impact on disease was glial fibrillary acidic protein (GFAP), supporting previous literature findings that GFAP can serve as an indicator of AD pathology (41). However, the levels of GFAP were significantly decreased in samples with AD and a co-occurring ICD-10 code for gastritis and duodenitis or non-insulin dependent diabetes mellitus. For PD, in concordance with previous studies, we identified peroxiredoxin 1 (PRDX1) as a potential biomarker for disease (42). Again, the levels of PRDX1 were significantly decreased in PD cases and a coinciding ICD-10 code for insulin-dependent diabetes mellitus. These differences in biomarker levels between samples solely with AD/PD vs. samples with additional diagnoses highlight the influence of comorbidities on disease manifestation. Although longitudinal research would need to be conducted, our findings suggest that these biomarkers could serve as valuable diagnostic or prognostic tools for AD and PD, potentially enhancing early detection and disease management.

The inclusion of multiple features, integrating clinical data on digestive, endocrine, nutritional and metabolic disorders, genetic risk scores, and proteomic data in our prediction model, demonstrated superior performance in predicting both AD and PD compared to single-variable paradigms. Co-occurring diagnosis for conditions influencing the gut-brain axis do not seem to influence the predictability of neither AD nor PD as much as the other variables (demographic factors, biomarkers, and genetic status), but the fact that for both AD and PD the combination of data (clinical, genetic, proteomic, and demographic) produces a higher AUC for the ROC curve compared to single modalities underscores the value in including multiple facets of data in predictive models. Overall, these results from our models reveal the promising predictive capabilities of our constructed multi-modal classification models in identifying individuals at risk for AD/PD. The study suggests that the selected proteomic biomarkers, when combined with genetic and other demographic and clinical variables, serve as robust predictors. These predictors have the potential to significantly enhance prediction diagnostic accuracy for AD/PD. In addition, neurofilament light polypeptide proteomic feature stands out as a key factor for both AD and PD, indicating its potential as a biomarker for various forms of neurodegeneration. Our approach highlights the potential clinical utility of multi-omics in enhancing diagnostic accuracy, and further emphasizes the importance of considering other bodily systems when predicting the risk underlying neurodegenerative diseases.

Transparency and reproducibility are critical aspects of science. This becomes even more important for machine learning models due to their dependency on data and the black-box nature of these models. To facilitate this, we made two contributions: (1) Model interpretability analysis, using SHAP values to identify the top features, corresponding to each modality, that can be validated against existing biological findings; (2) Development of an open-access, cloud-based platform for researchers to explore the model developed in this study and investigate how different factors may influence the classification (or, in some cases, misclassification) of a particular sample. Additionally, we incorporated a model perturbation analysis feature on our website, allowing researchers to manually adjust features and observe the resulting changes. These efforts enhance transparency and move the research community away from black-box predictors through interpretable modeling.

Some overall limitations of our work include the use of solely ICD-10 codes for diagnosis, and not other additional assays, which may overlook undiagnosed cases, leading to a potential underestimation of the true impact of these disorders on AD/PD risk. Across datasets, the available diagnosis codes also differ causing further limitations in comparison and validation. Because individuals with AD often present with different cognitive, behavioral, and pathological features, diagnosis of AD is often difficult and inconsistent (43). Though we use the ICD-10 code to filter for AD in this study, it is possible for AD to be interchanged with dementia as diagnosing AD is difficult. The sample makeup across data sources can also be different. Since the UKB participants are volunteers who may have agreed to participate before reaching old age, the incidence of AD/PD may be different compared to the SAIL and FinnGen datasets. The cross-sectional nature of the data used to study biomarker associations may limit any causal inference. The SAIL and FinnGen datasets do not include proteomic data, preventing us from further validating our modeling efforts. The focus on samples of European ancestry constrains the generalizability of our findings to other populations from other genetic backgrounds. Future work needs to be done across different ancestral backgrounds to be globally representative.

Our study delves into the intricate interactions between clinical, genetic and proteomic data, culminating in the construction of a comprehensive multi-modal classification model. This pioneering endeavor aims to shed light on the nuanced interplay between various physiological factors playing a role on the gut-brain axis and the development of AD and PD, offering a multifactorial systemic understanding that transcends traditional approaches. Our integrated approach serves as a proof of concept, aligning with the expanding body of evidence that underscores the intricate etiological foundations of neurodegenerative diseases and holds promise for refining risk prediction models and devising targeted preventive strategies. Further, we have developed an interactive resource for the scientific community [https://gut-brain-nexus.streamlit.app/]. This, in turn, propels our endeavors in elucidating clinical interventions aimed at addressing these debilitating conditions.

## Materials and Methods

This study utilizes data from three biobanks: the UK Biobank (UKB), FinnGen, and the Secure Anonymized Information Linkage (SAIL) (See fig. 4 for an workflow rationale of this study).

**Fig. 4.**
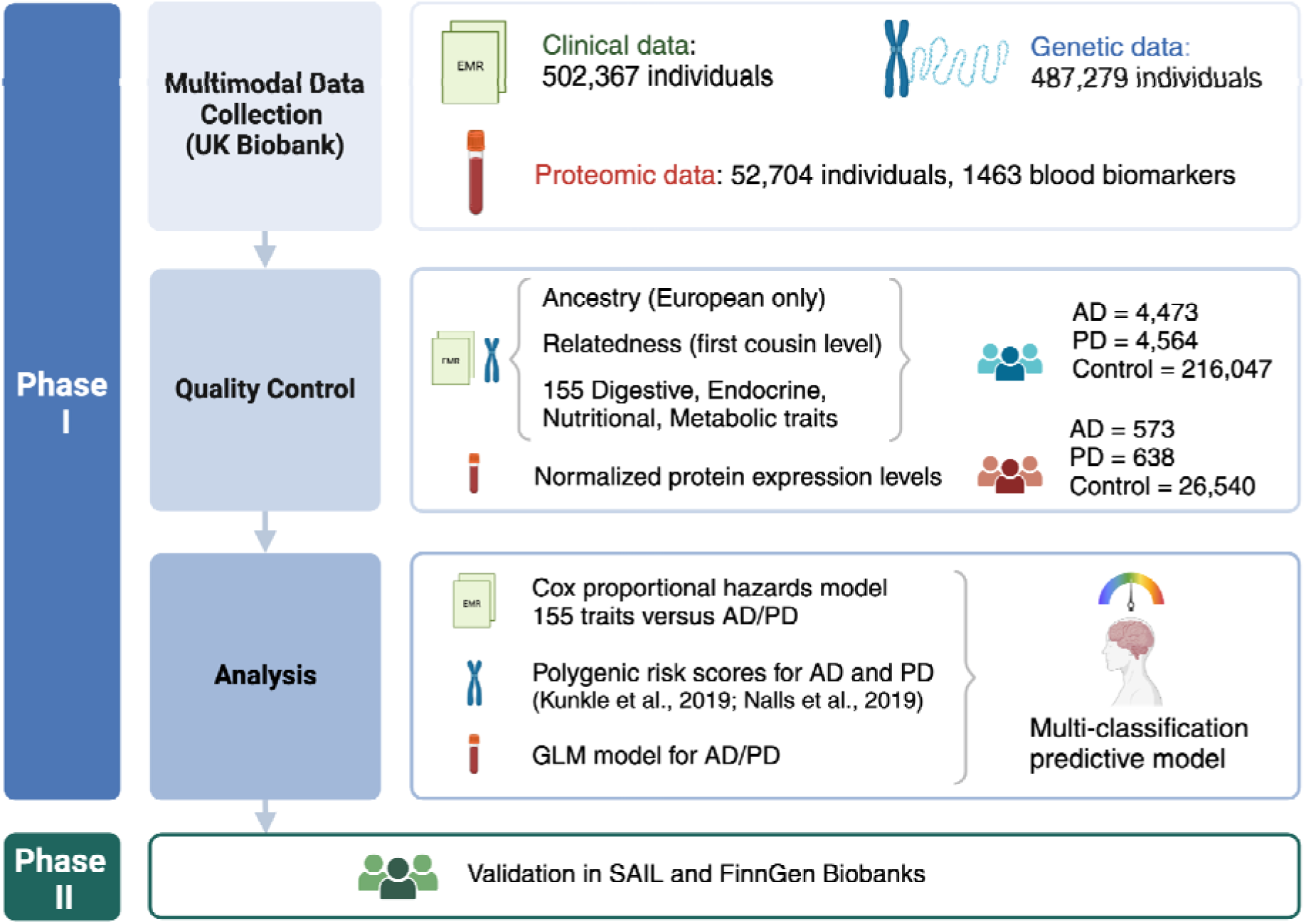
Study design. The initial phase of our study utilized clinical data sourced from electronic medical records (EMR) alongside genetic and proteomic data obtained from the UK Biobank. Quality control procedures were rigorously applied to clinical and genetic datasets, including filtering for individuals of European ancestry, exclusion of related samples, and extraction of 155 ICD-10 codes representing diagnoses related to digestive, endocrine, nutritional, and metabolic disorders. Proteomic data underwent normalization of protein expression levels as part of quality control measures. The culmination of this phase involved the application of a Cox proportional hazards model, examination of polygenic risk scores, and development of a generalized linear model (GLM). These analyses collectively contributed to the construction of a multi-classification predictive model for Alzheimer’s disease (AD) and Parkinson’s disease (PD). Phase II of our study entailed validating these findings using data from the Secure Anonymised Information Linkage (SAIL) and FinnGen biobanks.

### UK Biobank

The UKB data, accessed via DNAnexus under application number 33601, includes electronic health records of approximately 502,367 individuals, single nucleotide polymorphism (SNP) data of 487,279 individuals, and proteomic (Olink) data of 52,705 individuals (https://ukbiobank.dnanexus.com) (accessed on May 2023). The control group for the UKB dataset consisted of a subset of 352,610 individuals who have not been diagnosed with any neurodegenerative disease condition (table S10) and have no family history AD and PD diagnosis. For our approach, related individuals were filtered out from further analyses based on a kinship coefficient greater than 0.0884, and only individuals of European descent were selected for this study, based on field ID 22006, who self-identified as ’White British’ and confirmed through principal component (PC) analysis of the genotyping data to avoid potential confounding effects. We lacked a sufficient number of individuals with non-European ancestry to conduct a meaningful assessment. Our final dataset included 409,520 individuals of European ancestry. Endocrine, nutritional, metabolic, and digestive disorders, as well as AD and PD diagnosis, were derived from ICD-10 (International Classification of Diseases, 10th Revision) codes. The AD cohort was obtained from the G30 and F00 ICD-10 codes in the UKB, and the PD cohort was obtained from the G20 ICD-10 code. We excluded any individuals who had received an ICD-10 diagnosis for endocrine, nutritional, metabolic, and digestive disorders before January 1st, 1999, and right-censored any individuals who received an ICD-10 diagnosis after being diagnosed with AD or PD. Furthermore, any ICD-10 code with fewer than 5 cases was excluded from the analysis. Demographic characteristics are shown in table S11.

### SAIL Databank

The Secure Anonymised Information Linkage (SAIL) databank is a virtual platform providing anonymized medical data of the population in Wales (44). Diagnoses in SAIL are sourced from the Patient Episode Database for Wales (PEDW), records from clinicians and hospital staff, the Welsh Longitudinal General Practitioner dataset (WLGP), records from primary care physicians of diagnoses, treatments, symptoms, and referrals. Demographic information such as sex, age, address, and death were obtained from the Welsh Demographic Services Database (WDSD) and WLGP. Individuals with missing age or sex data and those without a Welsh address were excluded from further analysis. Diagnoses were identified in the PEDW using ICD-10 codes and in the WLGP using NHS read codes (CVT2,3). Neurodegenerative disorders from the outpatient data (OPDW) were excluded due to their minimal representation of dementia cases (only 0.1%) and the absence of reliable diagnosis dates. Similarly, dementia diagnoses from death records (ADDES) were not included due to inaccurate diagnosis dates. This study covered the period from January 1, 1999, to December 31, 2018. For inclusion, individuals were required to have been alive at the start of 1999 and to have been at least 45 years old on January 1, 1999.

### FinnGen Biobank

The FinnGen study is a large-scale genomics initiative that has analyzed over 500,000 Finnish biobank samples and correlated genetic variation with health data to understand disease mechanisms and predispositions. The project is a collaboration between research organisations and biobanks within Finland and international industry partners (45). FinnGen provides survival analyses across numerous clinical endpoints. The hazard ratios are adjusted for sex and year of birth. FinnGen bases the calculation of these hazard ratios on a wide array of clinical endpoints defined through data from nationwide registries, including, but not limited to, Statistics Finland [https://finngen.gitbook.io/documentation/methods/endpoints]. We downloaded the hazard ratios for AD/PD from FinnGen’s Risteys R10 platform for AD using the G6_AD_WIDE category matching more closely the UKB grouping than G6_AD [https://r10.risteys.finngen.fi/endpoints/G6_AD_WIDE] and for PD [https://r10.risteys.finngen.fi/endpoints/G6_PARKINSON] to explore the putative impact of endocrine, digestive, metabolic, and nutritional disorders on the risk of of AD/PD prior to diagnosis. It is important to note that not all ICD-10 codes used in our discovery phase in the UKB are represented in the FinnGen dataset. This discrepancy is perhaps due to the differences in health registries and data collection methodologies between the UKB and FinnGen biobanks.

### Polygenic Risk Score Analyses

Risk allele loci and beta values from GWAS summary statistics, specifically 23 risk predictors linked to AD risk from Kunkle 2019 and 90 risk predictors associated with PD from Nalls 2019, were used to estimate PRS for AD and PD respectively (46,47). Each risk allele was assigned a weight based on the magnitude of its effect in the published studies, giving greater emphasis to alleles with higher risk estimates. Genetic variants were extracted for each individual from the UKB imputed data using the ’bgenix’ package (48). The extracted variants were then converted to binary formats and used to compute the PRS score for each individual using PLINK 2.0 (49). Estimated profiles were then normalized to Z-scores based on a control reference.

### Cox Proportional Hazard Model

A Cox proportional hazards model was used to calculate the hazard ratio between risk for incident AD and PD and endocrine, nutritional, metabolic, and digestive system disorders. The disorders under study are represented as 155 ICD-10 codes (table S12). In this model, January 1st, 1999 was used as the cutoff date for the diagnosis of these conditions. Consequently, any individual diagnosed with these traits before January 1st, 1999 was excluded from the analysis. Logistic regressions and Cox proportional hazards models were adjusted for age, sex, the Townsend deprivation index, and 5 PCs to account for population stratification (precomputed in UKB only). Additionally, we conducted Time-Stratified Cox Regression Analysis, for which cohorts were divided into three strata based on ICD-10 codes: 1-5 years, 5-10 years, and 10-15 years prior to NDD diagnosis. ICD-10 diagnoses within the specified time periods were retained, while any ICD-10 codes outside of these time frames were converted to NaN.

### Polygenic Risk Score distribution for Alzheimer’s disease and Parkinson’s disease

A t-test was used to quantify the differences in the distribution of genetic alleles for AD or PD between individuals diagnosed only with AD or PD and individuals with an additional, significant ICD-10 diagnosis. This approach measures how much the cumulative effect of these alleles varies between the two groups. Additionally, it unveils the genetic risk profiles for AD and PD in both groups. By calculating and comparing PRSs based on alleles associated with AD or PD, the analysis evaluates how the genetic predisposition to AD or PD is increased or decreased among those individuals diagnosed with one of the ICD-10 codes in addition to AD or PD. This comparison aids in understanding whether genetic risk for AD or PD is similar or distinct in the compared subgroups of study.

### Statistical Analysis accounting for the Impact of APOE, LRRK2, and GBA1 risk variants

PRS analyses were adjusted for major genetic risk factors associated with AD and PD to determine if observed similarities or differences between AD/PD and the assessed ICD-10 codes could be attributed to pleiotropic effects. For AD, individuals homozygous for the ’C’ allele at both APOE rs429358 and rs7412 were identified as having two copies of the APOE-ε4 allele, coded as ’2’, and one copy of the APOE-ε4 allele coded as ‘1’ in our regression analysis to indicate a higher genetic risk. All other configurations were coded as ’0’. For PD, we examined LRRK2 at rs76904798, where homozygous ’C’ alleles were coded as ’0’, and heterozygous and homozygous for ’T’ alleles as ’1’. Similarly, at rs34637584 (LRRK2 G2019S), we applied ’0’ for homozygous ’G’ alleles, and ’1’ for heterozygous and homozygous ’A’ alleles. For GBA1, at rs35749011 (proxy for GBA1 E326K), ’0’ was assigned to homozygous ’G’ alleles, ’1’ to heterozygous and homozygous ’A’ alleles, and at rs76763715 (GBA1 N370S), ’0’ for homozygous ’T’, and ’1’ for heterozygous and homozygous ’C’ alleles.

### Interaction model for genetic risk across endocrine, metabolic, digestive system disorders and nutritional status

We aimed to understand how the interplay between genetics underlying AD and PD risk and a clinical diagnosis for any investigated ICD-10 code could eventually influence an AD or PD diagnosis. A Generalized Linear Model (GLM) was used to account for more complex relationships, where the impact of genetic risk for AD or PD (as measured by PRS) might interact differently across the diagnoses under study represented by ICD-10 codes. The interaction term used in this model was: Z-score * ICD-10 terms adjusted by sex, age and Townsend deprivation index.

### Proteomic Biomarker Data Analyses

We aimed to explore differences in the levels of proteomic biomarkers associated with AD or PD between individuals with and without co-occurring ICD-10 code diagnoses related to endocrine, nutritional, metabolic, and digestive system disorders among UKB participants. For this purpose, we utilized data from the Pharma Proteome Project, which provides thousands of plasma protein biomarkers in blood samples (https://olink.com/news/uk-biobank-pharma-proteomics-project/). In a cross-sectional analysis, we focused on baseline (Instance_0) proteomics data, as it includes measurements for the largest number of individuals, encompassing data for 52,705 individuals (at the time of data access) and a total of 1,463 proteins (table S13). These biomarkers span across cardiometabolics, inflammation, neurology, and oncology markers.

We applied GLM to analyze 1,463 available biomarkers on AD and PD risk. The regressions were adjusted for age at recruitment, Townsend deprivation index, sex, and 5 PCs. Subsequently, we selected False Discovery Rate (FDR)-corrected significant proteomic biomarkers from the GLM results. Additionally, we compared the average levels of proteomic biomarkers in isolated cases of AD/PD with cases of AD/PD co-occurring with selected ICD-10 codes (found to be significantly associated with AD or PD risk before AD/PD diagnosis). We selected FDR-corrected significant proteomic biomarkers with an OR greater than 1 and applied a t-test to compare their average levels in the isolated cases of AD or PD and cases co-occurring with specific ICD-10 codes.

### Multi-modal classification model for clinical, genetics and proteomics on Alzheimer’s disease and Parkinson’s disease risk

To evaluate the role of endocrine, metabolic, digestive and nutritional status-related diagnoses in predicting AD or PD status, we developed a multi-modal classification model on a subset of the UK Biobank dataset. This subset included individuals with variables encompassing clinical diagnosis for endocrine, metabolic, digestive and nutritional status; biomarkers; demographic factors (age at recruitment, sex, and Townsend deprivation index); derived variables (Z score for the PRS); PCs; and APOE status for AD, or LRRK2/GBA1 status for the PD model. The clinical diagnoses used in these models include the ICD-10 codes found to have significant HRs in UKB. We employed a machine learning approach to compare the predictive performance of various feature sets on AD/PD outcomes. These included genetics, clinical, proteomic, demographics (including sex, age at recruitment), and combinations thereof. Each dataset was independently analyzed using a Gradient Boosting Classifier, an ensemble learning method for classification tasks [https://scikit-learn.org/stable/modules/generated/sklearn.ensemble.GradientBoostingClassifier.html].

We conducted hyperparameter tuning through GridSearchCV, optimizing for the number of estimators, learning rate, and maximum depth. For AD classification, the grid search adjusts XGBClassifier parameters including n_estimators (2, 3, 5, 10, 15), learning_rate (0.001, 0.01, 0.1), and max_depth (3, 4, 5). For PD, it adjusts n_estimators (300, 500), learning_rate (0.005, 0.01), and max_depth (1, 2, 3). Both models use nested cross-validation, optimizing based on Receiver Operating Characteristic Area Under the Curve (ROC AUC). We used 5 folds for both inner and outer cross-validation. Feature selection was performed using the Least Absolute Shrinkage and Selection Operator (LASSO) as a part of the hyperparameter tuning procedure. The dataset was downsampled to have an equal number of cases and controls, and then scaled using StandardScaler to ensure uniformity and prevent bias due to variance in measurement scales before classification training. Model performance was evaluated based on ROC AUC and balanced accuracy (BA) scores. The 95% confidence intervals (CI) were calculated based on performance metrics obtained from a 5-fold cross-validation. We employed the Shapley Additive Explanations (SHAP) approach to assess the impact of each feature on the machine learning model predictions (50). SHAP values are derived from game theory and approximate a feature’s effect on the model. SHAP enhances understanding by creating accurate explanations for each observation. The SHAP package was used to calculate and visualize these Shapley values seen in the figures in the manuscript and the interactive website. A surrogate LightGBM regression model (https://lightgbm.readthedocs.io/) was trained on the risk estimates to calculate SHAP values.

## Data Availability

This paper analyzes existing, publicly available data. In addition, complete summary statistics describing these data/processed datasets derived from these data have been deposited in the supplementary materials connected to this publication and are publicly available as of the date of publication.

https://github.com/NIH-CARD/Gut-Brain-Nexus

https://gut-brain-nexus.streamlit.app/

## Funding

Intramural Research Program of the NIH, National Institute on Aging (NIA), National Institutes of Health, Department of Health and Human Services; project number ZO1 AG000534

National Institute of Neurological Disorders and Stroke

Dementia Research Institute [UKDRI supported by the Medical Research Council (UKDRI-3003), Alzheimer’s Research UK, and Alzheimer’s Society]

Welsh Government Joint Programming for Neurodegeneration (MRC: MR/T04604X/1) Dementia Platforms UK (MRC: MR/L023784/2)

MRC Centre for Neuropsychiatric Genetics and Genomics (MR/L010305/1)

## Author contributions

Conceptualization: MS, SBC, KSL

Data acquisition and processing: MS, ES, KSL, HLL, CB, HI, MAN

Data analyzation: MS, ES, KSL

Interpretation: All authors contributed

Writing: All authors contributed

## Competing interests

KSL, HLL, HI, and MAN’s participation in this project was part of a competitive contract awarded to Data Tecnica International LLC by the National Institutes of Health to support open science research. MAN also currently serves on the scientific advisory board at Clover Therapeutics and is an advisor and scientific founder at Neuron23 Inc. All other authors declare they have no competing interests.

## Data and materials availability

Notebooks containing code used in this analysis can be found in the GitHub link here: [https://github.com/NIH-CARD/Gut-Brain-Nexus]. This paper analyzes existing, publicly available data. In addition, complete summary statistics describing these data/processed datasets derived from these data have been deposited in the supplementary materials connected to this publication and are publicly available as of the date of publication. Further, we have developed an interactive resource for the scientific community [https://gut-brain-nexus.streamlit.app/] where researchers can investigate components of the predictive model and can investigate feature effects on a sample level.

## Notes

### Competing Interest Statement

K.S.L., H.L.L., H.I., M.B.M., and M.A.N.'s participation in this project was part of a competitive contract awarded to Data Tecnica International LLC by the National Institutes of Health to support open science research. M.A.N. also currently serves on the scientific advisory board at Clover Therapeutics and is an advisor and scientific founder at Neuron23 Inc.

### Funding Statement

This research was supported in part by the Intramural Research Program of the NIH, National Institute on Aging (NIA), National Institutes of Health, Department of Health and Human Services; project number ZO1 AG000534, as well as the National Institute of Neurological Disorders and Stroke. This work was also supported by the Dementia Research Institute [UKDRI supported by the Medical Research Council (UKDRI-3003), Alzheimer's Research UK, and Alzheimer's Society], Welsh Government, Joint Programming for Neurodegeneration (MRC: MR/T04604X/1), Dementia Platforms UK (MRC: MR/L023784/2) and MRC Centre for Neuropsychiatric Genetics and Genomics (MR/L010305/1).

### Summary of Updates

Revised to include updated classification models' performances and the interactive web application for investigating our predictive models.

