## Supplemental Figures for "Gut-Brain Nexus: Mapping Multi-Modal Links to Neurodegeneration at Biobank Scale"

**Supplementary Figure 1**

| Prior_Condition_Description | HR | 95% CI | P_VAL |
|---|---|---|---|
| Other Disorders of Peritoneum | 0.59 | 0.44 - 0.81 | 8.77e-04 |
| Haemorrhoids and Perianal Venous Thrombosis | 0.69 | 0.59 - 0.80 | 6.94e-07 |
| Obesity | 0.84 | 0.74 - 0.96 | 9.29e-03 |
| Disorders of Lipoprotein Metabolism and Other Lipidaemias | 1.18 | 1.09 - 1.27 | 1.53e-05 |
| Gastritis and Duodenitis | 1.20 | 1.09 - 1.32 | 1.45e-04 |
| Oesophagitis | 1.28 | 1.10 - 1.49 | 1.63e-03 |
| Disorders of Mineral Metabolism | 1.32 | 1.08 - 1.62 | 7.75e-03 |
| Other Non-Infective Gastro-Enteritis and Colitis | 1.37 | 1.20 - 1.56 | 1.75e-06 |
| Diseases of Pulp and Periapical Tissues | 1.44 | 1.11 - 1.87 | 6.21e-03 |
| Non-Insulin-Dependent Diabetes Mellitus | 1.47 | 1.33 - 1.63 | 1.19e-14 |
| Other Disorders of Fluid, Electrolyte and Acid-Base Balance | 1.51 | 1.34 - 1.70 | 6.43e-12 |
| Other Bacterial Intestinal Infections | 1.51 | 1.21 - 1.89 | 2.35e-04 |
| Unspecified Diabetes Mellitus | 1.53 | 1.33 - 1.75 | 1.21e-09 |
| Other Functional Intestinal Disorders | 1.55 | 1.40 - 1.73 | 3.55e-16 |
| Deficiency of Other b Group Vitamins | 1.78 | 1.40 - 2.27 | 2.61e-06 |
| Volume Depletion | 1.78 | 1.52 - 2.10 | 3.26e-12 |
| Vitamin d Deficiency | 1.95 | 1.57 - 2.41 | 9.97e-10 |
| Other Disorders of Pancreatic Internal Secretion | 2.23 | 1.75 - 2.83 | 5.27e-11 |
| Amyloidosis | 2.57 | 1.34 - 4.94 | 4.72e-03 |
| Insulin-Dependent Diabetes Mellitus | 3.09 | 2.45 - 3.90 | 1.77e-21 |

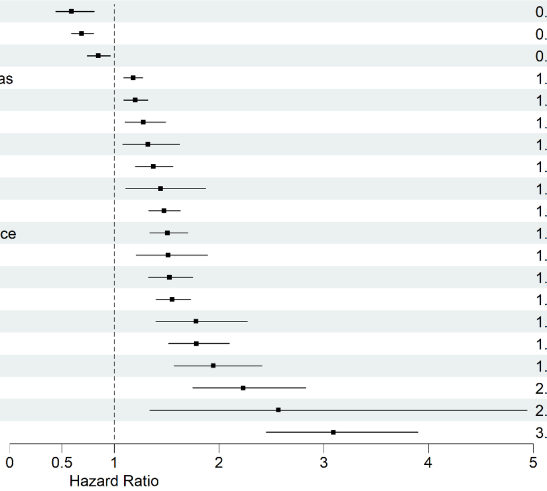


**Fig. S1**

Forest plot showing hazard ratios for the prior ICD-10 diagnoses that are significantly associated with the risk of Alzheimer’s Disease (AD). (AD ~ ICD-10 + sex + age + Townsend deprivation index)

**Supplementary Figure 2**

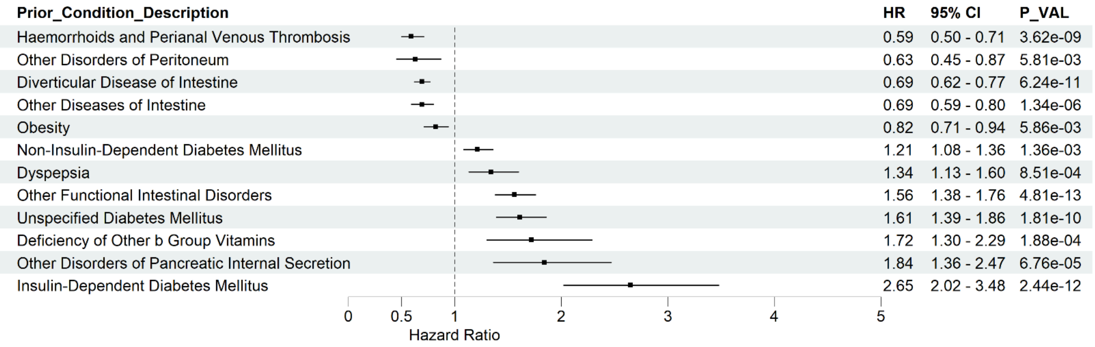


**Fig. S2**

Forest plot showing hazard ratios for the prior ICD-10 diagnoses that are significantly associated with the risk of Parkinson’s Disease (PD). (PD ~ ICD-10 + sex + age + Townsend deprivation index).

Supplementary Figure 3A

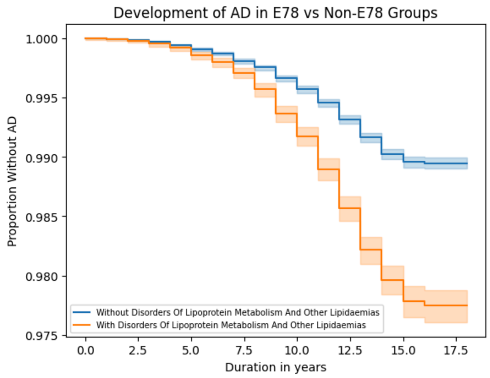


Supplementary Figure 3B

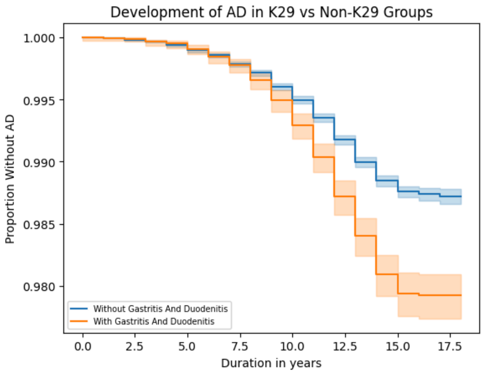


Supplementary Figure 3C

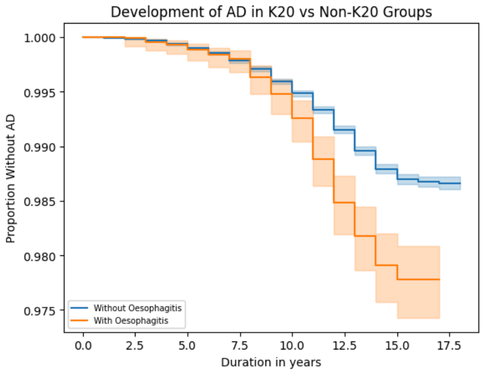


Supplementary Figure 3D

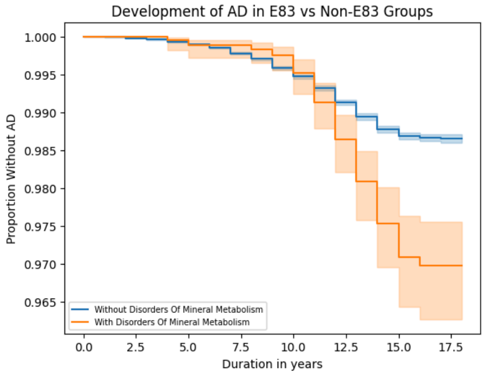


Supplementary Figure 3E

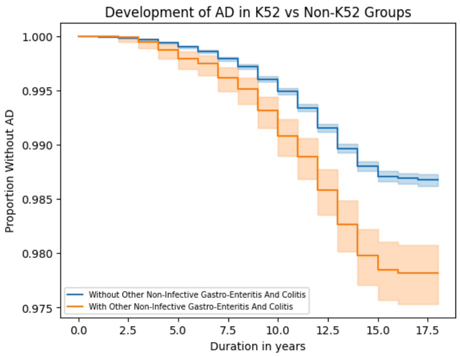


Supplementary Figure 3F

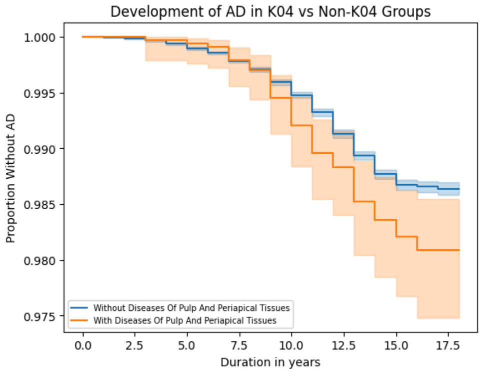

Supplementary Figure 3G

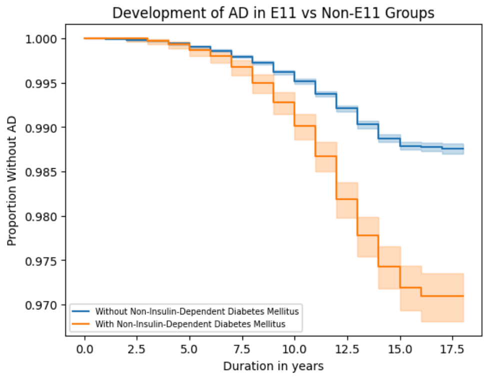


Supplementary Figure 3H

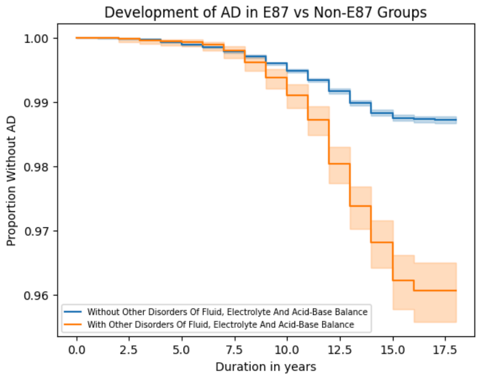


Supplementary Figure 3I

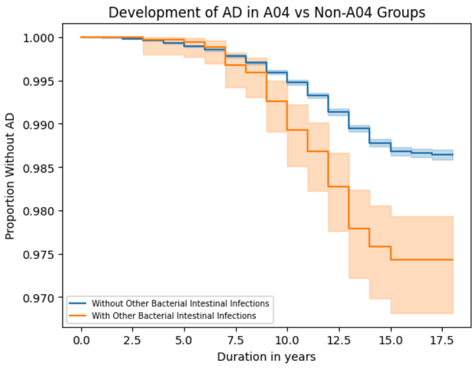


Supplementary Figure 3J

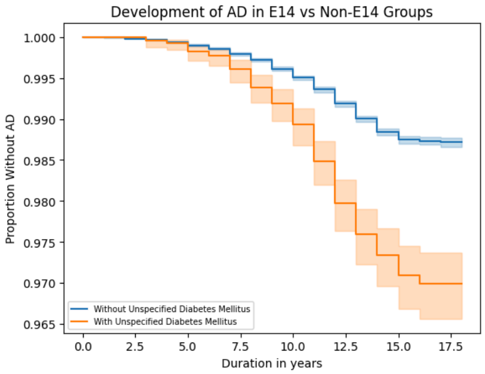


Supplementary Figure 3K

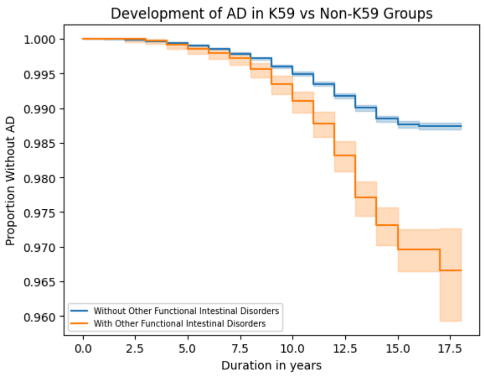


Supplementary Figure 3L

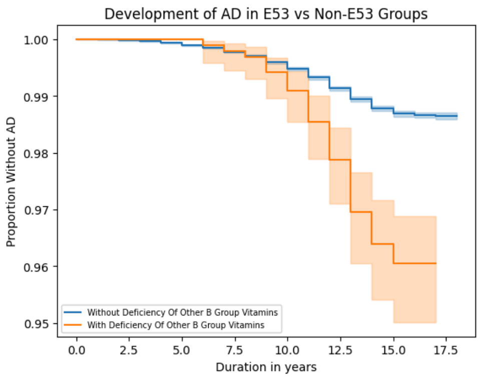

Supplementary Figure 3M

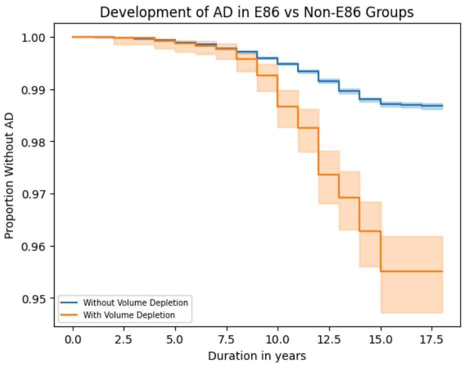


Supplementary Figure 3N

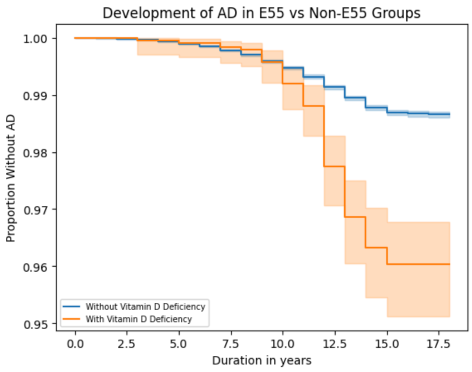


Supplementary Figure 3O

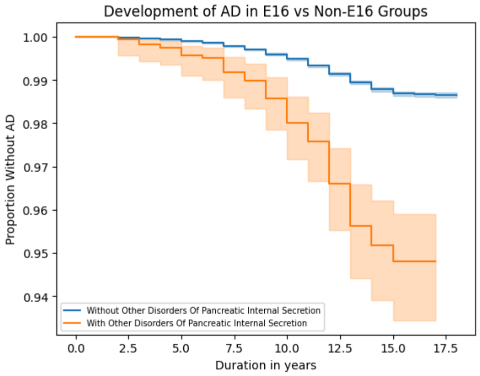


Supplementary Figure 3P

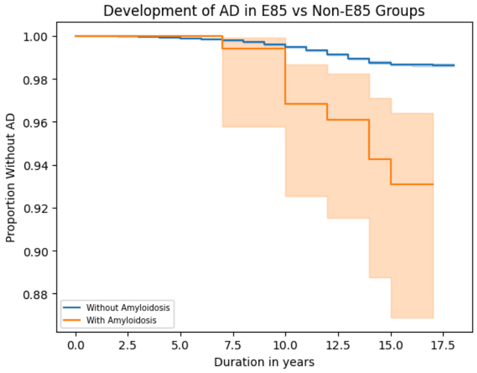


Supplementary Figure 3Q

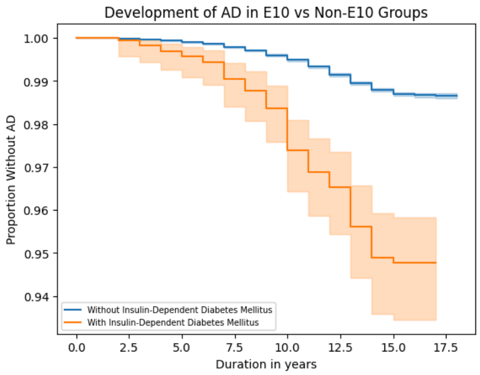


**Fig. S3** Kaplan-Meier plot displaying the probabilities of developing Alzheimer's Disease for individuals categorized by specific diagnoses

Supplementary Figure 4A

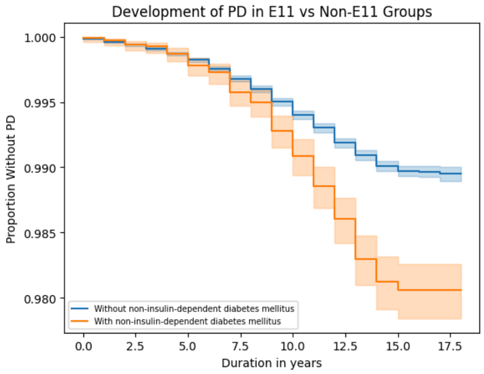


Supplementary Figure 4B

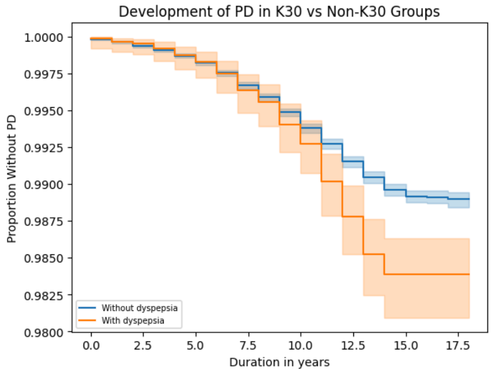


Supplementary Figure 4C

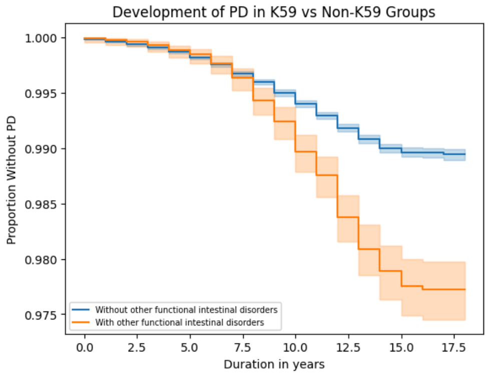


Supplementary Figure 4D

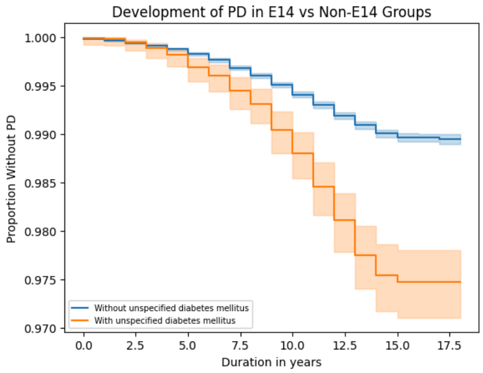


Supplementary Figure 4E

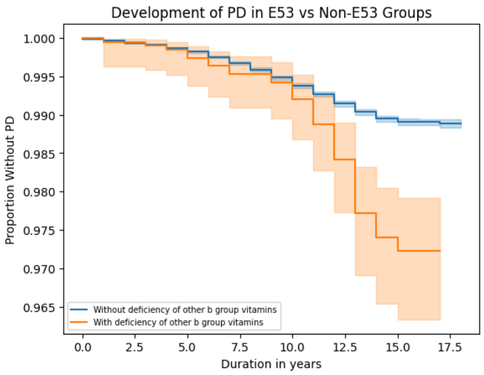


Supplementary Figure 4F

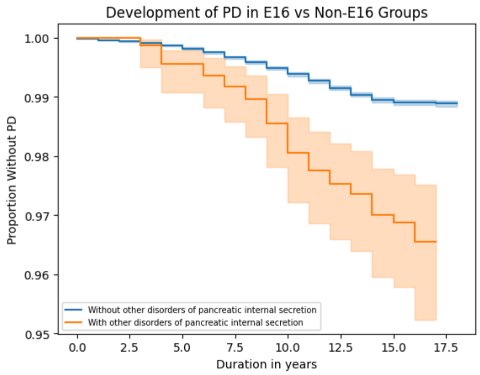

Supplementary Figure 4G

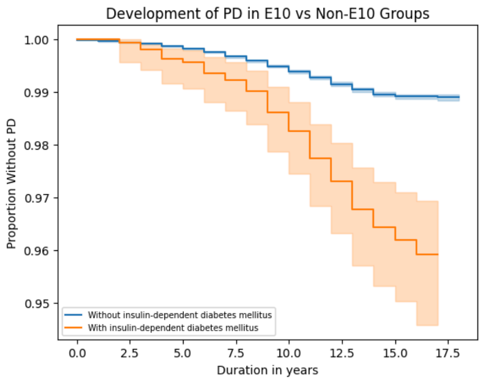


**Fig. S4** Kaplan-Meier plot displaying the probabilities of developing Parkinson's Disease for individuals categorized by specific diagnoses of a disorder affecting the gut-brain axis.

Supplementary Figure 5A

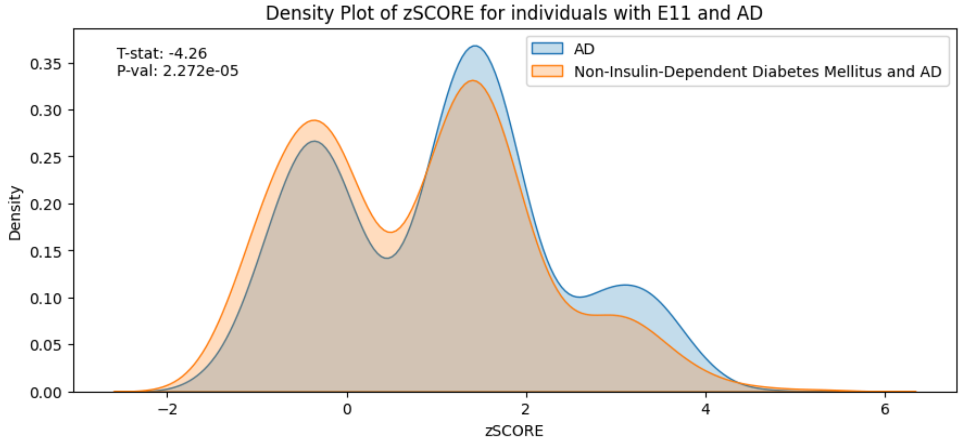


Supplementary Figure 5B

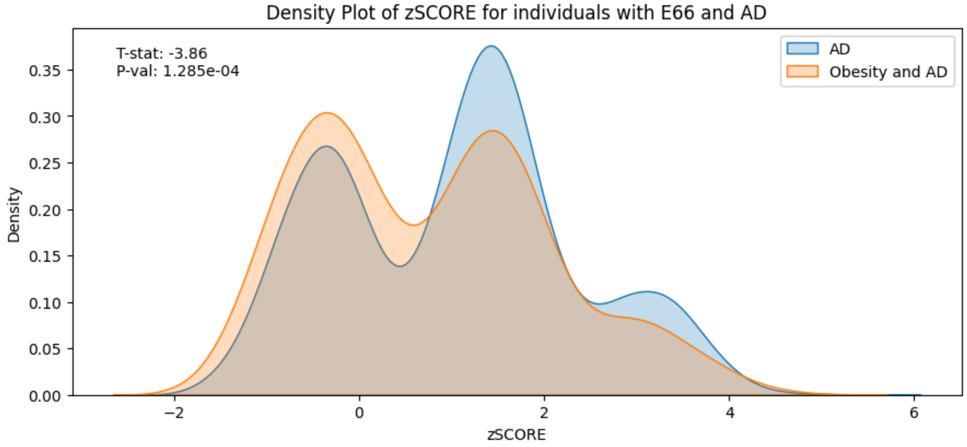


Supplementary Figure 5C

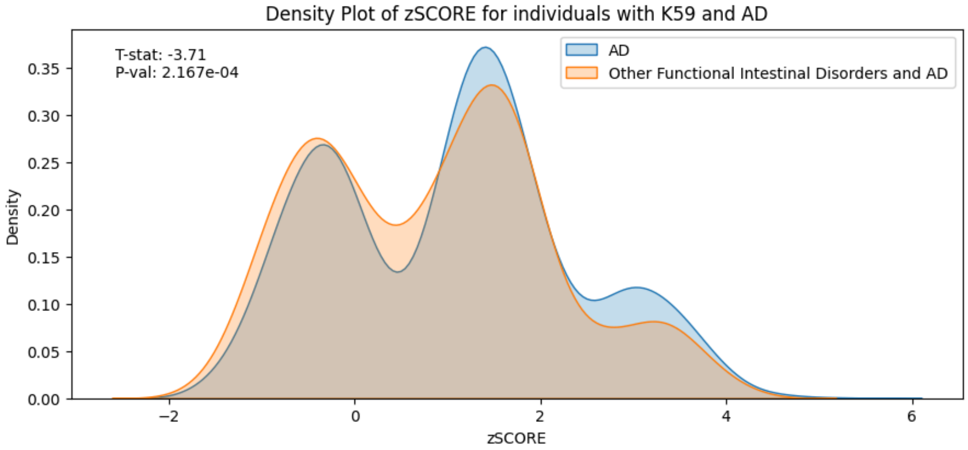

Supplementary Figure 5D

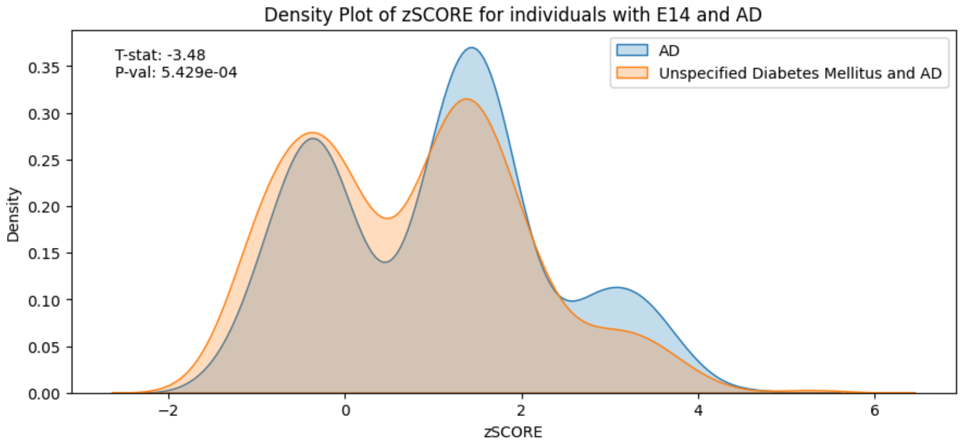


Supplementary Figure 5E

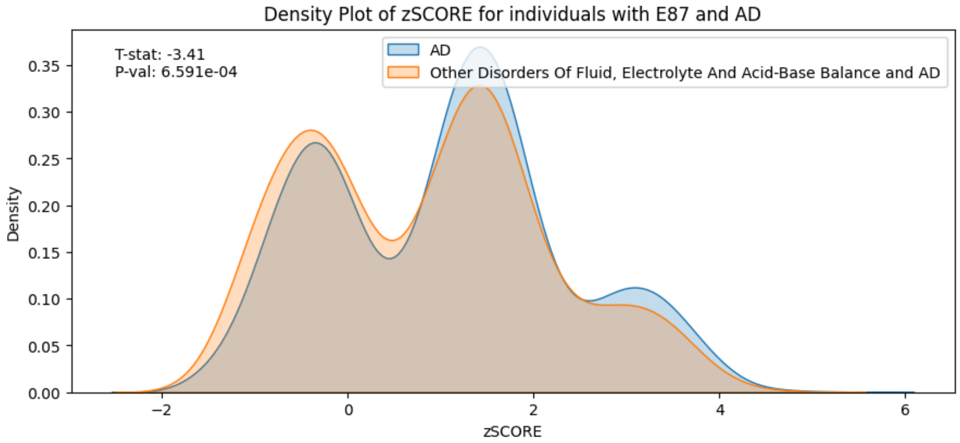


Supplementary Figure 5F

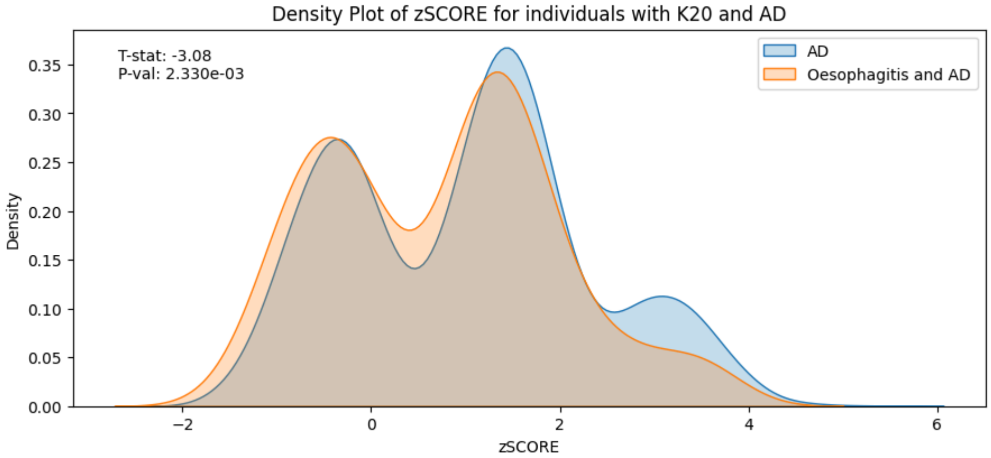

Supplementary Figure 5G

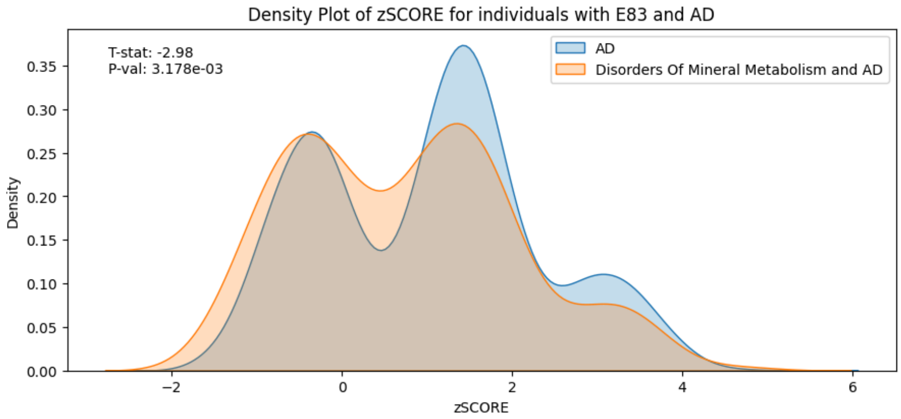


Supplementary Figure 5H

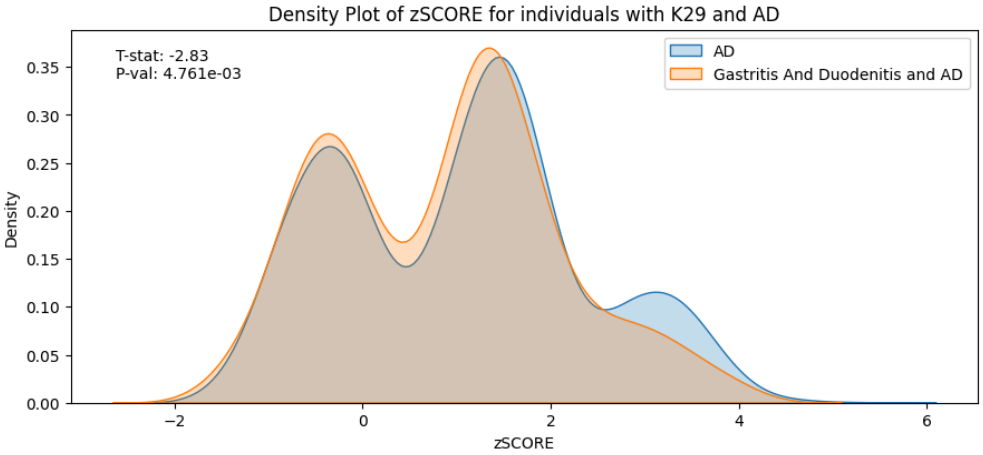


Supplementary Figure 5I

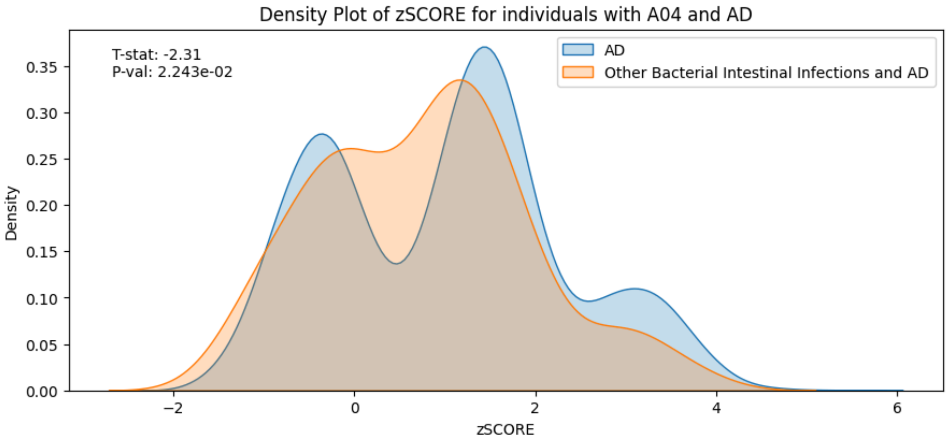

Supplementary Figure 5J

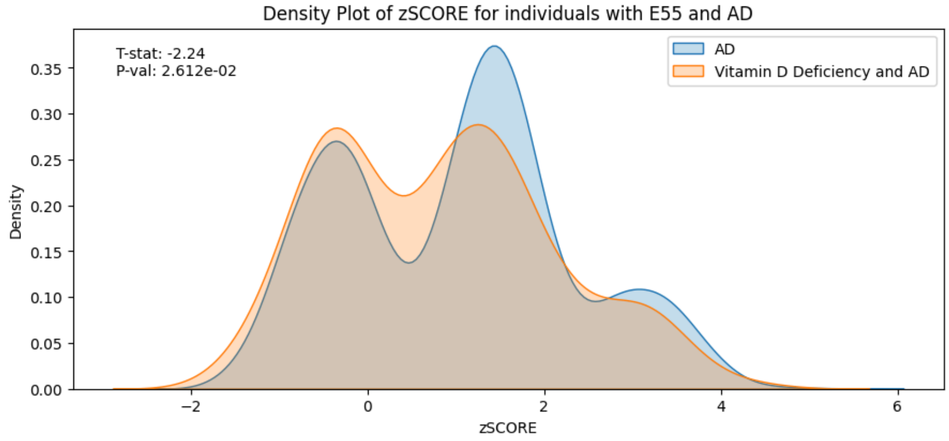


Supplementary Figure 5K

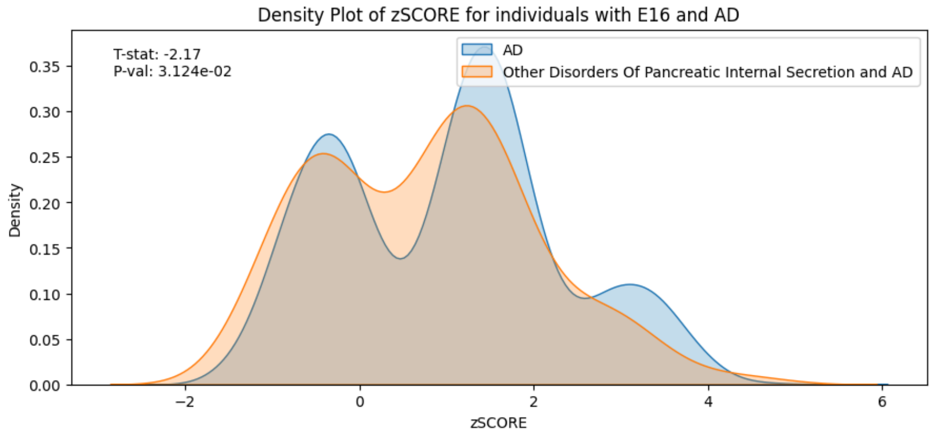


Supplementary Figure 5L

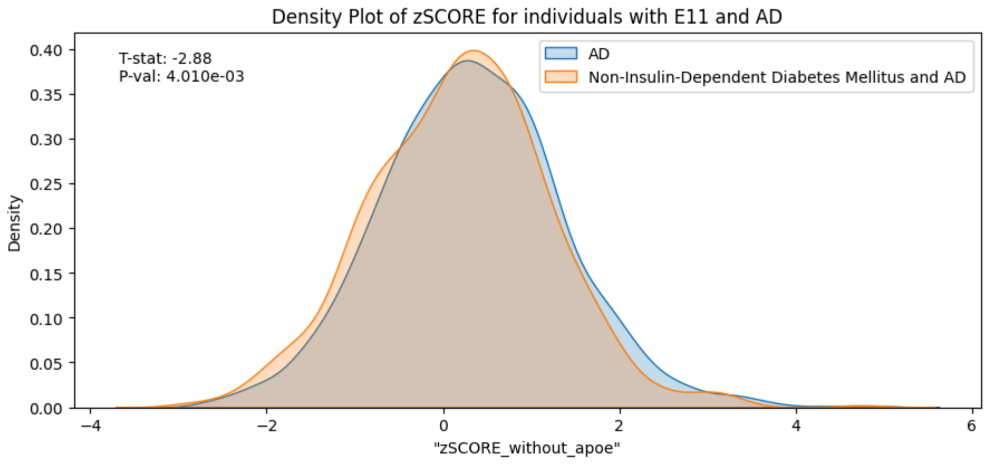

Supplementary Figure 5M

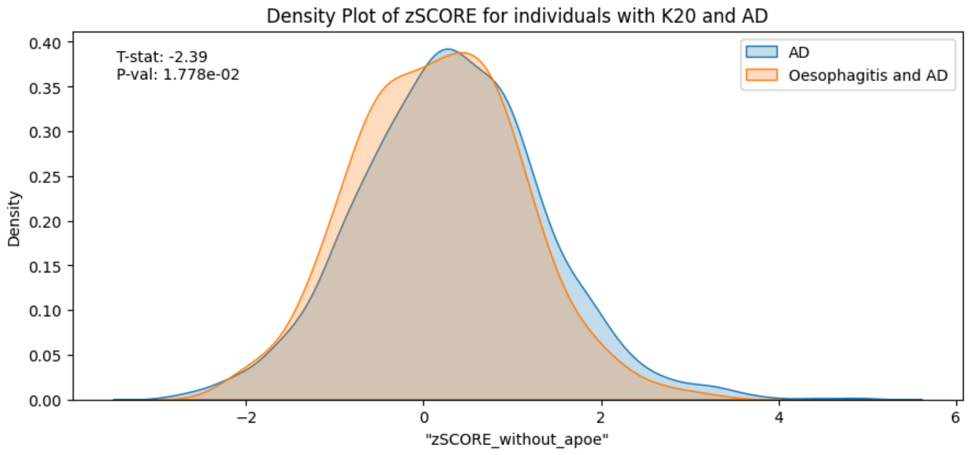


Supplementary Figure 5N

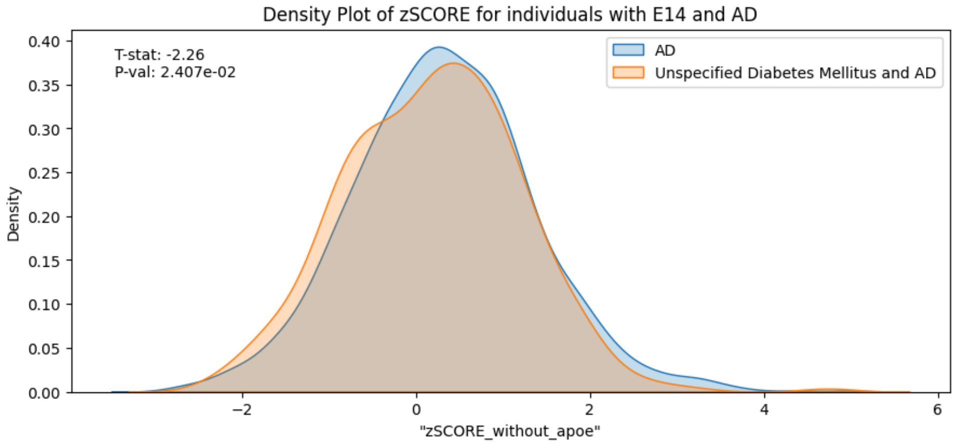


Supplementary Figure 5O

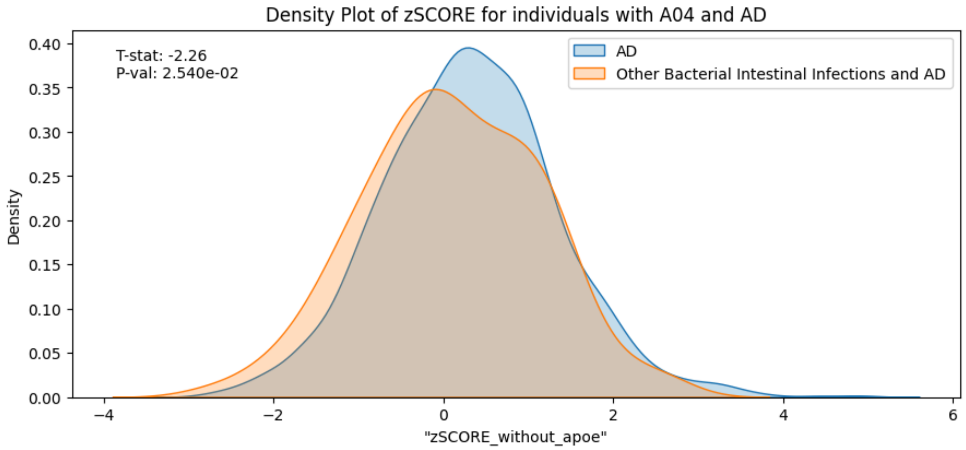

Supplementary Figure 5P

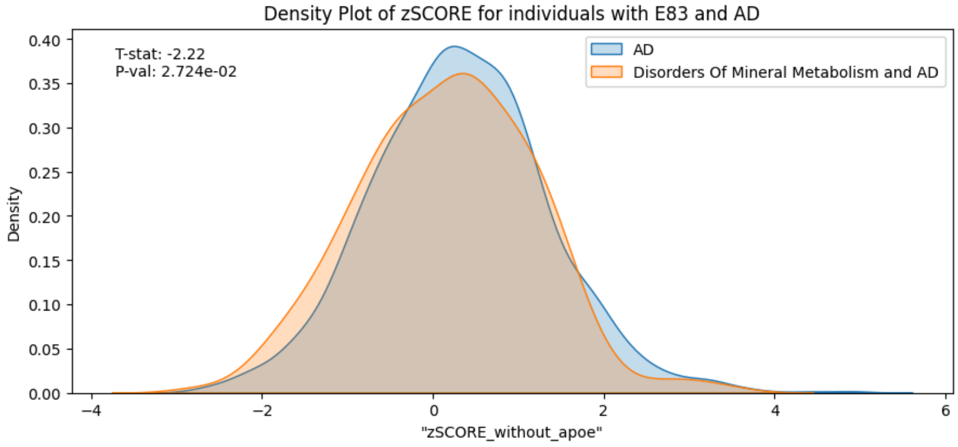


**Fig. S5** Density plots of Alzheimer's Disease (AD) polygenic risk scores (PRS) with *APOE*, comparing cases with and without prior diagnoses of a disorder affecting the gut-brain axis. AD PRS is distributed differently between cases of AD with and without prior ICD-10 Diagnosis.

Supplementary Figure 6A

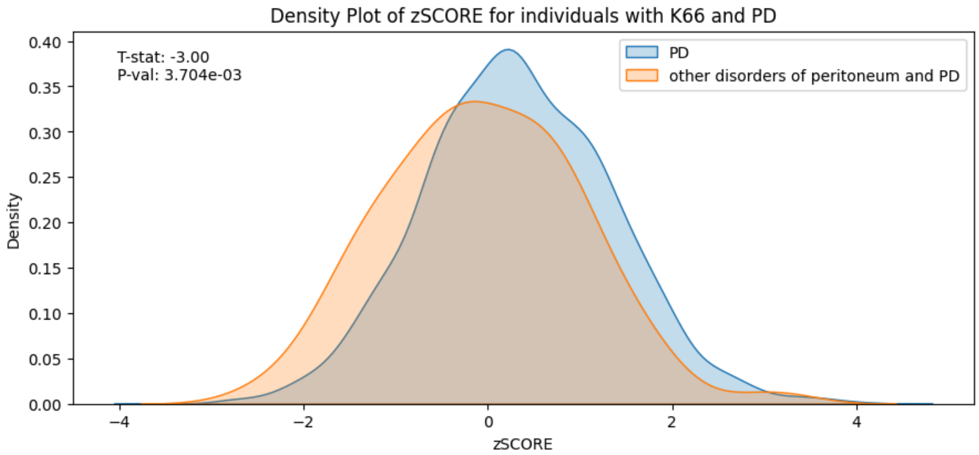


Supplementary Figure 6B

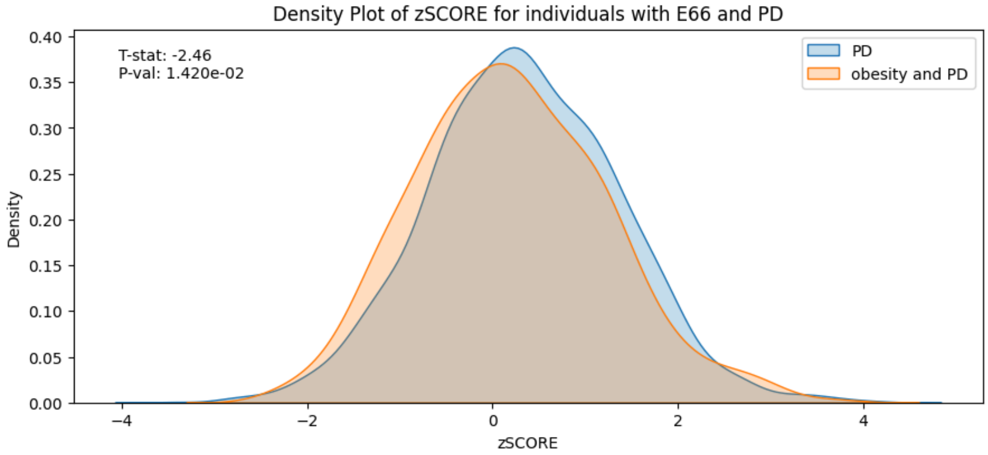


Supplementary Figure 6C

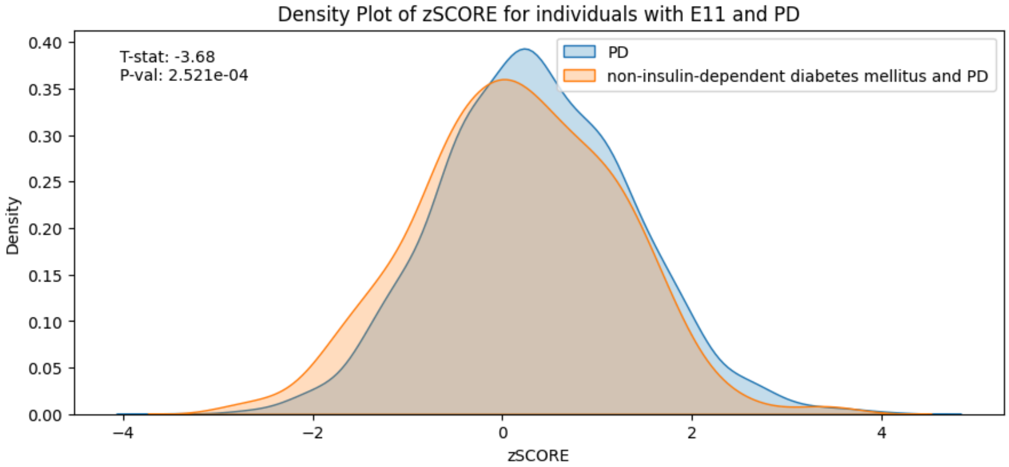

Supplementary Figure 6D

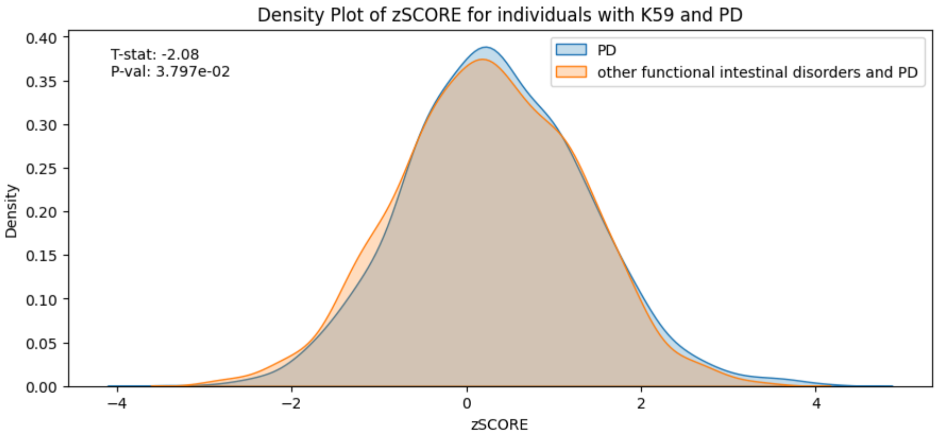


Supplementary Figure 6E

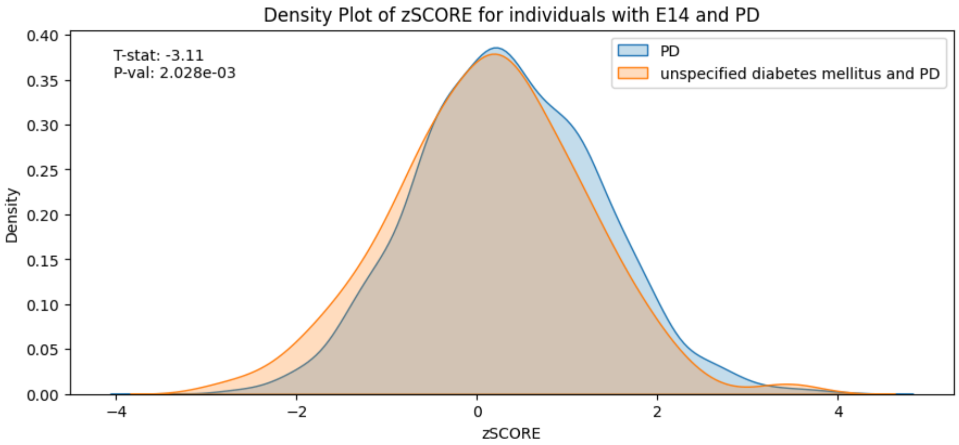


Supplementary Figure 6F

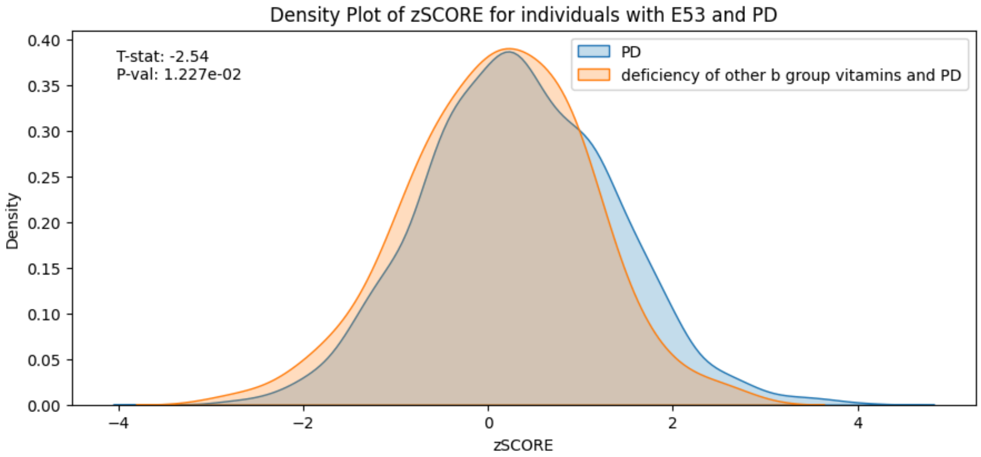

Supplementary Figure 6G

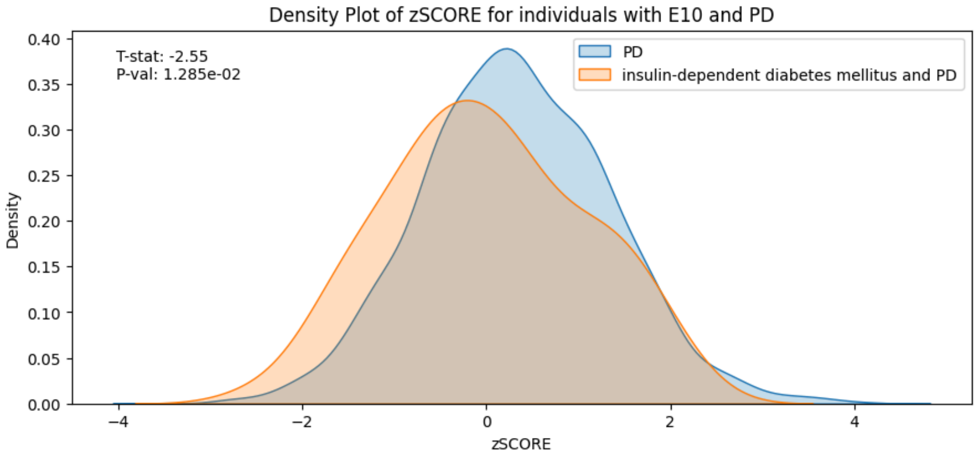


**Fig. S6** Density plots of Parkinson's Disease (PD) polygenic risk scores (PRS) comparing cases with and without prior diagnosis of a disorder affecting the gut-brain axis. AD PRS is distributed differently between cases of AD with and without prior ICD-10 Diagnosis.

Figure 7A.1

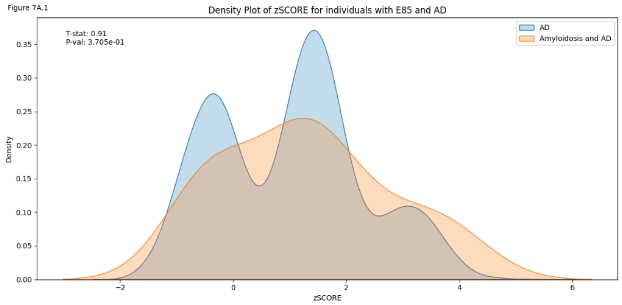


Figure 7A.2

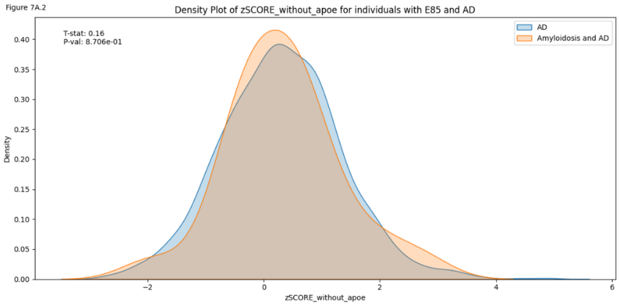


Figure 7B.1

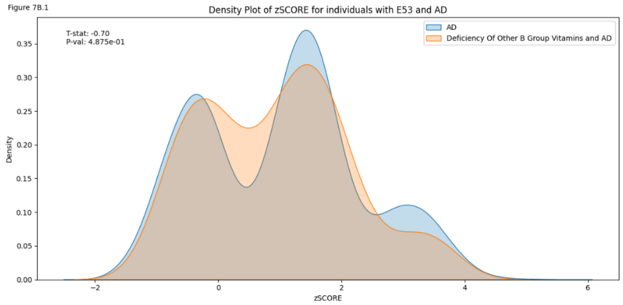


Figure 7B.2

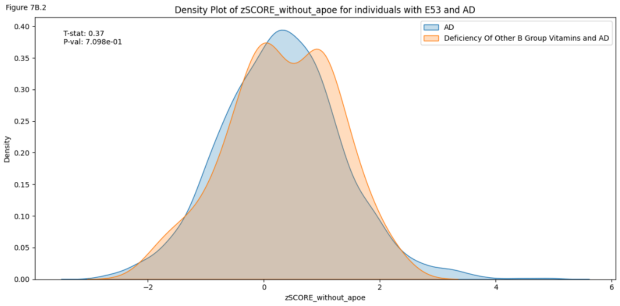


Figure 7C.1

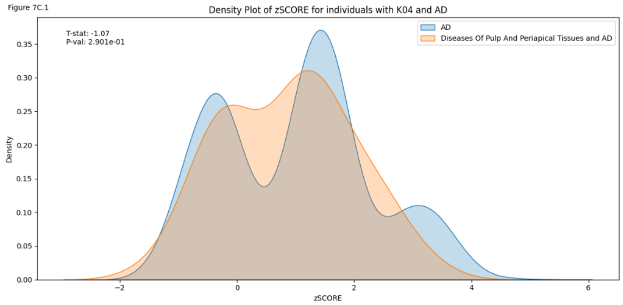


Figure 7C.2

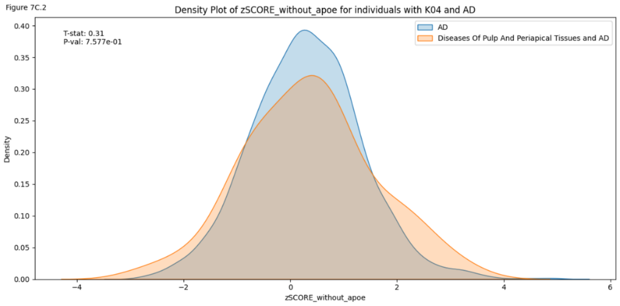


Figure 7D.1

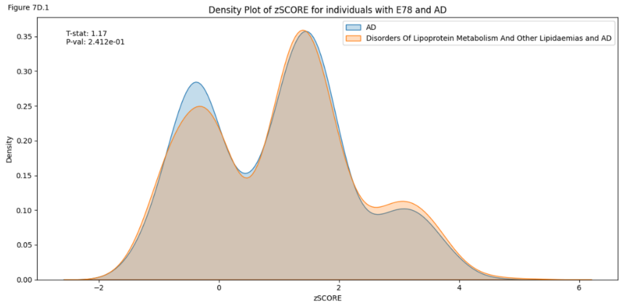


Figure 7D.2

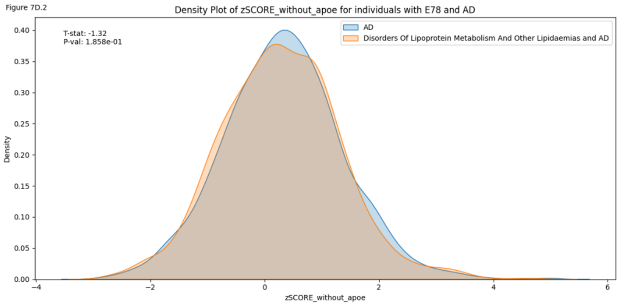


Figure 7E.1

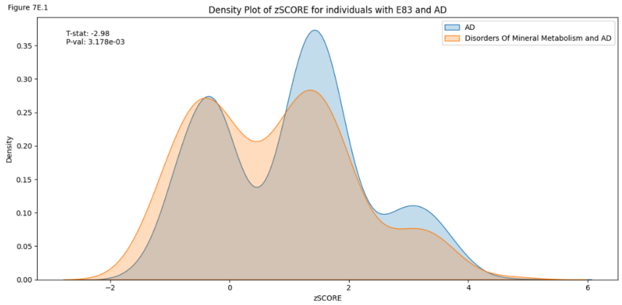


Figure 7E.2

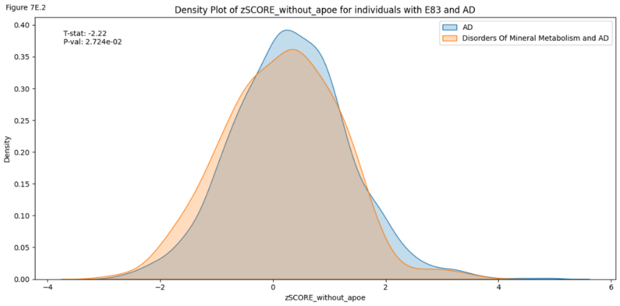

Figure 7F.1

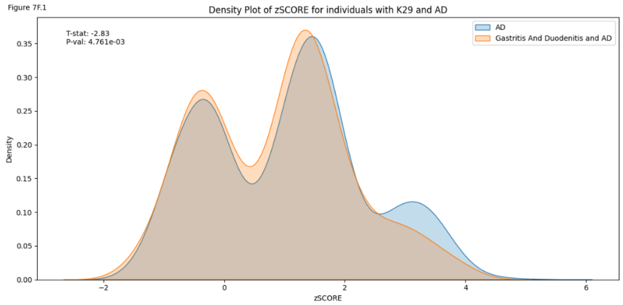


Figure 7F.2

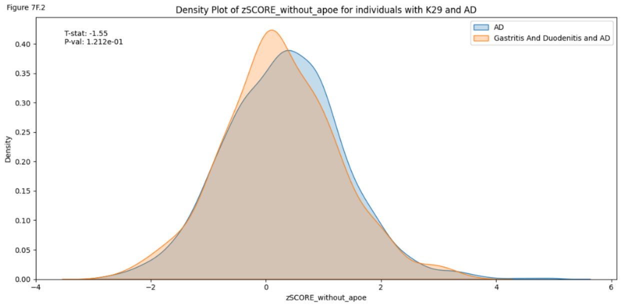


Figure 7G.1

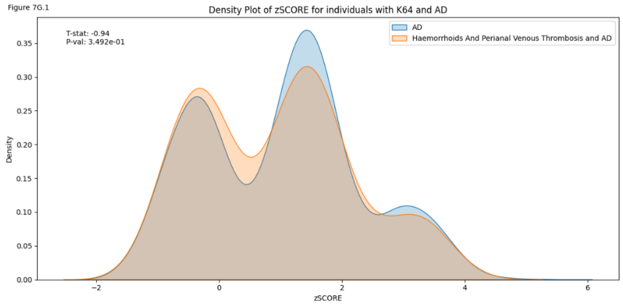


Figure 7G.2

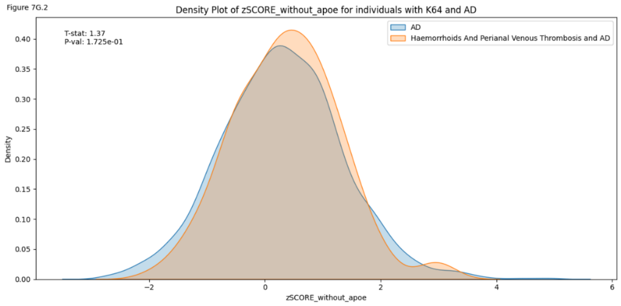


Figure 7H.1

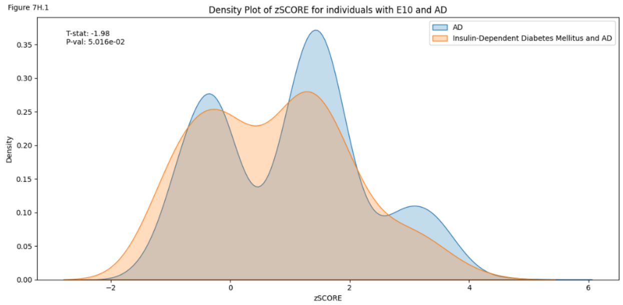


Figure 7H.2

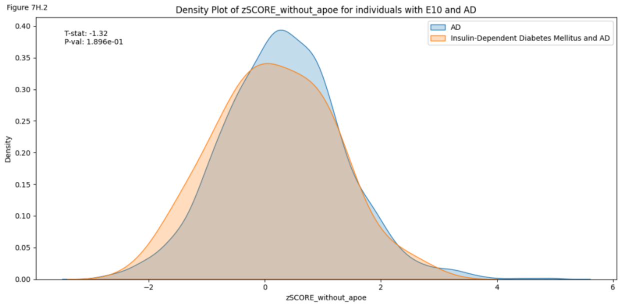


Figure 7I.1

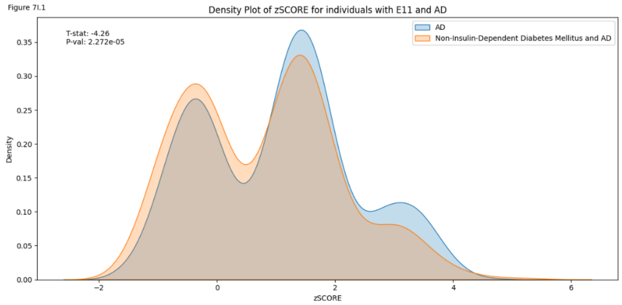


Figure 7I.2

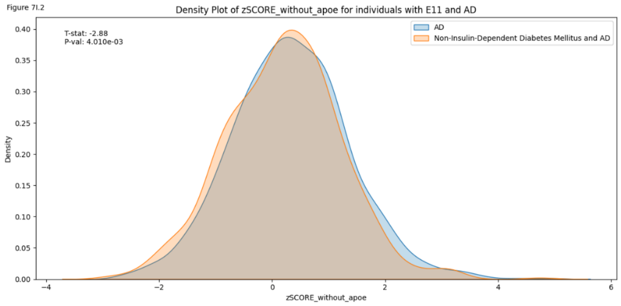


Figure 7J.1

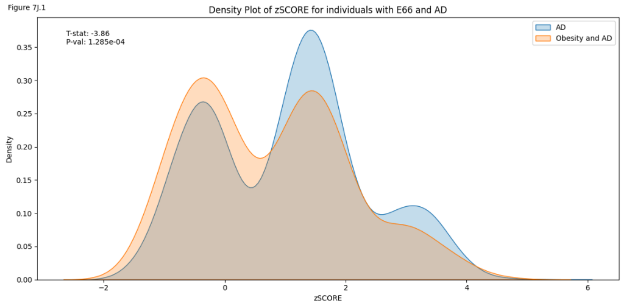


Figure 7J.2

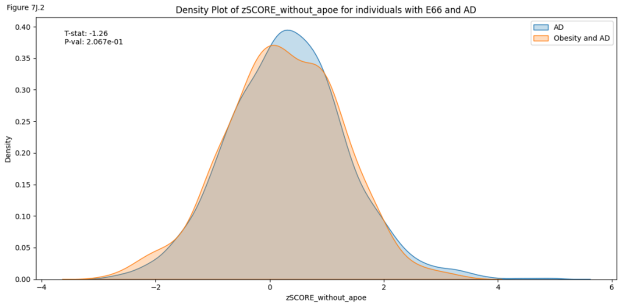

Figure 7K.1

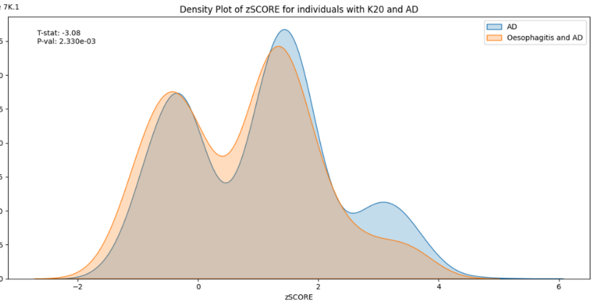


Figure 7K.2

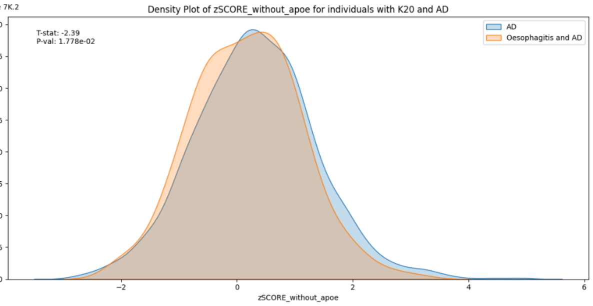


Figure 7L.1

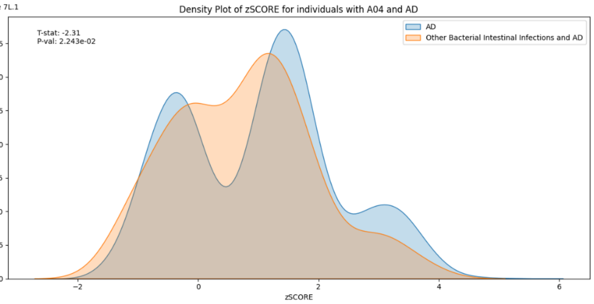


Figure 7L.2

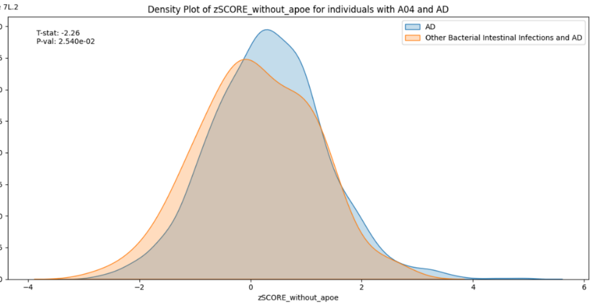


Figure 7M.1

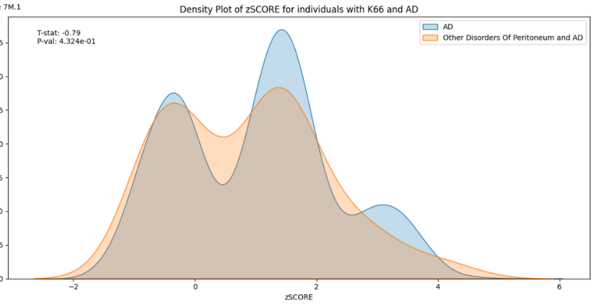


Figure 7M.2

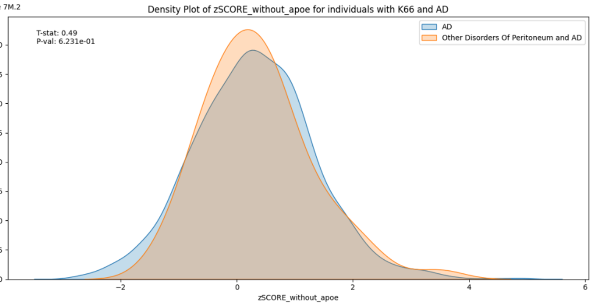


Figure 7N.1

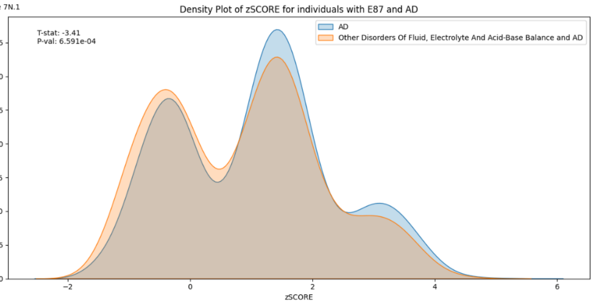


Figure 7N.2

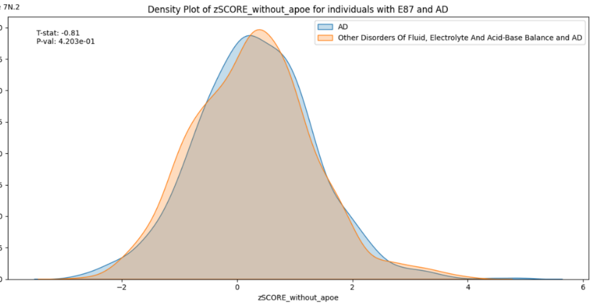


Figure 7O.1

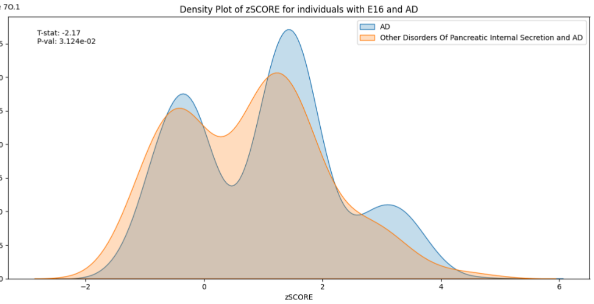


Figure 7O.2

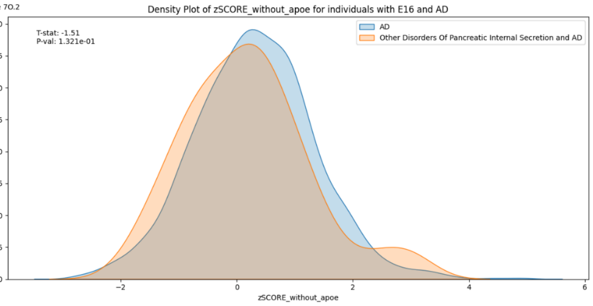

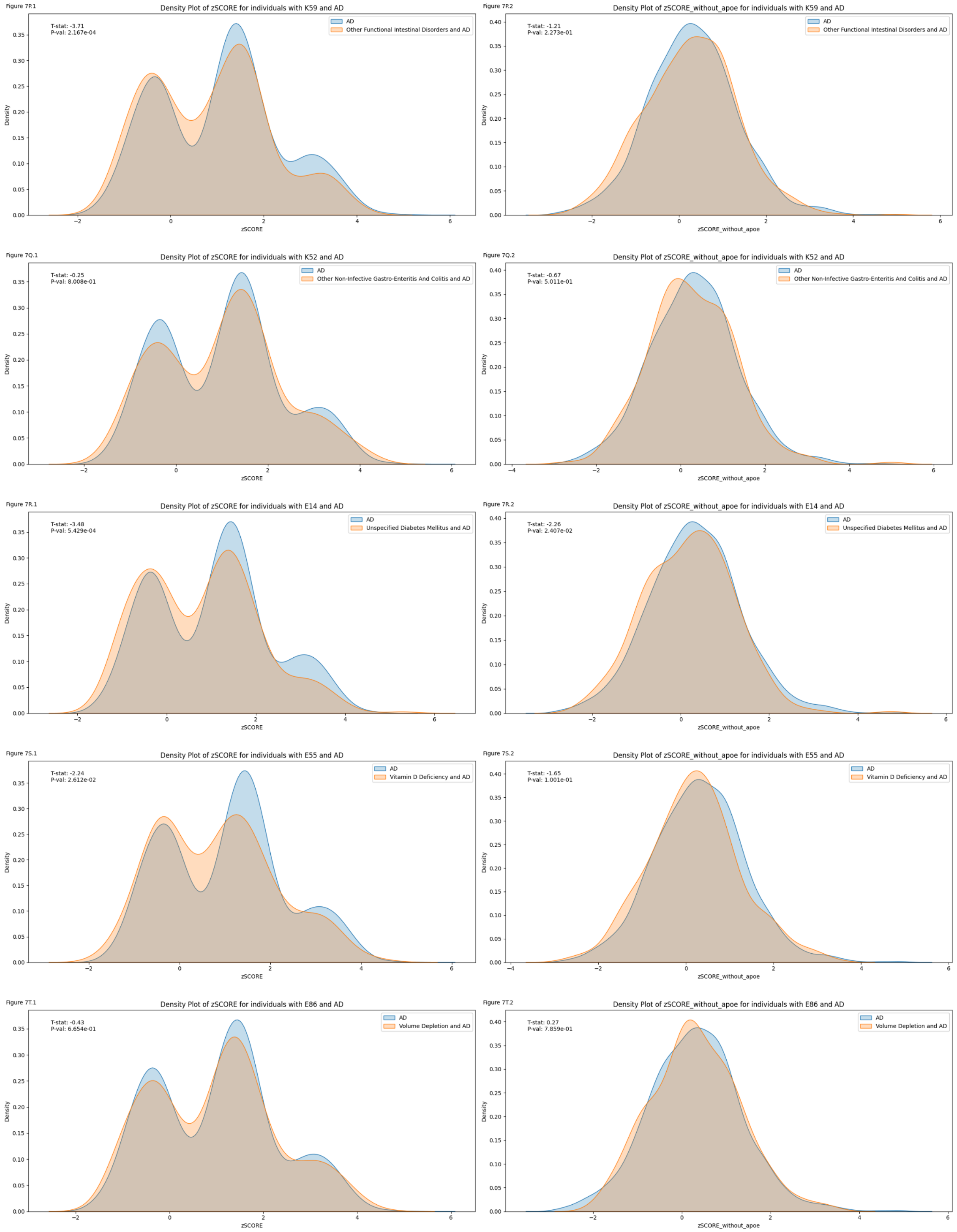


**Fig. S7** Alzheimer's Disease (AD) polygenic risk scores (PRS) with and without *APOE* are differently distributed between cases of AD with and without prior diagnosis of a disorder affecting the gut-brain axis.

Supplementary Figure 8A

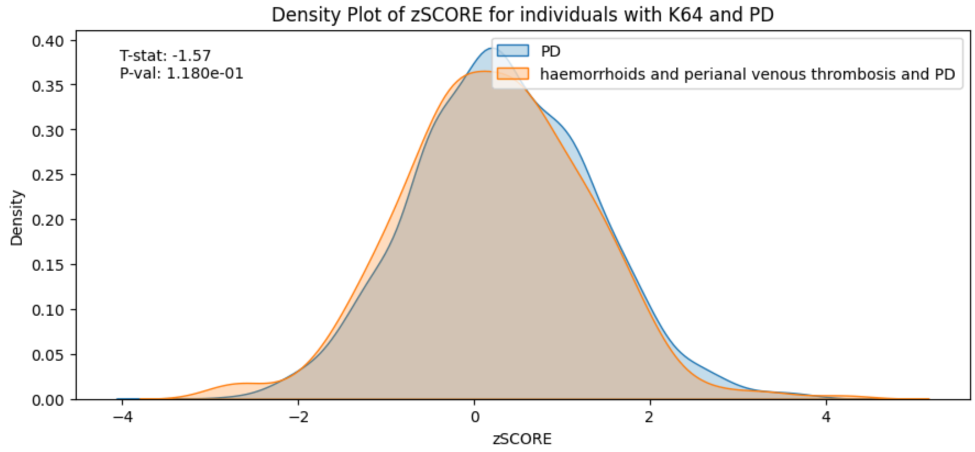


Supplementary Figure 8B

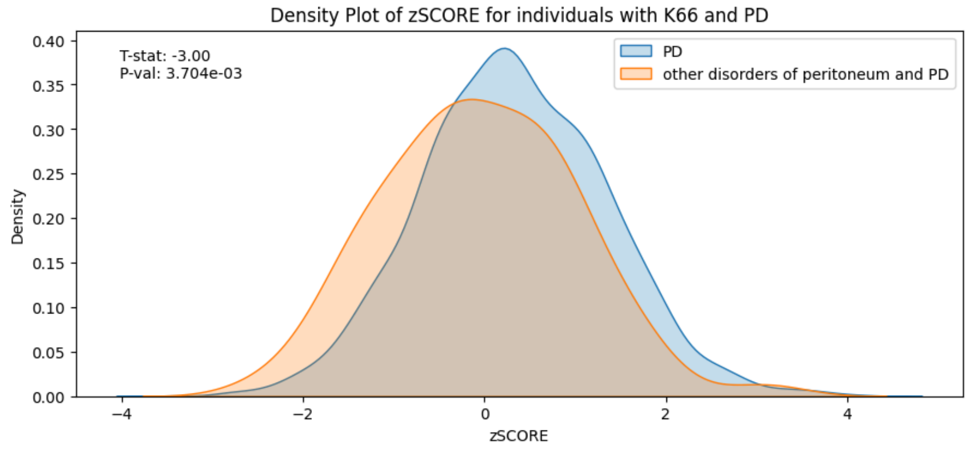

Supplementary Figure 8C

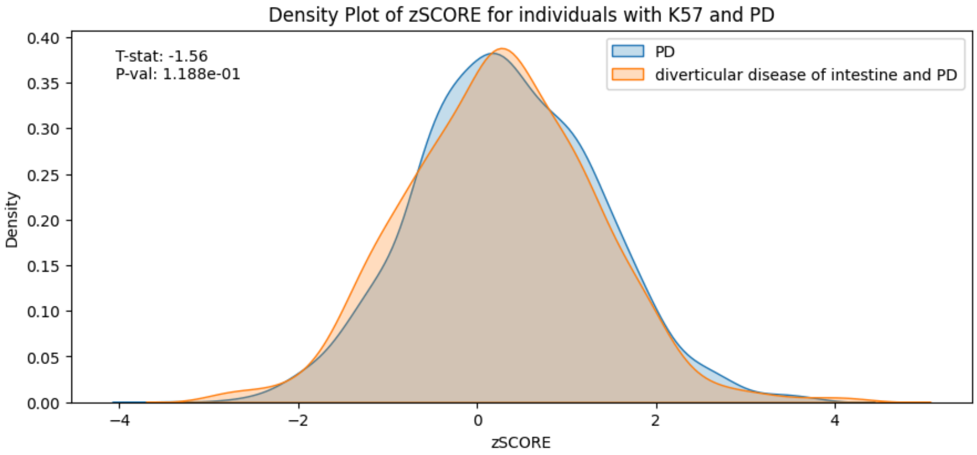


Supplementary Figure 8D

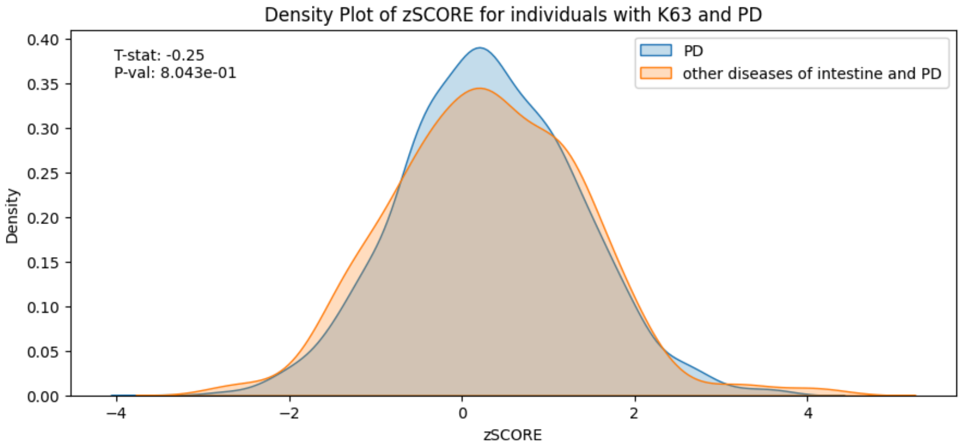


Supplementary Figure 8E

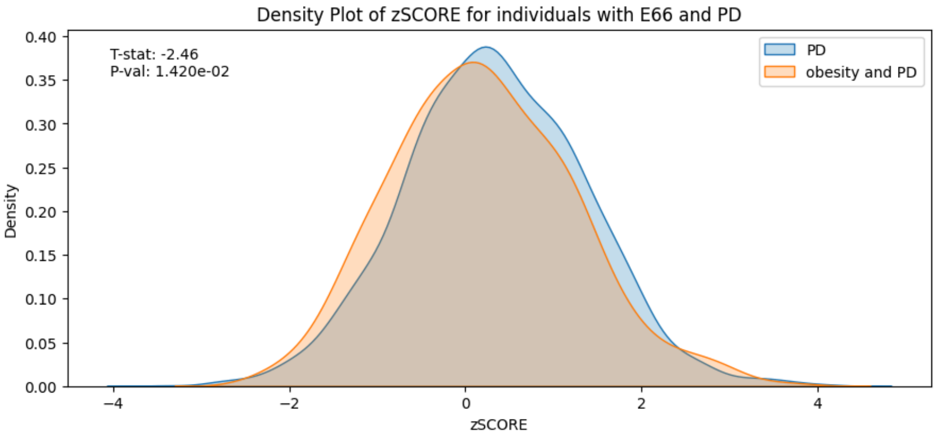

Supplementary Figure 8F

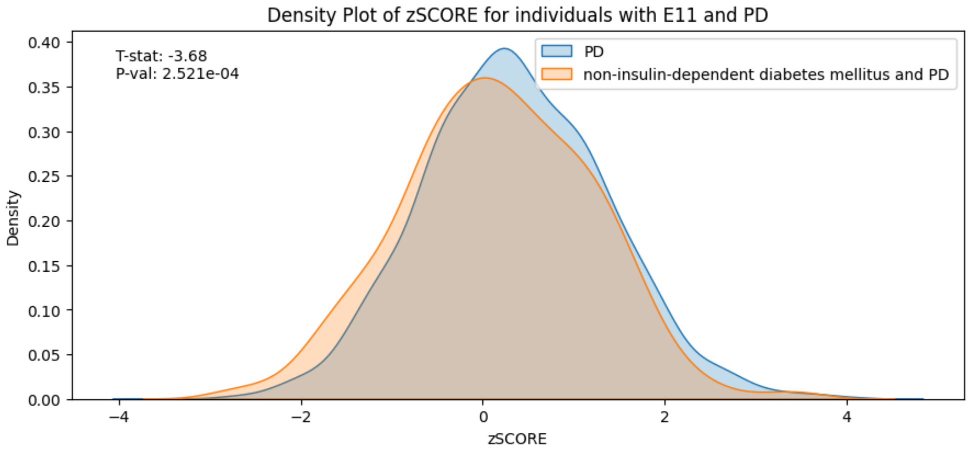


Supplementary Figure 8G

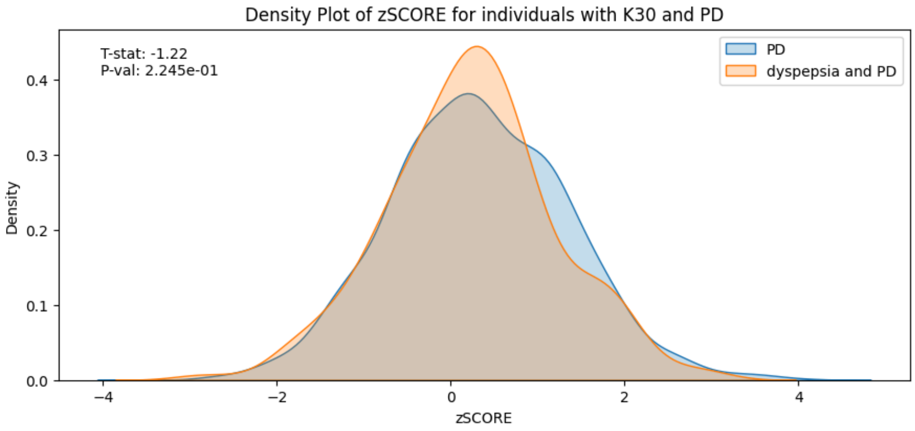


Supplementary Figure 8H

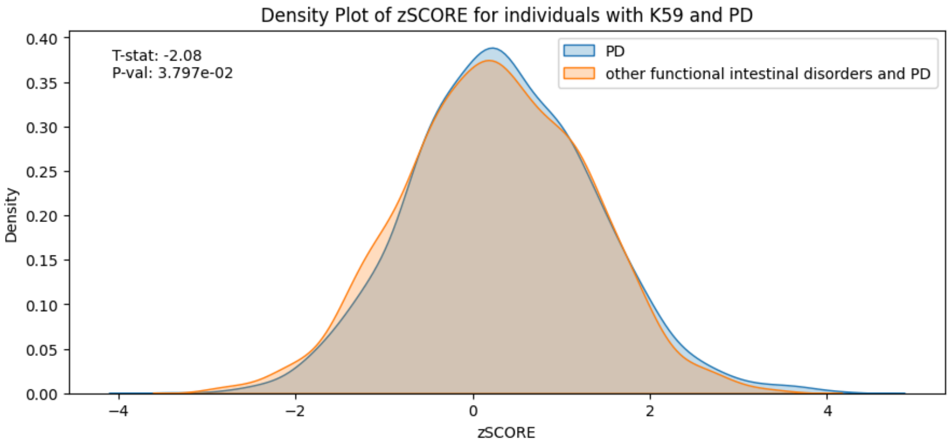

Supplementary Figure 8I

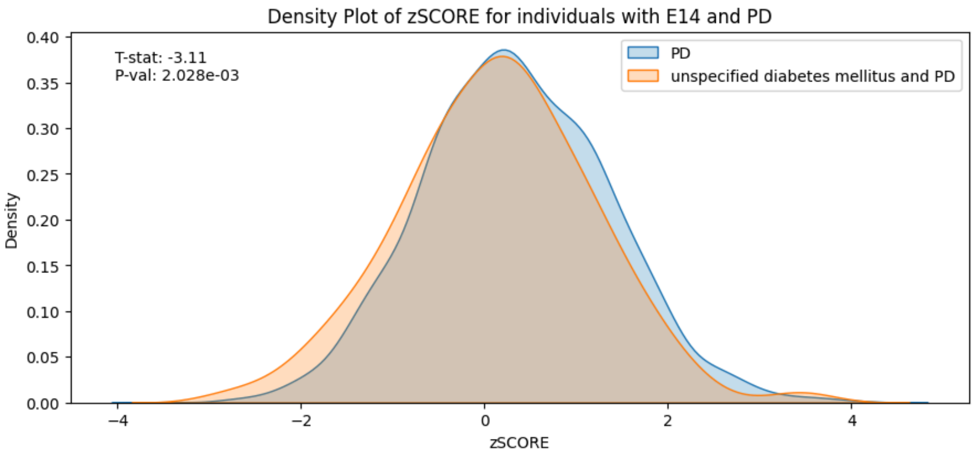


Supplementary Figure 8J

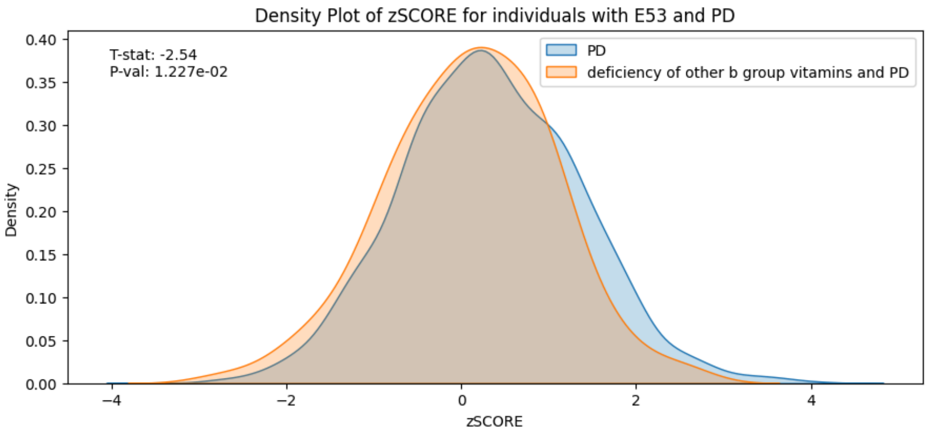


Supplementary Figure 8K

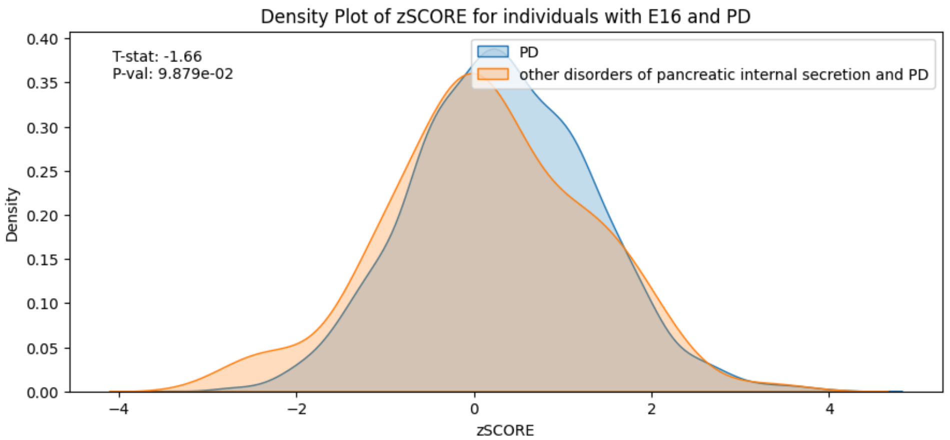

Supplementary Figure 8L

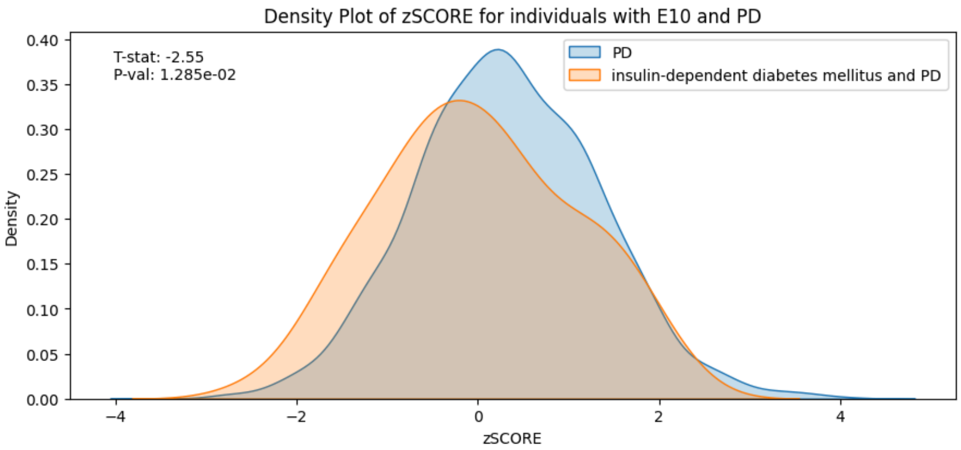


**Fig. S8** Parkinson's Disease (PD) polygenic risk scores (PRS) are differently distributed between cases of PDs with and without prior diagnosis of a disorder affecting the gut-brain axis.
