## Supplemental Tables for "Gut-Brain Nexus: Mapping Multi-Modal Links to Neurodegeneration at Biobank Scale"

| **Overview supplementary tables** |
|---|
| Supplementary Table 1. Cox proportional hazards regression analysis of Alzheimer's disease and endocrine, nutritional, metabolic, and digestive system disorders ICD-10 codes adjusted for year of birth, Townsend deprivation index, and sex |
| Supplementary Table 2. Cox proportional hazards regression analysis of Parkinson's disease and endocrine, nutritional, metabolic, and digestive system disorders ICD-10 codes adjusted for year of birth, Townsend deprivation index, and sex |
| Supplementary Table 3. Interaction terms between endocrine, nutritional, metabolic, digestive system disorders and Alzheimer’s disease polygenic risk score (excluding APOE) |
| Supplementary Table 4. Interaction terms between endocrine, nutritional, metabolic, and digestive system disorders and Alzheimer’s disease polygenic risk score |
| Supplementary Table 5. Interaction terms between endocrine, nutritional, metabolic, digestive system disorders and Parkinson’s disease polygenic risk score |
| Supplementary Table 6. Biomarkers Associated with AD |
| Supplementary Table 7. Biomarkers Associated with PD |
| Supplementary Table 8. Performance summary of different feature sets for AD classification |
| Supplementary Table 9. Performance summary of different feature sets for PD classification |
| Supplementary Table 10. ICD 10 codes used to create control cohort |
| Supplementary Table 11. Biobank cohort gender and Alzheimer’s disease/Parkinson’s disease demographics |
| Supplementary Table 12. UKB diagnosis and disorders under study |
| Supplementary Table 13. Proteomic biomarkers under study |
| Supplementary Table 14. Cox proportional hazards regression analysis of Alzheimer’s disease and endocrine, nutritional, metabolic, and digestive system disorders ICD-10 codes adjusted for APOE4/4 status, principal components 1-5, year of birth, Townsend deprivation index, and sex |
| Supplementary Table 15. Cox proportional hazards regression analysis of Alzheimer's disease and endocrine, nutritional, metabolic, and digestive system disorders ICD-10 codes adjusted for polygenic risk Z-score excluding APOE, APOE4/4 status, principal components 1-5, year of birth, Townsend deprivation index, and sex |
| Supplementary Table 16. Cox proportional hazards regression analysis of Alzheimer's disease and endocrine, nutritional, metabolic, and digestive system disorders ICD-10 codes adjusted for polygenic risk Z-scores excluding APOE, principal components 1-5, year of birth, Townsend deprivation index, and sex |
| Supplementary Table 17. Cox proportional hazards regression analysis of Alzheimer's disease and endocrine, nutritional, metabolic, and digestive system disorders ICD-10 codes adjusted for polygenic risk Z-score, APOE4/4 status, principal components 1-5, year of birth, Townsend deprivation index, and sex |
| Supplementary Table 18. Cox proportional hazards regression analysis of Alzheimer's disease and endocrine, nutritional, metabolic, and digestive system disorders ICD-10 codes adjusted for polygenic risk Z-scores, principal components 1-5, year of birth, Townsend deprivation index, and sex |
| Supplementary Table 19. Cox proportional hazards regression analysis of Alzheimer's disease and endocrine, nutritional, metabolic, and digestive system disorders ICD-10 codes adjusted for principal components 1-5, year of birth, Townsend deprivation index, and sex |
| Supplementary Table 20. Cox proportional hazards regression analysis of Parkinson's disease and endocrine, nutritional, metabolic, and digestive system disorders ICD-10 codes adjusted for principal components 1-5, year of birth, Townsend deprivation index, and sex |
| Supplementary Table 21. Cox proportional hazards regression analysis of Parkinson's disease and endocrine, nutritional, metabolic, and digestive system disorders ICD-10 codes adjusted for polygenic risk Z-scores, principal components 1-5, Townsend deprivation index, and sex |
| Supplementary Table 22. Cox proportional hazards regression analysis of Parkinson's disease and endocrine, nutritional, metabolic, and digestive system disorders ICD-10 codes adjusted for GBA1 (G_1_155162560 + T_1_155235843) status, principal components 1-5, year of birth, Townsend deprivation index, and sex |
| Supplementary Table 23. Cox proportional hazards regression analysis of Parkinson's disease and endocrine, nutritional, metabolic, and digestive system disorders ICD-10 codes adjusted for polygenic risk Z-scores, GBA1 (G_1_155162560 + T_1_155235843) status, principal components 1-5, year of birth, Townsend deprivation index, and sex |
| Supplementary Table 24. Cox proportional hazards regression analysis of Parkinson's disease and endocrine, nutritional, metabolic, and digestive system disorders ICD-10 codes adjusted for LRRK2 (C_12_40220632 + G_12_40340400) status, principal components 1-5, year of birth, Townsend deprivation index, and sex |
| Supplementary Table 25. Cox proportional hazards regression analysis of Parkinson's disease and endocrine, nutritional, metabolic, and digestive system disorders ICD-10 codes adjusted for polygenic risk Z-scores, LRRK2 (C_12_40220632 + G_12_40340400) status, principal components 1-5, year of birth, Townsend deprivation index, and sex |

**Cox proportional hazards regression analysis of Alzheimer's disease and endocrine, nutritional, metabolic, and digestive system disorders ICD-10 codes adjusted for year of birth, Townsend deprivation index, and sex**

| UKB field corresponding to the ICD 10 code | ICD10 code | Definition of ICD10 code | Hazard Ratio | ci_min | ci_max | P_VAL | N_pairs | n | P_VAL_FDR_CORRECTED | rejected |
|---|---|---|---|---|---|---|---|---|---|---|
| p131654 | K66 | other disorders of peritoneum | 0.59 | 0.44 | 0.81 | 8.77E-04 | 41 | 4636 | 5.76E-03 | TRUE |
| p131650 | K64 | haemorrhoids and perianal venous thrombosis | 0.69 | 0.59 | 0.80 | 6.94E-07 | 186 | 20162 | 8.10E-06 | TRUE |
| p130792 | E66 | obesity | 0.85 | 0.74 | 0.96 | 9.29E-03 | 260 | 22619 | 4.43E-02 | TRUE |
| p130814 | E78 | disorders of lipoprotein metabolism and other lipidaemias | 1.18 | 1.09 | 1.27 | 1.53E-05 | 1090 | 53797 | 1.34E-04 | TRUE |
| p131598 | K29 | gastritis and duodenitis | 1.20 | 1.09 | 1.32 | 1.45E-04 | 523 | 27914 | 1.17E-03 | TRUE |
| p131582 | K20 | oesophagitis | 1.28 | 1.10 | 1.49 | 1.63E-03 | 175 | 8660 | 1.00E-02 | TRUE |
| p130820 | E83 | disorders of mineral metabolism | 1.32 | 1.08 | 1.62 | 7.75E-03 | 94 | 4435 | 3.88E-02 | TRUE |
| p131630 | K52 | other non-infective gastro-enteritis and colitis | 1.37 | 1.20 | 1.56 | 1.75E-06 | 250 | 12984 | 1.84E-05 | TRUE |
| p131560 | K04 | diseases of pulp and periapical tissues | 1.44 | 1.11 | 1.87 | 6.21E-03 | 57 | 3365 | 3.26E-02 | TRUE |
| p130708 | E11 | non-insulin-dependent diabetes mellitus | 1.47 | 1.33 | 1.63 | 1.19E-14 | 476 | 19048 | 4.18E-13 | TRUE |
| p130828 | E87 | other disorders of fluid, electrolyte and acid-base balance | 1.51 | 1.34 | 1.70 | 6.43E-12 | 312 | 11313 | 1.35E-10 | TRUE |
| p130008 | A04 | other bacterial intestinal infections | 1.51 | 1.21 | 1.89 | 2.35E-04 | 81 | 3585 | 1.76E-03 | TRUE |
| p130714 | E14 | unspecified diabetes mellitus | 1.53 | 1.33 | 1.75 | 1.21E-09 | 224 | 8733 | 1.58E-08 | TRUE |
| p131640 | K59 | other functional intestinal disorders | 1.55 | 1.40 | 1.73 | 3.55E-16 | 394 | 15596 | 1.86E-14 | TRUE |
| p130770 | E53 | deficiency of other b group vitamins | 1.78 | 1.40 | 2.27 | 2.61E-06 | 68 | 1939 | 2.49E-05 | TRUE |
| p130826 | E86 | volume depletion | 1.78 | 1.52 | 2.10 | 3.26E-12 | 153 | 4911 | 8.57E-11 | TRUE |
| p130774 | E55 | vitamin d deficiency | 1.95 | 1.57 | 2.41 | 9.97E-10 | 87 | 2405 | 1.50E-08 | TRUE |
| p130718 | E16 | other disorders of pancreatic internal secretion | 2.23 | 1.75 | 2.83 | 5.27E-11 | 69 | 1637 | 9.22E-10 | TRUE |
| p130824 | E85 | amyloidosis | 2.57 | 1.34 | 4.94 | 4.72E-03 | 9 | 182 | 2.61E-02 | TRUE |
| p130706 | E10 | insulin-dependent diabetes mellitus | 3.09 | 2.45 | 3.90 | 1.77E-21 | 73 | 1633 | 1.86E-19 | TRUE |

UKB: UK Biobank
AD: Alzheimer's disease
ci_min: Confidence Interval minimum
ci_max: Confidence Interval maximum
P_VAL: p-value
N_pairs: Number of individuals identified with both ICD-10 code and neurodegenerative disease outcome
n: Number of Individulas Identified with ICD10_code
P_VAL_FDR_CORRECTED: p-value after False Discovery Rate corrected
Model: ICD10 + Year_of_birth + Townsend_deprivation_index + sex

**Cox proportional hazards regression analysis of Parkinson's disease and endocrine, nutritional, metabolic, and digestive system disorders ICD-10 codes adjusted for year of birth, Townsend deprivation index, and sex**

| UKB field corresponding to the ICD_10 code | ICD10_code | Definition of ICD10_code | Hazard Ratio | ci_min | ci_max | P_VAL | N_pairs | n | P_VAL_FDR_CORRECTED | rejected |
|---|---|---|---|---|---|---|---|---|---|---|
| p131650 | K64 | haemorrhoids and perianal venous thrombosis | 0.59 | 0.50 | 0.71 | 3.62E-09 | 136 | 20006 | 7.10E-08 | TRUE |
| p131654 | K66 | other disorders of peritoneum | 0.63 | 0.45 | 0.87 | 5.81E-03 | 35 | 4612 | 4.42E-02 | TRUE |
| p131636 | K57 | diverticular disease of intestine | 0.69 | 0.62 | 0.77 | 6.24E-11 | 354 | 34801 | 2.04E-09 | TRUE |
| p131648 | K63 | other diseases of intestine | 0.69 | 0.59 | 0.80 | 1.34E-06 | 183 | 19139 | 2.20E-05 | TRUE |
| p130792 | E66 | obesity | 0.82 | 0.71 | 0.94 | 5.86E-03 | 211 | 22453 | 4.42E-02 | TRUE |
| p130708 | E11 | non-insulin-dependent diabetes mellitus | 1.21 | 1.08 | 1.36 | 1.36E-03 | 330 | 18815 | 1.33E-02 | TRUE |
| p131600 | K30 | dyspepsia | 1.34 | 1.13 | 1.60 | 8.51E-04 | 135 | 8811 | 9.26E-03 | TRUE |
| p131640 | K59 | other functional intestinal disorders | 1.56 | 1.38 | 1.76 | 4.81E-13 | 301 | 15431 | 4.71E-11 | TRUE |
| p130714 | E14 | unspecified diabetes mellitus | 1.61 | 1.39 | 1.86 | 1.81E-10 | 197 | 8658 | 4.44E-09 | TRUE |
| p130770 | E53 | deficiency of other b group vitamins | 1.72 | 1.30 | 2.29 | 1.88E-04 | 48 | 1907 | 2.30E-03 | TRUE |
| p130718 | E16 | other disorders of pancreatic internal secretion | 1.84 | 1.36 | 2.47 | 6.76E-05 | 44 | 1602 | 9.47E-04 | TRUE |
| p130706 | E10 | insulin-dependent diabetes mellitus | 2.65 | 2.02 | 3.48 | 2.44E-12 | 53 | 1608 | 1.20E-10 | TRUE |

UKB: UK Biobank
PD: Parkinson's disease
ci_min: Confidence Interval minimum
ci_max: Confidence Interval maximum
P_VAL: p-value
N_pairs: Number of individuals identified with both ICD-10 code and neurodegenerative disease outcome
n: Number of Individulas Identified with ICD10_code
P_VAL_FDR_CORRECTED: p-value after False Discovery Rate corrected
Model: ICD10 + Year_of_birth + Townsend_deprivation_index + sex

**Interaction terms between endocrine, nutritional, metabolic, digestive system disorders and Alzheimer's disease polygenic risk score (excluding APOE)**

| Interaction term | Odds Ratio | 95% CI low | 95% CI high | Beta | SE | 95% CI low | 95% CI high | z | P-value | N_pairs | n | P_VAL_FDR_CORRECTED | rejected |
|---|---|---|---|---|---|---|---|---|---|---|---|---|---|
| zSCORE_without_apoe:(Other Bacterial Intestinal Infections)p130008 | 0.79 | 0.64 | 0.99 | -0.23 | 0.11 | -0.45 | -0.01 | -2.07 | 3.88E-02 | 81 | 3591 | 2.59E-01 | FALSE |
| zSCORE_without_apoe:(Non-Insulin-Dependent Diabetes Mellitus)p130708 | 0.89 | 0.81 | 0.98 | -0.11 | 0.05 | -0.21 | -0.02 | -2.26 | 2.36E-02 | 476 | 19070 | 2.36E-01 | FALSE |
| zSCORE_without_apoe:(Oesophagitis)p131582 | 0.82 | 0.70 | 0.95 | -0.20 | 0.08 | -0.35 | -0.05 | -2.55 | 1.08E-02 | 175 | 8668 | 2.16E-01 | FALSE |

AD: Alzheimer's disease
CI: Confidence interval
SE: Standard error
N_pairs: Number of individuals identified with both ICD-10 code and neurodegenerative disease outcome
n: Number of Individulas Identified with ICD10_code
P_VAL_FDR_CORRECTED: p-value after False Discovery Rate corrected

**Interaction terms between endocrine, nutritional, metabolic, and digestive system disorders and Alzheimer's disease polygenic risk score**

| Interaction term | Odds Ratio | 95% CI low | 95% CI high | Beta | SE | 95% CI low | 95% CI high | z | P-value | N_pairs | n | P_VAL_FDR_CORRECTED | rejected |
|---|---|---|---|---|---|---|---|---|---|---|---|---|---|
| zSCORE:(Non-Insulin-Dependent Diabetes Mellitus)p130708 | 0.91 | 0.84 | 0.99 | -0.10 | 0.04 | -0.18 | -0.01 | -2.27 | 2.30E-02 | 476 | 19070 | 6.57E-02 | FALSE |
| zSCORE:(Obesity)p130792 | 0.84 | 0.75 | 0.93 | -0.18 | 0.05 | -0.28 | -0.07 | -3.24 | 1.18E-03 | 260 | 22645 | 7.89E-03 | TRUE |
| zSCORE:(Disorders Of Lipoprotein Metabolism And Other Lipidaemias)p130814 | 0.92 | 0.87 | 0.98 | -0.08 | 0.03 | -0.14 | -0.02 | -2.52 | 1.16E-02 | 1090 | 53859 | 4.71E-02 | TRUE |
| zSCORE:(Disorders Of Mineral Metabolism)p130820 | 0.74 | 0.61 | 0.88 | -0.31 | 0.09 | -0.49 | -0.12 | -3.29 | 9.95E-04 | 94 | 4440 | 7.89E-03 | TRUE |
| zSCORE:(Volume Depletion)p130826 | 0.86 | 0.75 | 0.99 | -0.15 | 0.07 | -0.28 | -0.01 | -2.16 | 3.07E-02 | 153 | 4913 | 7.68E-02 | FALSE |
| zSCORE:(Other Disorders Of Fluid, Electrolyte And Acid-Base Balance)p130828 | 0.88 | 0.80 | 0.97 | -0.13 | 0.05 | -0.23 | -0.03 | -2.52 | 1.18E-02 | 312 | 11321 | 4.71E-02 | TRUE |
| zSCORE:(Oesophagitis)p131582 | 0.86 | 0.76 | 0.98 | -0.15 | 0.07 | -0.28 | -0.02 | -2.29 | 2.18E-02 | 175 | 8668 | 6.57E-02 | FALSE |
| zSCORE:(Gastritis And Duodenitis)p131598 | 0.92 | 0.85 | 1.00 | -0.08 | 0.04 | -0.16 | 0.00 | -2.00 | 4.58E-02 | 523 | 27949 | 1.02E-01 | FALSE |
| zSCORE:(Other Functional Intestinal Disorders)p131640 | 0.81 | 0.74 | 0.89 | -0.21 | 0.05 | -0.30 | -0.12 | -4.52 | 6.09E-06 | 395 | 15620 | 1.22E-04 | TRUE |

AD: Alzheimer's disease
CI: Confidence interval
SE: Standard error
N_pairs: Number of individuals identified with both ICD-10 code and neurodegenerative disease outcome
n: Number of Individulas Identified with ICD10_code
P_VAL_FDR_CORRECTED: p-value after False Discovery Rate corrected

**Interaction terms between endocrine, nutritional, metabolic, digestive system disorders and Parkinson's disease polygenic risk score**

| Interaction term | Odds Ratio | 95% CI low | 95% CI high | Beta | SE | 95% CI low | 95% CI high | z | P-value | N_pairs | n | P_VAL_FDR_CORRECTED | rejected |
|---|---|---|---|---|---|---|---|---|---|---|---|---|---|
| zSCORE:(insulin-dependent diabetes mellitus)p130706 | 0.76 | 0.58 | 0.99 | -0.28 | 0.14 | -0.55 | -0.01 | -2.02 | 4.36E-02 | 53 | 1613 | 1.05E-01 | FALSE |
| zSCORE:(non-insulin-dependent diabetes mellitus)p130708 | 0.84 | 0.75 | 0.94 | -0.17 | 0.06 | -0.29 | -0.06 | -2.94 | 3.32E-03 | 333 | 18927 | 1.55E-02 | TRUE |
| zSCORE:(deficiency of other b group vitamins)p130770 | 0.70 | 0.52 | 0.94 | -0.36 | 0.15 | -0.65 | -0.06 | -2.39 | 1.69E-02 | 48 | 1922 | 5.07E-02 | FALSE |
| zSCORE:(other functional intestinal disorders)p131640 | 0.84 | 0.74 | 0.94 | -0.18 | 0.06 | -0.30 | -0.06 | -2.89 | 3.87E-03 | 303 | 15528 | 1.55E-02 | TRUE |
| zSCORE:(other disorders of peritoneum)p131654 | 0.60 | 0.43 | 0.84 | -0.52 | 0.17 | -0.85 | -0.18 | -3.00 | 2.67E-03 | 36 | 4635 | 1.55E-02 | TRUE |

PD: Parkinson's disease
CI: Confidence interval
SE: Standard error
N_pairs: Number of individuals identified with both ICD-10 code and neurodegenerative disease outcome
n: Number of Individulas Identified with ICD10_code
P_VAL_FDR_CORRECTED: p-value after False Discovery Rate corrected

| Biomarkers Associated with AD | | | | | | | | |
|---|---|---|---|---|---|---|---|---|
| Olink_marker | Olink_marker_definition | Odds Ratio | ci_min | ci_max | P_VAL | Bonferroni_Significant | P_VAL_FDR_CORRECTED | rejected |
| tnfsf10 | Tumor necrosis factor ligand superfamily member 10 | 0.56 | 0.41 | 0.77 | 3.37E-04 | FALSE | 2.68E-02 | TRUE |
| nptxr | Neuronal pentraxin receptor | 0.57 | 0.44 | 0.73 | 1.28E-05 | TRUE | 2.67E-03 | TRUE |
| igfbp3 | Insulin-like growth factor-binding protein 3 | 0.60 | 0.46 | 0.77 | 5.86E-05 | FALSE | 7.80E-03 | TRUE |
| hpgds | Hematopoietic prostaglandin D synthase | 0.65 | 0.52 | 0.82 | 2.27E-04 | FALSE | 2.07E-02 | TRUE |
| bcan | Brevican core protein | 0.65 | 0.52 | 0.82 | 2.04E-04 | FALSE | 1.99E-02 | TRUE |
| cst5 | Cystatin-D | 0.67 | 0.57 | 0.78 | 2.08E-07 | TRUE | 7.75E-05 | TRUE |
| adamts8 | A disintegrin and metalloproteinase with thrombospondin motifs 8 | 0.70 | 0.59 | 0.84 | 7.00E-05 | FALSE | 7.88E-03 | TRUE |
| psg1 | Pregnancy-specific beta-1-glycoprotein 1 | 1.19 | 1.11 | 1.29 | 4.94E-06 | TRUE | 1.20E-03 | TRUE |
| adgrg1 | Adhesion G-protein coupled receptor G1 | 1.20 | 1.10 | 1.31 | 6.74E-05 | FALSE | 7.88E-03 | TRUE |
| ren | Renin | 1.22 | 1.11 | 1.34 | 2.33E-05 | TRUE | 3.41E-03 | TRUE |
| timp4 | Metalloproteinase inhibitor 4 | 1.38 | 1.15 | 1.67 | 6.74E-04 | FALSE | 4.49E-02 | TRUE |
| pvr | Poliovirus receptor | 1.50 | 1.22 | 1.85 | 1.51E-04 | FALSE | 1.58E-02 | TRUE |
| calb1 | Calbindin | 1.50 | 1.24 | 1.82 | 2.29E-05 | TRUE | 3.41E-03 | TRUE |
| tcn2 | Transcobalamin-2 | 1.51 | 1.20 | 1.89 | 3.48E-04 | FALSE | 2.68E-02 | TRUE |
| gdf15 | Growth/differentiation factor 15 | 1.52 | 1.30 | 1.78 | 2.12E-07 | TRUE | 7.75E-05 | TRUE |
| il1rl1 | Interleukin-1 receptor-like 1 | 1.52 | 1.28 | 1.82 | 3.56E-06 | TRUE | 1.04E-03 | TRUE |
| ltbp2 | Latent-transforming growth factor beta-binding protein 2 | 1.67 | 1.32 | 2.11 | 1.53E-05 | TRUE | 2.80E-03 | TRUE |
| il1r1 | Interleukin-1 receptor type 1 | 1.87 | 1.31 | 2.66 | 5.13E-04 | FALSE | 3.57E-02 | TRUE |
| igf2r | Cation-independent mannose-6-phosphate receptor | 1.88 | 1.32 | 2.66 | 3.94E-04 | FALSE | 2.88E-02 | TRUE |
| dcn | Decorin | 1.89 | 1.34 | 2.67 | 2.75E-04 | FALSE | 2.36E-02 | TRUE |
| nefl | Neurofilament light polypeptide | 2.50 | 2.12 | 2.95 | 4.39E-28 | TRUE | 3.21E-25 | TRUE |
| gfap | Glial fibrillary acidic protein | 3.03 | 2.60 | 3.53 | 2.16E-45 | TRUE | 3.16E-42 | TRUE |

AD: Alzheimer's disease
ci_min: Confidence interval minimum
ci_max: Confidence interval maximum
P_VAL: p-value
P_VAL_FDR_CORRECTED: p-value after False Discovery Rate corrected
Model: AD ~ biomarker + zSCORE + Age_at_recruitment + Townsend_deprivation_index + sex + p22009_a1 + p22009_a2 + p22009_a3 + p22009_a4 + p22009_a5

| Biomarkers Associated with PD | | | | | | | | |
|---|---|---|---|---|---|---|---|---|
| **Olink_marker** | **Olink_marker_definition** | **Odds Ratio** | **ci_min** | **ci_max** | **P_VAL** | **Bonferroni_Significant** | **P_VAL_FDR_CORRECTED** | **rejected** |
| itgav | Integrin alpha-V | 0.12 | 0.081 | 0.19 | 3.54E-23 | TRUE | 5.19E-20 | TRUE |
| vat1 | Synaptic vesicle membrane protein VAT-1 homolog | 0.37 | 0.252 | 0.54 | 3.06E-07 | TRUE | 2.80E-05 | TRUE |
| egfr | Epidermal growth factor receptor | 0.37 | 0.240 | 0.57 | 6.45E-06 | TRUE | 4.10E-04 | TRUE |
| megf9 | Multiple epidermal growth factor-like domains protein 9 | 0.43 | 0.298 | 0.63 | 1.05E-05 | TRUE | 5.67E-04 | TRUE |
| adgrg2 | Adhesion G-protein coupled receptor G2 | 0.44 | 0.340 | 0.58 | 1.58E-09 | TRUE | 2.56E-07 | TRUE |
| tnxb | Tenascin-X | 0.45 | 0.345 | 0.59 | 1.18E-08 | TRUE | 1.57E-06 | TRUE |
| itgam | Integrin alpha-M | 0.45 | 0.358 | 0.58 | 7.87E-11 | TRUE | 1.64E-08 | TRUE |
| il13ra1 | Interleukin-13 receptor subunit alpha-1 | 0.46 | 0.335 | 0.64 | 3.57E-06 | TRUE | 2.49E-04 | TRUE |
| itgb1 | Integrin beta-1 | 0.47 | 0.332 | 0.67 | 2.63E-05 | TRUE | 1.07E-03 | TRUE |
| cd99 | CD99 antigen | 0.47 | 0.324 | 0.69 | 9.33E-05 | FALSE | 2.90E-03 | TRUE |
| clec10a | C-type lectin domain family 10 member A | 0.48 | 0.390 | 0.59 | 4.72E-12 | TRUE | 2.30E-09 | TRUE |
| itga11 | Integrin alpha-11 | 0.49 | 0.400 | 0.61 | 2.75E-11 | TRUE | 1.01E-08 | TRUE |
| setmar | Histone-lysine N-methyltransferase SETMAR | 0.50 | 0.396 | 0.63 | 9.17E-09 | TRUE | 1.34E-06 | TRUE |
| itgb2 | Integrin beta-2 | 0.50 | 0.388 | 0.66 | 3.29E-07 | TRUE | 2.83E-05 | TRUE |
| hpgds | Hematopoietic prostaglandin D synthase | 0.51 | 0.416 | 0.62 | 5.45E-11 | TRUE | 1.55E-08 | TRUE |
| bag3 | BAG family molecular chaperone regulator 3 | 0.51 | 0.441 | 0.60 | 1.02E-17 | TRUE | 7.45E-15 | TRUE |
| hyou1 | Hypoxia up-regulated protein 1 | 0.53 | 0.387 | 0.73 | 7.55E-05 | FALSE | 2.57E-03 | TRUE |
| f9 | Coagulation factor IX | 0.53 | 0.365 | 0.78 | 1.15E-03 | FALSE | 1.77E-02 | TRUE |
| eps8l2 | Epidermal growth factor receptor kinase substrate 8-like protein 2 | 0.54 | 0.425 | 0.70 | 1.26E-06 | TRUE | 9.19E-05 | TRUE |
| nomo1 | Nodal modulator 1 | 0.54 | 0.401 | 0.74 | 1.01E-04 | FALSE | 3.07E-03 | TRUE |
| cdon | Cell adhesion molecule-related/down-regulated by oncogenes | 0.55 | 0.419 | 0.72 | 1.15E-05 | TRUE | 5.98E-04 | TRUE |
| ezr | Ezrin | 0.55 | 0.406 | 0.74 | 9.30E-05 | FALSE | 2.90E-03 | TRUE |
| cant1 | Soluble calcium-activated nucleotidase 1 | 0.55 | 0.384 | 0.78 | 9.96E-04 | FALSE | 1.57E-02 | TRUE |
| smad5 | Mothers against decapentaplegic homolog 5 | 0.55 | 0.387 | 0.78 | 8.62E-04 | FALSE | 1.48E-02 | TRUE |
| ptprn2 | Receptor-type tyrosine-protein phosphatase N2 | 0.55 | 0.441 | 0.69 | 1.36E-07 | TRUE | 1.53E-05 | TRUE |
| scg2 | Secretogranin-2 | 0.55 | 0.441 | 0.69 | 1.68E-07 | TRUE | 1.76E-05 | TRUE |
| erbb3 | Receptor tyrosine-protein kinase erbB-3 | 0.56 | 0.381 | 0.82 | 2.79E-03 | FALSE | 3.18E-02 | TRUE |
| klk8 | Kallikrein-8 | 0.57 | 0.460 | 0.71 | 2.54E-07 | TRUE | 2.48E-05 | TRUE |
| dctpp1 | dCTP pyrophosphatase 1 | 0.57 | 0.457 | 0.71 | 9.16E-07 | TRUE | 7.10E-05 | TRUE |
| crhbp | Corticotropin-releasing factor-binding protein | 0.57 | 0.445 | 0.74 | 1.96E-05 | TRUE | 8.96E-04 | TRUE |
| boc | Brother of CDO | 0.58 | 0.423 | 0.79 | 6.99E-04 | FALSE | 1.31E-02 | TRUE |
| ifnlr1 | Interferon lambda receptor 1 | 0.59 | 0.473 | 0.74 | 5.47E-06 | TRUE | 3.64E-04 | TRUE |
| tppp3 | Tubulin polymerization-promoting protein family member 3 | 0.60 | 0.476 | 0.75 | 1.05E-05 | TRUE | 5.67E-04 | TRUE |
| rtbdn | Retbindin | 0.60 | 0.466 | 0.77 | 7.80E-05 | FALSE | 2.58E-03 | TRUE |
| angptl3 | Angiopoietin-related protein 3 | 0.60 | 0.491 | 0.74 | 9.22E-07 | TRUE | 7.10E-05 | TRUE |
| tnfsf10 | Tumor necrosis factor ligand superfamily member 10 | 0.60 | 0.457 | 0.80 | 4.58E-04 | FALSE | 1.01E-02 | TRUE |
| ptprf | Receptor-type tyrosine-protein phosphatase F | 0.61 | 0.472 | 0.78 | 7.39E-05 | FALSE | 2.57E-03 | TRUE |
| hnmt | Histamine N-methyltransferase | 0.61 | 0.526 | 0.71 | 6.35E-11 | TRUE | 1.55E-08 | TRUE |
| st6gal1 | Beta-galactoside alpha-2,6-sialyltransferase 1 | 0.62 | 0.483 | 0.79 | 1.05E-04 | FALSE | 3.10E-03 | TRUE |
| erp44 | Endoplasmic reticulum resident protein 44 | 0.62 | 0.464 | 0.82 | 9.54E-04 | FALSE | 1.55E-02 | TRUE |
| cd99l2 | CD99 antigen-like protein 2 | 0.62 | 0.461 | 0.84 | 1.71E-03 | FALSE | 2.32E-02 | TRUE |
| comp | Cartilage oligomeric matrix protein | 0.62 | 0.499 | 0.77 | 2.03E-05 | TRUE | 9.00E-04 | TRUE |
| calb2 | Calretinin | 0.62 | 0.483 | 0.80 | 2.77E-04 | FALSE | 7.16E-03 | TRUE |
| asgr1 | Asialoglycoprotein receptor 1 | 0.62 | 0.495 | 0.79 | 7.09E-05 | FALSE | 2.53E-03 | TRUE |
| furin | Furin | 0.62 | 0.507 | 0.77 | 9.61E-06 | TRUE | 5.62E-04 | TRUE |
| igfbp7 | Insulin-like growth factor-binding protein 7 | 0.63 | 0.494 | 0.81 | 2.80E-04 | FALSE | 7.16E-03 | TRUE |
| robo2 | Roundabout homolog 2 | 0.63 | 0.463 | 0.87 | 4.29E-03 | FALSE | 4.24E-02 | TRUE |
| apom | Apolipoprotein M | 0.63 | 0.496 | 0.81 | 2.75E-04 | FALSE | 7.16E-03 | TRUE |
| erbb2 | Receptor tyrosine-protein kinase erbB-2 | 0.64 | 0.473 | 0.85 | 2.59E-03 | FALSE | 3.08E-02 | TRUE |
| rgmb | Repulsive guidance molecule B | 0.64 | 0.486 | 0.83 | 9.36E-04 | FALSE | 1.54E-02 | TRUE |
| ptprs | Receptor-type tyrosine-protein phosphatase S | 0.64 | 0.477 | 0.85 | 2.23E-03 | FALSE | 2.79E-02 | TRUE |
| efna1 | Ephrin-A1 | 0.64 | 0.493 | 0.83 | 6.92E-04 | FALSE | 1.31E-02 | TRUE |
| colec12 | Collectin-12 | 0.64 | 0.491 | 0.83 | 7.87E-04 | FALSE | 1.37E-02 | TRUE |
| matn2 | Matrilin-2 | 0.64 | 0.503 | 0.82 | 4.72E-04 | FALSE | 1.01E-02 | TRUE |
| col18a1 | Collagen alpha-1(XVIII) chain | 0.64 | 0.478 | 0.87 | 3.91E-03 | FALSE | 3.99E-02 | TRUE |
| tafa5 | Chemokine-like protein TAFA-5 | 0.65 | 0.527 | 0.79 | 2.34E-05 | TRUE | 9.83E-04 | TRUE |
| tgfbr3 | Transforming growth factor beta receptor type 3 | 0.65 | 0.519 | 0.81 | 1.06E-04 | FALSE | 3.10E-03 | TRUE |
| vasn | Vasorin | 0.65 | 0.496 | 0.85 | 1.44E-03 | FALSE | 2.11E-02 | TRUE |
| ggh | Gamma-glutamyl hydrolase | 0.65 | 0.526 | 0.80 | 7.01E-05 | FALSE | 2.53E-03 | TRUE |
| itgb6 | Integrin beta-6 | 0.65 | 0.513 | 0.83 | 4.49E-04 | FALSE | 1.01E-02 | TRUE |
| crtac1 | Cartilage acidic protein 1 | 0.65 | 0.518 | 0.82 | 2.69E-04 | FALSE | 7.16E-03 | TRUE |
| prcp | Lysosomal Pro-X carboxypeptidase | 0.65 | 0.520 | 0.82 | 3.07E-04 | FALSE | 7.50E-03 | TRUE |
| acvrl1 | Serine/threonine-protein kinase receptor R3 | 0.66 | 0.493 | 0.88 | 4.91E-03 | FALSE | 4.60E-02 | TRUE |
| dpp4 | Dipeptidyl peptidase 4 | 0.66 | 0.509 | 0.86 | 2.12E-03 | FALSE | 2.67E-02 | TRUE |
| lgals1 | Galectin-1 | 0.66 | 0.540 | 0.82 | 1.17E-04 | FALSE | 3.28E-03 | TRUE |
| ogfr | Opioid growth factor receptor | 0.67 | 0.530 | 0.84 | 4.62E-04 | FALSE | 1.01E-02 | TRUE |
| pamr1 | Inactive serine protease PAMR1 | 0.67 | 0.517 | 0.86 | 1.93E-03 | FALSE | 2.55E-02 | TRUE |
| fap | Prolyl endopeptidase FAP | 0.67 | 0.518 | 0.87 | 2.72E-03 | FALSE | 3.18E-02 | TRUE |
| il17d | Interleukin-17D | 0.67 | 0.520 | 0.87 | 2.68E-03 | FALSE | 3.16E-02 | TRUE |

| | | | | | | | | |
|---|---|---|---|---|---|---|---|---|
| ntf4 | Neurotrophin-4 | 0.68 | 0.533 | 0.86 | 1.54E-03 | FALSE | 2.15E-02 | TRUE |
| efna4 | Ephrin-A4 | 0.68 | 0.537 | 0.86 | 1.50E-03 | FALSE | 2.15E-02 | TRUE |
| cd93 | Complement component C1q receptor | 0.68 | 0.536 | 0.87 | 2.28E-03 | FALSE | 2.82E-02 | TRUE |
| thop1 | Thimet oligopeptidase | 0.69 | 0.539 | 0.87 | 2.09E-03 | FALSE | 2.67E-02 | TRUE |
| tnfrsf13b | Tumor necrosis factor receptor superfamily member 13B | 0.70 | 0.560 | 0.87 | 1.20E-03 | FALSE | 1.80E-02 | TRUE |
| il6r | Interleukin-6 receptor subunit alpha | 0.70 | 0.566 | 0.86 | 9.36E-04 | FALSE | 1.54E-02 | TRUE |
| itgb5 | Integrin beta-5 | 0.70 | 0.572 | 0.86 | 5.55E-04 | FALSE | 1.10E-02 | TRUE |
| cxcl17 | C-X-C motif chemokine 17 | 0.70 | 0.595 | 0.82 | 1.77E-05 | TRUE | 8.37E-04 | TRUE |
| cdh2 | Cadherin-2 | 0.70 | 0.574 | 0.86 | 5.43E-04 | FALSE | 1.09E-02 | TRUE |
| clec14a | C-type lectin domain family 14 member A | 0.70 | 0.558 | 0.89 | 2.78E-03 | FALSE | 3.18E-02 | TRUE |
| tfpi | Tissue factor pathway inhibitor | 0.71 | 0.554 | 0.90 | 4.65E-03 | FALSE | 4.39E-02 | TRUE |
| slitrk6 | SLIT and NTRK-like protein 6 | 0.71 | 0.570 | 0.88 | 1.54E-03 | FALSE | 2.15E-02 | TRUE |
| mfap5 | Microfibrillar-associated protein 5 | 0.71 | 0.599 | 0.83 | 3.68E-05 | FALSE | 1.42E-03 | TRUE |
| enah | Protein enabled homolog | 0.71 | 0.603 | 0.83 | 2.84E-05 | TRUE | 1.12E-03 | TRUE |
| prss27 | Serine protease 27 | 0.71 | 0.591 | 0.85 | 2.84E-04 | FALSE | 7.16E-03 | TRUE |
| blmh | Bleomycin hydrolase | 0.71 | 0.564 | 0.90 | 4.35E-03 | FALSE | 4.27E-02 | TRUE |
| lama4 | Laminin subunit alpha-4 | 0.71 | 0.575 | 0.89 | 2.56E-03 | FALSE | 3.08E-02 | TRUE |
| art3 | Ecto-ADP-ribosyltransferase 3 | 0.71 | 0.581 | 0.88 | 1.53E-03 | FALSE | 2.15E-02 | TRUE |
| ctsd | Cathepsin D | 0.72 | 0.596 | 0.88 | 1.18E-03 | FALSE | 1.79E-02 | TRUE |
| myoc | Myocilin | 0.73 | 0.617 | 0.85 | 1.19E-04 | FALSE | 3.28E-03 | TRUE |
| il16 | Pro-interleukin-16 | 0.73 | 0.609 | 0.87 | 3.88E-04 | FALSE | 9.15E-03 | TRUE |
| nxph1 | Neurexophilin-1 | 0.73 | 0.595 | 0.89 | 1.82E-03 | FALSE | 2.43E-02 | TRUE |
| sost | Sclerostin | 0.73 | 0.607 | 0.88 | 7.27E-04 | FALSE | 1.34E-02 | TRUE |
| ret | Proto-oncogene tyrosine-protein kinase receptor Ret | 0.73 | 0.607 | 0.88 | 7.48E-04 | FALSE | 1.34E-02 | TRUE |
| ccl23 | C-C motif chemokine 23 | 0.73 | 0.612 | 0.87 | 4.59E-04 | FALSE | 1.01E-02 | TRUE |
| agrn | Agrin | 0.73 | 0.594 | 0.90 | 3.68E-03 | FALSE | 3.82E-02 | TRUE |
| crip2 | Cysteine-rich protein 2 | 0.74 | 0.601 | 0.90 | 3.31E-03 | FALSE | 3.61E-02 | TRUE |
| clec4c | C-type lectin domain family 4 member C | 0.74 | 0.633 | 0.86 | 7.95E-05 | FALSE | 2.58E-03 | TRUE |
| dsg3 | Desmoglein-3 | 0.74 | 0.599 | 0.91 | 5.04E-03 | FALSE | 4.70E-02 | TRUE |
| p4hb | Protein disulfide-isomerase | 0.74 | 0.611 | 0.90 | 2.09E-03 | FALSE | 2.67E-02 | TRUE |
| xg | Glycoprotein Xg | 0.74 | 0.611 | 0.90 | 2.10E-03 | FALSE | 2.67E-02 | TRUE |
| cdhr5 | Cadherin-related family member 5 | 0.74 | 0.623 | 0.88 | 7.70E-04 | FALSE | 1.36E-02 | TRUE |
| vwa1 | von Willebrand factor A domain-containing protein 1 | 0.74 | 0.626 | 0.88 | 7.40E-04 | FALSE | 1.34E-02 | TRUE |
| ism1 | Isthmin-1 | 0.75 | 0.620 | 0.90 | 1.80E-03 | FALSE | 2.41E-02 | TRUE |
| clec4a | C-type lectin domain family 4 member A | 0.75 | 0.610 | 0.91 | 4.27E-03 | FALSE | 4.24E-02 | TRUE |
| ly6d | Lymphocyte antigen 6D | 0.75 | 0.622 | 0.92 | 4.48E-03 | FALSE | 4.31E-02 | TRUE |
| vwc2 | Brorin | 0.76 | 0.631 | 0.91 | 2.77E-03 | FALSE | 3.18E-02 | TRUE |
| fabp9 | Fatty acid-binding protein 9 | 0.77 | 0.672 | 0.88 | 1.17E-04 | FALSE | 3.28E-03 | TRUE |
| pcsk9 | Proprotein convertase subtilisin/kexin type 9 | 0.77 | 0.645 | 0.92 | 4.04E-03 | FALSE | 4.08E-02 | TRUE |
| fst | Follistatin | 0.77 | 0.664 | 0.90 | 8.90E-04 | FALSE | 1.51E-02 | TRUE |
| fcn2 | Ficolin-2 | 0.77 | 0.655 | 0.91 | 2.29E-03 | FALSE | 2.82E-02 | TRUE |
| cd70 | CD70 antigen | 0.78 | 0.655 | 0.92 | 3.23E-03 | FALSE | 3.57E-02 | TRUE |
| ccl27 | C-C motif chemokine 27 | 0.78 | 0.691 | 0.88 | 3.78E-05 | FALSE | 1.42E-03 | TRUE |
| fcrl2 | Fc receptor-like protein 2 | 0.78 | 0.672 | 0.90 | 9.94E-04 | FALSE | 1.57E-02 | TRUE |
| s100a11 | Protein S100-A11 | 0.79 | 0.663 | 0.93 | 5.12E-03 | FALSE | 4.71E-02 | TRUE |
| sftpd | Pulmonary surfactant-associated protein D | 0.79 | 0.708 | 0.88 | 1.31E-05 | TRUE | 6.41E-04 | TRUE |
| fcrl1 | Fc receptor-like protein 1 | 0.79 | 0.673 | 0.92 | 3.24E-03 | FALSE | 3.57E-02 | TRUE |
| spink6 | Serine protease inhibitor Kazal-type 6 | 0.79 | 0.692 | 0.90 | 6.09E-04 | FALSE | 1.18E-02 | TRUE |
| adamts15 | A disintegrin and metalloproteinase with thrombospondin motifs 15 | 0.79 | 0.682 | 0.92 | 2.83E-03 | FALSE | 3.21E-02 | TRUE |
| stc1 | Stanniocalcin-1 | 0.79 | 0.675 | 0.93 | 5.30E-03 | FALSE | 4.82E-02 | TRUE |
| lamp3 | Lysosome-associated membrane glycoprotein 3 | 0.80 | 0.701 | 0.91 | 1.08E-03 | FALSE | 1.68E-02 | TRUE |
| dpy30 | Protein dpy-30 homolog | 0.80 | 0.707 | 0.91 | 4.76E-04 | FALSE | 1.01E-02 | TRUE |
| ldlr | Low-density lipoprotein receptor | 0.80 | 0.698 | 0.92 | 1.62E-03 | FALSE | 2.23E-02 | TRUE |
| sumf2 | Inactive C-alpha-formylglycine-generating enzyme 2 | 0.80 | 0.703 | 0.92 | 1.18E-03 | FALSE | 1.79E-02 | TRUE |
| fabp4 | Fatty acid-binding protein, adipocyte | 0.80 | 0.713 | 0.91 | 3.37E-04 | FALSE | 8.08E-03 | TRUE |
| krt5 | Keratin, type II cytoskeletal 5 | 0.80 | 0.690 | 0.94 | 5.11E-03 | FALSE | 4.71E-02 | TRUE |
| inpp1 | Inositol polyphosphate 1-phosphatase | 0.80 | 0.699 | 0.93 | 2.59E-03 | FALSE | 3.08E-02 | TRUE |
| serpinb8 | Serpin B8 | 0.81 | 0.705 | 0.92 | 1.46E-03 | FALSE | 2.12E-02 | TRUE |
| tnfsf11 | Tumor necrosis factor ligand superfamily member 11 | 0.81 | 0.716 | 0.91 | 5.19E-04 | FALSE | 1.05E-02 | TRUE |
| ctsb | Cathepsin B | 0.81 | 0.704 | 0.93 | 2.35E-03 | FALSE | 2.87E-02 | TRUE |
| aif1 | Allograft inflammatory factor 1 | 0.81 | 0.703 | 0.93 | 3.44E-03 | FALSE | 3.70E-02 | TRUE |
| ccl16 | C-C motif chemokine 16 | 0.81 | 0.713 | 0.92 | 1.71E-03 | FALSE | 2.32E-02 | TRUE |
| slc39a5 | Zinc transporter ZIP5 | 0.81 | 0.706 | 0.94 | 3.81E-03 | FALSE | 3.93E-02 | TRUE |
| il11 | Interleukin-11 | 0.82 | 0.716 | 0.94 | 3.93E-03 | FALSE | 3.99E-02 | TRUE |
| selp | P-selectin | 0.82 | 0.724 | 0.94 | 3.50E-03 | FALSE | 3.71E-02 | TRUE |
| ces3 | Carboxylesterase 3 | 0.83 | 0.738 | 0.92 | 9.03E-04 | FALSE | 1.52E-02 | TRUE |
| fbp1 | Fructose-1,6-bisphosphatase 1 | 0.84 | 0.756 | 0.92 | 4.64E-04 | FALSE | 1.01E-02 | TRUE |
| clc | Galectin-10 | 0.84 | 0.746 | 0.94 | 2.08E-03 | FALSE | 2.67E-02 | TRUE |
| gpr37 | Prosaposin receptor GPR37 | 0.85 | 0.762 | 0.95 | 3.60E-03 | FALSE | 3.76E-02 | TRUE |
| fabp5 | Fatty acid-binding protein 5 | 0.85 | 0.763 | 0.95 | 3.41E-03 | FALSE | 3.70E-02 | TRUE |
| crh | Corticoliberin | 0.87 | 0.795 | 0.95 | 1.28E-03 | FALSE | 1.89E-02 | TRUE |
| tcl1a | T-cell leukemia/lymphoma protein 1A | 0.88 | 0.820 | 0.95 | 7.52E-04 | FALSE | 1.34E-02 | TRUE |

| | | | | | | | | |
|---|---|---|---|---|---|---|---|---|
| pspn | Persephin | 0.90 | 0.830 | 0.97 | 4.46E-03 | FALSE | 4.31E-02 | TRUE |
| ppy | Pancreatic prohormone | 0.90 | 0.840 | 0.97 | 4.20E-03 | FALSE | 4.21E-02 | TRUE |
| mep1b | Meprin A subunit beta | 0.92 | 0.864 | 0.97 | 4.53E-03 | FALSE | 4.33E-02 | TRUE |
| epcam | Epithelial cell adhesion molecule | 1.12 | 1.039 | 1.20 | 2.93E-03 | FALSE | 3.29E-02 | TRUE |
| prdx1 | Peroxiredoxin-1 | 1.12 | 1.033 | 1.21 | 5.51E-03 | FALSE | 4.98E-02 | TRUE |
| lxn | Latexin | 1.16 | 1.050 | 1.28 | 3.54E-03 | FALSE | 3.73E-02 | TRUE |
| padi2 | Protein-arginine deiminase type-2 | 1.21 | 1.085 | 1.34 | 5.07E-04 | FALSE | 1.04E-02 | TRUE |
| il1rl1 | Interleukin-1 receptor-like 1 | 1.27 | 1.080 | 1.48 | 3.48E-03 | FALSE | 3.71E-02 | TRUE |
| mmp13 | Collagenase 3 | 1.39 | 1.106 | 1.74 | 4.65E-03 | FALSE | 4.39E-02 | TRUE |
| dsg2 | Desmoglein-2 | 1.49 | 1.145 | 1.94 | 2.98E-03 | FALSE | 3.32E-02 | TRUE |
| ebi3_il27 | Interleukin-27 | 1.50 | 1.189 | 1.89 | 6.14E-04 | FALSE | 1.18E-02 | TRUE |
| ncam1 | Neural cell adhesion molecule 1 | 1.51 | 1.209 | 1.89 | 2.96E-04 | FALSE | 7.34E-03 | TRUE |
| cd276 | CD276 antigen | 1.58 | 1.278 | 1.95 | 2.35E-05 | TRUE | 9.83E-04 | TRUE |
| mertk | Tyrosine-protein kinase Mer | 1.60 | 1.210 | 2.12 | 9.84E-04 | FALSE | 1.57E-02 | TRUE |
| nefl | Neurofilament light polypeptide | 1.62 | 1.395 | 1.88 | 2.19E-10 | TRUE | 4.01E-08 | TRUE |

PD: Parkinson's disease

ci_min: Confidence interval minimum

ci_max: Confidence interval maximum

P_VAL: p-value

P_VAL_FDR_CORRECTED: p-value after False Discovery Rate corrected

Model: PD ~ biomarker + zSCORE + Age_at_recruitment + Townsend_deprivation_index + sex + p22009_a1 + p22009_a2 + p22009_a3 + p22009_a4 + p22009_a5

**Performance summary of different feature sets for AD classification**

| Feature Set | Number of Features | Train AUC | Test AUC | Train Balanced Accuracy | Test Balanced Accuracy | Sensitivity | Specificity | Train AUC_CI | Test AUC_CI | Train Balanced Accuracy_CI | Test Balanced Accuracy_CI | Sensitivity_CI | Specificity_CI |
|---|---|---|---|---|---|---|---|---|---|---|---|---|---|
| Genetics | 7 | 0.81 | 0.74 | 0.73 | 0.69 | 0.634 | 0.74 | 0.81 ± 0.01 | 0.74 ± 0.04 | 0.73 ± 0.02 | 0.69 ± 0.03 | 0.63 ± 0.11 | 0.74 ± 0.09 |
| Clinical | 17 | 0.66 | 0.61 | 0.64 | 0.60 | 0.506 | 0.69 | 0.66 ± 0.02 | 0.61 ± 0.01 | 0.64 ± 0.01 | 0.60 ± 0.02 | 0.51 ± 0.05 | 0.69 ± 0.05 |
| Olink | 10 | 0.94 | 0.87 | 0.87 | 0.79 | 0.776 | 0.80 | 0.94 ± 0.01 | 0.87 ± 0.02 | 0.87 ± 0.02 | 0.79 ± 0.03 | 0.78 ± 0.07 | 0.80 ± 0.05 |
| Demographics | 3 | 0.85 | 0.82 | 0.78 | 0.75 | 0.780 | 0.72 | 0.85 ± 0.02 | 0.82 ± 0.05 | 0.78 ± 0.02 | 0.75 ± 0.03 | 0.78 ± 0.06 | 0.72 ± 0.11 |
| Combined without Clinical | 20 | 0.94 | 0.89 | 0.87 | 0.82 | 0.820 | 0.82 | 0.94 ± 0.00 | 0.89 ± 0.02 | 0.87 ± 0.01 | 0.82 ± 0.04 | 0.82 ± 0.06 | 0.82 ± 0.04 |
| Combined | 37 | 0.95 | 0.90 | 0.88 | 0.83 | 0.820 | 0.83 | 0.95 ± 0.02 | 0.90 ± 0.02 | 0.88 ± 0.02 | 0.83 ± 0.03 | 0.82 ± 0.06 | 0.83 ± 0.04 |

AD: Alzheimer's Disease

AUC: area under the receiver operating characteristic (ROC) curve

**Performance summary of different feature sets for PD classification**

| Feature Set | Number of Features | Train AUC | Test AUC | Train Balanced Accuracy | Test Balanced Accuracy | Sensitivity | Specificity | Train AUC_CI | Test AUC_CI | Train Balanced Accuracy_CI | Test Balanced Accuracy_CI | Sensitivity_CI | Specificity_CI |
|---|---|---|---|---|---|---|---|---|---|---|---|---|---|
| Genetics | 10 | 0.71 | 0.58 | 0.66 | 0.56 | 0.571 | 0.56 | 0.71 ± 0.14 | 0.58 ± 0.04 | 0.66 ± 0.11 | 0.56 ± 0.02 | 0.57 ± 0.12 | 0.56 ± 0.10 |
| Clinical | 7 | 0.55 | 0.52 | 0.54 | 0.53 | 0.141 | 0.91 | 0.55 ± 0.02 | 0.52 ± 0.03 | 0.54 ± 0.01 | 0.53 ± 0.03 | 0.14 ± 0.04 | 0.91 ± 0.09 |
| Olink | 34 | 0.91 | 0.73 | 0.83 | 0.67 | 0.690 | 0.65 | 0.91 ± 0.05 | 0.73 ± 0.03 | 0.83 ± 0.08 | 0.67 ± 0.03 | 0.69 ± 0.05 | 0.65 ± 0.06 |
| Demographics | 3 | 0.80 | 0.77 | 0.73 | 0.72 | 0.822 | 0.61 | 0.80 ± 0.01 | 0.77 ± 0.03 | 0.73 ± 0.01 | 0.72 ± 0.04 | 0.82 ± 0.10 | 0.61 ± 0.10 |
| Combined without Clinical | 47 | 0.90 | 0.78 | 0.81 | 0.71 | 0.793 | 0.62 | 0.90 ± 0.07 | 0.78 ± 0.03 | 0.81 ± 0.10 | 0.71 ± 0.02 | 0.79 ± 0.03 | 0.62 ± 0.06 |
| Combined | 54 | 0.88 | 0.78 | 0.78 | 0.70 | 0.804 | 0.60 | 0.88 ± 0.03 | 0.78 ± 0.04 | 0.78 ± 0.04 | 0.70 ± 0.02 | 0.80 ± 0.04 | 0.60 ± 0.04 |

PD: Parkinson's Disease

AUC: area under the receiver operating characteristic (ROC) curve

| ICD 10 codes used to create control cohort | |
|---|---|
| **ICD-10 Code** | **Description** |
| G10 | Huntington's Disease |
| G11 | Hereditary Ataxia |
| G12 | Spinal Muscular Atrophy and Related Syndromes |
| G13 | Systemic Atrophies Primarily Affecting Central Nervous System in Diseases Classified Elsewhere |
| G14 | Postpolio Syndrome |
| G20 | Parkinson's Disease |
| G21 | Secondary Parkinsonism |
| G22 | Parkinsonism in Diseases Classified Elsewhere |
| G23 | Other Degenerative Diseases of Basal Ganglia |
| G24 | Dystonia |
| G25 | Other Extrapyramidal and Movement Disorders |
| G30 | Alzheimer's Disease |
| G31 | Other Degenerative Diseases of Nervous System, Not Elsewhere Classified |
| G32 | Other Degenerative Disorders of Nervous System in Diseases Classified Elsewhere |
| G35 | Multiple Sclerosis |
| G36 | Other Acute Disseminated Demyelination |
| G37 | Other Demyelinating Diseases of Central Nervous System |
| G45 | Transient Cerebral Ischaemic Attacks and Related Syndromes |
| G46 | Vascular Syndromes of Brain in Cerebrovascular Diseases |
| G50 | Disorders of Trigeminal Nerve |
| G52 | Disorders of Other Cranial Nerves |
| G53 | Cranial Nerve Disorders in Diseases Classified Elsewhere |
| G54 | Nerve Root and Plexus Disorders |
| G55 | Nerve Root and Plexus Compressions in Diseases Classified Elsewhere |
| G56 | Mononeuropathies of Upper Limb |
| G57 | Mononeuropathies of Lower Limb |
| G58 | Other Mononeuropathies |
| G59 | Mononeuropathy in Diseases Classified Elsewhere |
| G60 | Hereditary and Idiopathic Neuropathy |
| G61 | Inflammatory Polyneuropathy |
| G62 | Other Polyneuropathies |
| G63 | Polyneuropathy in Diseases Classified Elsewhere |
| G64 | Other Disorders of Peripheral Nervous System |
| G70 | Myasthenia Gravis and Other Myoneural Disorders |
| G71 | Primary Disorders of Muscles |
| G72 | Other Myopathies |
| G73 | Disorders of Myoneural Junction and Muscle in Diseases Classified Elsewhere |
| G80 | Infantile Cerebral Palsy |
| G81 | Hemiplegia |
| G82 | Paraplegia and Tetraplegia |
| G83 | Other Paralytic Syndromes |
| G90 | Disorders of Autonomic Nervous System |

| ICD 10 codes used to create control cohort | |
|---|---|
| **ICD-10 Code** | **Description** |
| G91 | Hydrocephalus |
| G92 | Toxic Encephalopathy |
| G93 | Other Disorders of Brain |
| G94 | Other Disorders of Brain in Diseases Classified Elsewhere |
| G96 | Other Disorders of Central Nervous System |
| G97 | Postprocedural Disorders of Nervous System, Not Elsewhere Classified |
| G98 | Other Disorders of Nervous System, Not Elsewhere Classified |
| G99 | Other Disorders of Nervous System in Diseases Classified Elsewhere |

**Biobank cohort gender and Alzheimer’s disease/Parkinson’s disease demographics**

| | UKB | | | SAIL | | | Finngen | | |
|---|---|---|---|---|---|---|---|---|---|
| **Neurodegenerative disease** | **All** | **Female** | **Male** | **All** | **Female** | **Male** | **All** | **Female** | **Male** |
| AD | 3,308 | 1,721 | 1,587 | 38,973 | 24,743 | 14,230 | 15,617 | 6,875 | 8,742 |
| PD | 2,780 | 1,024 | 1,756 | 24,270 | 11,387 | 12,883 | 4,681 | 1,858 | 2,823 |
| controls | 261,814 | 138,666 | 123,148 | 2,106,924 | 1,129,391 | 977,533 | 412,181 | 230,310 | 181,871 |

UKB: UK Biobank
SAIL: Secure Anonymised Information Linkage Databank
AD: Alzheimer's disease
PD: Parkinson's disease

AD for UKB is G30 and F00 ICD10 codes
Finngen summary stat https://r10.risteys.finngen.fi/endpoints/G6_AD_WIDE
https://r10.risteys.finngen.fi/endpoints/G6_PARKINSON
https://www.finngen.fi/en/access_results

| UKB diagnosis and disorders under study | | |
|---|---|---|
| **UKB field corresponding to the ICD_10 code** | **ICD10_code** | **Definition of ICD10_code** |
| p130000 | A00 | Cholera |
| p130002 | A01 | Typhoid And Paratyphoid Fevers |
| p130004 | A02 | Other Salmonella Infections |
| p130006 | A03 | Shigellosis |
| p130008 | A04 | Other Bacterial Intestinal Infections |
| p130010 | A05 | Other Bacterial Foodborne Intoxications |
| p130012 | A06 | Amoebiasis |
| p130014 | A07 | Other Protozoal Intestinal Diseases |
| p130016 | A08 | Viral And Other Specified Intestinal Infections |
| p130018 | A09 | Diarrhoea And Gastro-Enteritis Of Presumed Infectious Origin |
| p131552 | K00 | Disorders Of Tooth Development And Eruption |
| p131554 | K01 | Embedded And Impacted Teeth |
| p131556 | K02 | Dental Caries |
| p131558 | K03 | Other Diseases Of Hard Tissues Of Teeth |
| p131560 | K04 | Diseases Of Pulp And Periapical Tissues |
| p131562 | K05 | Gingivitis And Periodontal Diseases |
| p131564 | K06 | Other Disorders Of Gingiva And Edentulous Alveolar Ridge |
| p131566 | K07 | Dentofacial Anomalies [Including Malocclusion] |
| p131568 | K08 | Other Disorders Of Teeth And Supporting Structures |
| p131570 | K09 | Cysts Of Oral Region, Not Elsewhere Classified |
| p131572 | K10 | Other Diseases Of Jaws |
| p131574 | K11 | Diseases Of Salivary Glands |
| p131576 | K12 | Stomatitis And Related Lesions |
| p131578 | K13 | Other Diseases Of Lip And Oral Mucosa |
| p131580 | K14 | Diseases Of Tongue |
| p131582 | K20 | Oesophagitis |
| p131584 | K21 | Gastro-Oesophageal Reflux Disease |
| p131586 | K22 | Other Diseases Of Oesophagus |
| p131588 | K23 | Disorders Of Oesophagus In Diseases Classified Elsewhere |
| p131590 | K25 | Gastric Ulcer |
| p131592 | K26 | Duodenal Ulcer |
| p131594 | K27 | Peptic Ulcer, Site Unspecified |
| p131596 | K28 | Gastrojejunal Ulcer |
| p131598 | K29 | Gastritis And Duodenitis |
| p131600 | K30 | Dyspepsia |
| p131602 | K31 | Other Diseases Of Stomach And Duodenum |
| p131604 | K35 | Acute Appendicitis |
| p131606 | K36 | Other Appendicitis |
| p131608 | K37 | Unspecified Appendicitis |
| p131610 | K38 | Other Diseases Of Appendix |
| p131612 | K40 | Inguinal Hernia |
| p131614 | K41 | Femoral Hernia |
| p131616 | K42 | Umbilical Hernia |
| p131618 | K43 | Ventral Hernia |
| p131620 | K44 | Diaphragmatic Hernia |
| p131622 | K45 | Other Abdominal Hernia |
| p131624 | K46 | Unspecified Abdominal Hernia |
| p131626 | K50 | Crohn'S Disease [Regional Enteritis] |
| p131628 | K51 | Ulcerative Colitis |
| p131630 | K52 | Other Non-Infective Gastro-Enteritis And Colitis |
| p131632 | K55 | Vascular Disorders Of Intestine |
| p131634 | K56 | Paralytic Ileus And Intestinal Obstruction Without Hernia |
| p131636 | K57 | Diverticular Disease Of Intestine |
| p131638 | K58 | Irritable Bowel Syndrome |
| p131640 | K59 | Other Functional Intestinal Disorders |
| p131642 | K60 | Fissure And Fistula Of Anal And Rectal Regions |
| p131644 | K61 | Abscess Of Anal And Rectal Regions |
| p131646 | K62 | Other Diseases Of Anus And Rectum |
| p131648 | K63 | Other Diseases Of Intestine |
| p131650 | K64 | Haemorrhoids And Perianal Venous Thrombosis |
| p131652 | K65 | Peritonitis |

| UKB diagnosis and disorders under study | | |
|---|---|---|
| **UKB field corresponding to the ICD_10 code** | **ICD10_code** | **Definition of ICD10_code** |
| p131654 | K66 | Other Disorders Of Peritoneum |
| p131656 | K67 | Disorders Of Peritoneum In Infectious Diseases Classified Elsewhere |
| p131658 | K70 | Alcoholic Liver Disease |
| p131660 | K71 | Toxic Liver Disease |
| p131662 | K72 | Hepatic Failure, Not Elsewhere Classified |
| p131664 | K73 | Chronic Hepatitis, Not Elsewhere Classified |
| p131666 | K74 | Fibrosis And Cirrhosis Of Liver |
| p131668 | K75 | Other Inflammatory Liver Diseases |
| p131670 | K76 | Other Diseases Of Liver |
| p131672 | K77 | Liver Disorders In Diseases Classified Elsewhere |
| p131674 | K80 | Cholelithiasis |
| p131676 | K81 | Cholecystitis |
| p131678 | K82 | Other Diseases Of Gallbladder |
| p131680 | K83 | Other Diseases Of Biliary Tract |
| p131682 | K85 | Acute Pancreatitis |
| p131684 | K86 | Other Diseases Of Pancreas |
| p131686 | K87 | Disorders Of Gallbladder, Biliary Tract And Pancreas In Diseases Classified Elsewhere |
| p131688 | K90 | Intestinal Malabsorption |
| p131690 | K91 | Postprocedural Disorders Of Digestive System, Not Elsewhere Classified |
| p131692 | K92 | Other Diseases Of Digestive System |
| p131694 | K93 | Disorders Of Other Digestive Organs In Diseases Classified Elsewhere |
| p130690 | E00 | Congenital Iodine-Deficiency Syndrome |
| p130692 | E01 | Iodine-Deficiency-Related Thyroid Disorders And Allied Conditions |
| p130694 | E02 | Subclinical Iodine-Deficiency Hypothyroidism |
| p130696 | E03 | Other Hypothyroidism |
| p130698 | E04 | Other Non-Toxic Goitre |
| p130700 | E05 | Thyrotoxicosis [Hyperthyroidism] |
| p130702 | E06 | Thyroiditis |
| p130704 | E07 | Other Disorders Of Thyroid |
| p130706 | E10 | Insulin-Dependent Diabetes Mellitus |
| p130708 | E11 | Non-Insulin-Dependent Diabetes Mellitus |
| p130710 | E12 | Malnutrition-Related Diabetes Mellitus |
| p130712 | E13 | Other Specified Diabetes Mellitus |
| p130714 | E14 | Unspecified Diabetes Mellitus |
| p130716 | E15 | Nondiabetic Hypoglycaemic Coma |
| p130718 | E16 | Other Disorders Of Pancreatic Internal Secretion |
| p130720 | E20 | Hypoparathyroidism |
| p130722 | E21 | Hyperparathyroidism And Other Disorders Of Parathyroid Gland |
| p130724 | E22 | Hyperfunction Of Pituitary Gland |
| p130726 | E23 | Hypofunction And Other Disorders Of Pituitary Gland |
| p130728 | E24 | Cushing'S Syndrome |
| p130730 | E25 | Adrenogenital Disorders |
| p130732 | E26 | Hyperaldosteronism |
| p130734 | E27 | Other Disorders Of Adrenal Gland |
| p130736 | E28 | Ovarian Dysfunction |
| p130738 | E29 | Testicular Dysfunction |
| p130740 | E30 | Disorders Of Puberty, Not Elsewhere Classified |
| p130742 | E31 | Polyglandular Dysfunction |
| p130744 | E32 | Diseases Of Thymus |
| p130746 | E34 | Other Endocrine Disorders |
| p130748 | E35 | Disorders Of Endocrine Glands In Diseases Classified Elsewhere |
| p130750 | E40 | Kwashiorkor |
| p130752 | E41 | Nutritional Marasmus |
| p130754 | E42 | Marasmic Kwashiorkor |
| p130756 | E43 | Unspecified Severe Protein-Energy Malnutrition |
| p130758 | E44 | Protein-Energy Malnutrition Of Moderate And Mild Degree |
| p130760 | E45 | Retarded Development Following Protein-Energy Malnutrition |
| p130762 | E46 | Unspecified Protein-Energy Malnutrition |
| p130764 | E50 | Vitamin A Deficiency |
| p130766 | E51 | Thiamine Deficiency |
| p130768 | E52 | Niacin Deficiency [Pellagra] |

| UKB diagnosis and disorders under study | | |
|---|---|---|
| **UKB field corresponding to the ICD_10 code** | **ICD10_code** | **Definition of ICD10_code** |
| p130770 | E53 | Deficiency Of Other B Group Vitamins |
| p130772 | E54 | Ascorbic Acid Deficiency |
| p130774 | E55 | Vitamin D Deficiency |
| p130776 | E56 | Other Vitamin Deficiencies |
| p130778 | E58 | Dietary Calcium Deficiency |
| p130780 | E59 | Dietary Selenium Deficiency |
| p130782 | E60 | Dietary Zinc Deficiency |
| p130784 | E61 | Deficiency Of Other Nutrient Elements |
| p130786 | E63 | Other Nutritional Deficiencies |
| p130788 | E64 | Sequelae Of Malnutrition And Other Nutritional Deficiencies |
| p130790 | E65 | Localised Adiposity |
| p130792 | E66 | Obesity |
| p130794 | E67 | Other Hyperalimentation |
| p130796 | E68 | Sequelae Of Hyperalimentation |
| p130798 | E70 | Disorders Of Aromatic Amino-Acid Metabolism |
| p130800 | E71 | Disorders Of Branched-Chain Amino-Acid Metabolism And Fatty-Acid Metabolism |
| p130802 | E72 | Other Disorders Of Amino-Acid Metabolism |
| p130804 | E73 | Lactose Intolerance |
| p130806 | E74 | Other Disorders Of Carbohydrate Metabolism |
| p130808 | E75 | Disorders Of Sphingolipid Metabolism And Other Lipid Storage Disorders |
| p130810 | E76 | Disorders Of Glycosaminoglycan Metabolism |
| p130812 | E77 | Disorders Of Glycoprotein Metabolism |
| p130814 | E78 | Disorders Of Lipoprotein Metabolism And Other Lipidaemias |
| p130816 | E79 | Disorders Of Purine And Pyrimidine Metabolism |
| p130818 | E80 | Disorders Of Porphyrin And Bilirubin Metabolism |
| p130820 | E83 | Disorders Of Mineral Metabolism |
| p130822 | E84 | Cystic Fibrosis |
| p130824 | E85 | Amyloidosis |
| p130826 | E86 | Volume Depletion |
| p130828 | E87 | Other Disorders Of Fluid, Electrolyte And Acid-Base Balance |
| p130830 | E88 | Other Metabolic Disorders |
| p130832 | E89 | Postprocedural Endocrine And Metabolic Disorders, Not Elsewhere Classified |
| p130834 | E90 | Nutritional And Metabolic Disorders In Diseases Classified Elsewhere |
| UKB: UK Biobank | | |

| Proteomic biomarkers under study |
|---|
| **Olink proteomic targets** |
| aarsd1 |
| abhd14b |
| abl1 |
| acaa1 |
| acan |
| ace2 |
| acox1 |
| acp5 |
| acp6 |
| acta2 |
| actn4 |
| acvrl1 |
| acy1 |
| ada |
| ada2 |
| adam15 |
| adam22 |
| adam23 |
| adam8 |
| adamts13 |
| adamts15 |
| adamts16 |
| adamts8 |
| adcyap1r1 |
| adgrb3 |
| adgre2 |
| adgre5 |
| adgrg1 |
| adgrg2 |
| adh4 |
| adm |
| afp |
| ager |
| agr2 |
| agr3 |
| agrn |
| agrp |
| agxt |
| ahcy |
| ahsp |
| aif1 |
| aifm1 |

| Proteomic biomarkers under study |
| --- |
| **Olink proteomic targets** |
| ak1 |
| akr1b1 |
| akr1c4 |
| akt1s1 |
| akt3 |
| alcam |
| aldh1a1 |
| aldh3a1 |
| alpp |
| ambn |
| ambp |
| amfr |
| amigo2 |
| amn |
| amy2a |
| amy2b |
| ang |
| angpt1 |
| angpt2 |
| angptl1 |
| angptl2 |
| angptl3 |
| angptl4 |
| angptl7 |
| ankrd54 |
| anpep |
| anxa10 |
| anxa11 |
| anxa3 |
| anxa4 |
| anxa5 |
| aoc1 |
| aoc3 |
| apbb1ip |
| apex1 |
| aplp1 |
| apoh |
| apom |
| app |
| aprt |
| areg |
| arg1 |

| Proteomic biomarkers under study |
|---|
| **Olink proteomic targets** |
| arhgap1 |
| arhgap25 |
| arhgef12 |
| arid4b |
| arnt |
| arsa |
| arsb |
| art3 |
| artn |
| asah2 |
| asgr1 |
| atf2 |
| atg4a |
| atox1 |
| atp5if1 |
| atp5po |
| atp6ap2 |
| atp6v1d |
| atp6v1f |
| atxn10 |
| axin1 |
| axl |
| azu1 |
| b4galt1 |
| b4gat1 |
| bach1 |
| bag3 |
| bag6 |
| baiap2 |
| bambi |
| bank1 |
| bax |
| bcam |
| bcan |
| bcl2l11 |
| bcr |
| bgn |
| bid |
| bin2 |
| birc2 |
| blmh |
| blvrb |

| Proteomic biomarkers under study |
|---|
| **Olink proteomic targets** |
| bmp4 |
| bmp6 |
| boc |
| bpifb1 |
| brk1 |
| bsg |
| bst1 |
| bst2 |
| btc |
| btn2a1 |
| btn3a2 |
| c19orf12 |
| c1qa |
| c1qtnf1 |
| c2 |
| c2cd2l |
| c4bpb |
| ca1 |
| ca11 |
| ca12 |
| ca13 |
| ca14 |
| ca2 |
| ca3 |
| ca4 |
| ca5a |
| ca6 |
| ca9 |
| calb1 |
| calb2 |
| calca |
| calcoco1 |
| camkk1 |
| cant1 |
| capg |
| carhsp1 |
| casp1 |
| casp10 |
| casp2 |
| casp3 |
| casp8 |
| cblif |

| Proteomic biomarkers under study |
|---|
| **Olink proteomic targets** |
| cbln4 |
| cc2d1a |
| ccdc80 |
| ccl11 |
| ccl13 |
| ccl14 |
| ccl15 |
| ccl16 |
| ccl17 |
| ccl18 |
| ccl19 |
| ccl2 |
| ccl20 |
| ccl21 |
| ccl22 |
| ccl23 |
| ccl24 |
| ccl25 |
| ccl26 |
| ccl27 |
| ccl28 |
| ccl3 |
| ccl4 |
| ccl5 |
| ccl7 |
| ccl8 |
| ccn1 |
| ccn2 |
| ccn3 |
| ccn4 |
| ccn5 |
| ccs |
| cct5 |
| cd109 |
| cd14 |
| cd160 |
| cd163 |
| cd164 |
| cd177 |
| cd1c |
| cd200 |
| cd200r1 |

| Proteomic biomarkers under study |
|---|
| **Olink proteomic targets** |
| cd207 |
| cd209 |
| cd22 |
| cd244 |
| cd27 |
| cd274 |
| cd276 |
| cd28 |
| cd2ap |
| cd300c |
| cd300e |
| cd300lf |
| cd300lg |
| cd302 |
| cd33 |
| cd34 |
| cd38 |
| cd4 |
| cd40 |
| cd40lg |
| cd46 |
| cd48 |
| cd5 |
| cd55 |
| cd58 |
| cd59 |
| cd6 |
| cd63 |
| cd69 |
| cd70 |
| cd74 |
| cd79b |
| cd83 |
| cd84 |
| cd8a |
| cd93 |
| cd99 |
| cd99l2 |
| cdc27 |
| cdc37 |
| cdcp1 |
| cdh1 |

| Proteomic biomarkers under study |
|---|
| **Olink proteomic targets** |
| cdh15 |
| cdh17 |
| cdh2 |
| cdh3 |
| cdh5 |
| cdh6 |
| cdhr1 |
| cdhr2 |
| cdhr5 |
| cdkn1a |
| cdkn2d |
| cdnf |
| cdon |
| cdsn |
| ceacam1 |
| ceacam21 |
| ceacam3 |
| ceacam5 |
| ceacam8 |
| cebpb |
| cela3a |
| cep164 |
| cep20 |
| cep43 |
| cep85 |
| cert |
| ces1 |
| ces2 |
| ces3 |
| cetn2 |
| cfc1 |
| cga |
| cgref1 |
| chac2 |
| chek2 |
| chgb |
| chi3l1 |
| chit1 |
| chl1 |
| chmp1a |
| chrdl1 |
| chrdl2 |

| Proteomic biomarkers under study |
|---|
| **Olink proteomic targets** |
| ciapin1 |
| ckap4 |
| ckmt1a_ckmt1b |
| clc |
| clec10a |
| clec11a |
| clec14a |
| clec1a |
| clec1b |
| clec4a |
| clec4c |
| clec4d |
| clec4g |
| clec5a |
| clec6a |
| clec7a |
| clip2 |
| clmp |
| clpp |
| clps |
| clspn |
| clstn1 |
| clstn2 |
| clta |
| clul1 |
| cndp1 |
| cnpy2 |
| cnpy4 |
| cnst |
| cntn1 |
| cntn2 |
| cntn3 |
| cntn4 |
| cntn5 |
| cntnap2 |
| col18a1 |
| col1a1 |
| col4a1 |
| col6a3 |
| col9a1 |
| colec12 |
| comp |

| Proteomic biomarkers under study |
|---|
| **Olink proteomic targets** |
| comt |
| cope |
| coro1a |
| cox5b |
| cpa1 |
| cpa2 |
| cpb1 |
| cpe |
| cpm |
| cpped1 |
| cpvl |
| cpxm1 |
| cr2 |
| cracr2a |
| cradd |
| creg1 |
| creld2 |
| crh |
| crhbp |
| crhr1 |
| crim1 |
| crip2 |
| crisp2 |
| crkl |
| crlf1 |
| crnn |
| crtac1 |
| crtam |
| crx |
| csf1 |
| csf2ra |
| csf3 |
| cst3 |
| cst5 |
| cst6 |
| cst7 |
| cstb |
| ctf1 |
| ctrb1 |
| ctrc |
| ctsb |
| ctsc |

| Proteomic biomarkers under study |
|---|
| **Olink proteomic targets** |
| ctsd |
| ctsf |
| ctsh |
| ctsl |
| ctso |
| ctss |
| ctsv |
| ctsz |
| cx3cl1 |
| cxadr |
| cxcl1 |
| cxcl10 |
| cxcl11 |
| cxcl12 |
| cxcl13 |
| cxcl14 |
| cxcl16 |
| cxcl17 |
| cxcl3 |
| cxcl5 |
| cxcl6 |
| cxcl8 |
| cxcl9 |
| dab2 |
| dag1 |
| dapp1 |
| dars1 |
| dbi |
| dbnl |
| dcbld2 |
| dcn |
| dctn1 |
| dctn2 |
| dctn6 |
| dctpp1 |
| dcxr |
| ddah1 |
| ddc |
| ddr1 |
| ddx58 |
| decr1 |
| defa1_defa1b |

| Proteomic biomarkers under study |
|---|
| **Olink proteomic targets** |
| defb4a_defb4b |
| dffa |
| dgkz |
| diablo |
| dkk1 |
| dkk3 |
| dkk4 |
| dkkl1 |
| dlk1 |
| dll1 |
| dnaja2 |
| dnajb1 |
| dnajb8 |
| dner |
| dnmbp |
| dnph1 |
| dok2 |
| dpep1 |
| dpep2 |
| dpp10 |
| dpp4 |
| dpp6 |
| dpp7 |
| dpt |
| dpy30 |
| draxin |
| drg2 |
| dsc2 |
| dsg2 |
| dsg3 |
| dsg4 |
| dtx3 |
| duox2 |
| dusp3 |
| ebag9 |
| ebi3_il27 |
| ece1 |
| eda2r |
| edar |
| edil3 |
| efemp1 |
| efna1 |

| Proteomic biomarkers under study |
|---|
| **Olink proteomic targets** |
| efna4 |
| egf |
| egfl7 |
| egfr |
| egln1 |
| eif4b |
| eif4ebp1 |
| eif4g1 |
| eif5a |
| eloa |
| enah |
| eng |
| eno1 |
| eno2 |
| enpp2 |
| enpp5 |
| enpp7 |
| entpd2 |
| entpd5 |
| entpd6 |
| epcam |
| epha1 |
| epha10 |
| epha2 |
| ephb4 |
| ephb6 |
| ephx2 |
| epo |
| eps8l2 |
| erbb2 |
| erbb3 |
| erbb4 |
| erbin |
| ereg |
| erp44 |
| esam |
| esm1 |
| ezr |
| f11r |
| f2r |
| f3 |
| f7 |

| Proteomic biomarkers under study |
|---|
| **Olink proteomic targets** |
| f9 |
| fabp1 |
| fabp2 |
| fabp4 |
| fabp5 |
| fabp6 |
| fabp9 |
| fadd |
| fam3b |
| fam3c |
| fap |
| fas |
| faslg |
| fbp1 |
| fcar |
| fcer2 |
| fcgr2a |
| fcgr2b |
| fcgr3b |
| fcn2 |
| fcrl1 |
| fcrl2 |
| fcrl3 |
| fcrl5 |
| fcrl6 |
| fcrlb |
| fen1 |
| fes |
| fetub |
| fgf19 |
| fgf2 |
| fgf21 |
| fgf23 |
| fgf5 |
| fgfbp1 |
| fgfr2 |
| fgr |
| fhit |
| fis1 |
| fkbp1b |
| fkbp4 |
| fkbp5 |

| Proteomic biomarkers under study |
|---|
| **Olink proteomic targets** |
| fkbp7 |
| fli1 |
| flrt2 |
| flt1 |
| flt3 |
| flt3lg |
| flt4 |
| fmnl1 |
| fmr1 |
| folr1 |
| folr2 |
| folr3 |
| fosb |
| foxo1 |
| foxo3 |
| frzb |
| fst |
| fstl3 |
| fuca1 |
| furin |
| fus |
| fut3_fut5 |
| fut8 |
| fxn |
| fxyd5 |
| fyb1 |
| gal |
| galnt10 |
| galnt2 |
| galnt3 |
| galnt7 |
| gas6 |
| gbp2 |
| gbp4 |
| gcg |
| gcnt1 |
| gdf15 |
| gdf2 |
| gdnf |
| gfap |
| gfer |
| gfod2 |

| **Proteomic biomarkers under study** |
|---|
| **Olink proteomic targets** |
| gfra1 |
| gfra2 |
| gfra3 |
| gga1 |
| ggh |
| ggt1 |
| ggt5 |
| gh1 |
| gh2 |
| ghrhr |
| ghrl |
| gkn1 |
| glb1 |
| glo1 |
| glod4 |
| glrx |
| glt8d2 |
| gmpr |
| gne |
| gnly |
| golm2 |
| gopc |
| gp1ba |
| gp2 |
| gp6 |
| gpa33 |
| gpc1 |
| gpc5 |
| gpkow |
| gpnmb |
| gpr37 |
| grap2 |
| grk5 |
| grn |
| grpel1 |
| gsap |
| gsta1 |
| gsta3 |
| gstp1 |
| guca2a |
| gusb |
| gys1 |

| Proteomic biomarkers under study |
| --- |
| **Olink proteomic targets** |
| gzma |
| gzmb |
| gzmh |
| hagh |
| hao1 |
| hars1 |
| havcr1 |
| havcr2 |
| hbegf |
| hbq1 |
| hcls1 |
| hdgf |
| hebp1 |
| hexim1 |
| hgf |
| hgs |
| hk2 |
| hla_dra |
| hla_e |
| hmbs |
| hmox1 |
| hmox2 |
| hnmt |
| hnrnpk |
| hpcal1 |
| hpgds |
| hs3st3b1 |
| hs6st1 |
| hsd11b1 |
| hsp90b1 |
| hspa1a |
| hspb1 |
| hspb6 |
| hspg2 |
| htra2 |
| hyal1 |
| hyou1 |
| ica1 |
| icam1 |
| icam2 |
| icam3 |
| icam4 |

| Proteomic biomarkers under study |
|---|
| **Olink proteomic targets** |
| icam5 |
| icoslg |
| idi2 |
| ids |
| idua |
| ifng |
| ifngr1 |
| ifngr2 |
| ifnl1 |
| ifnlr1 |
| igf1r |
| igf2r |
| igfbp1 |
| igfbp2 |
| igfbp3 |
| igfbp4 |
| igfbp6 |
| igfbp7 |
| igfbpl1 |
| igsf3 |
| igsf8 |
| ikbkg |
| ikzf2 |
| il10 |
| il10ra |
| il10rb |
| il11 |
| il12a_il12b |
| il12b |
| il12rb1 |
| il13 |
| il13ra1 |
| il15 |
| il15ra |
| il16 |
| il17a |
| il17c |
| il17d |
| il17f |
| il17ra |
| il17rb |
| il18 |

| Proteomic biomarkers under study |
|---|
| **Olink proteomic targets** |
| il18bp |
| il18r1 |
| il18rap |
| il19 |
| il1a |
| il1b |
| il1r1 |
| il1r2 |
| il1rap |
| il1rl1 |
| il1rl2 |
| il1rn |
| il2 |
| il20 |
| il20ra |
| il22ra1 |
| il24 |
| il2ra |
| il2rb |
| il32 |
| il33 |
| il34 |
| il3ra |
| il4 |
| il4r |
| il5 |
| il5ra |
| il6 |
| il6r |
| il6st |
| il7 |
| il7r |
| ilkap |
| impa1 |
| ing1 |
| inhbc |
| inpp1 |
| inppl1 |
| ipcef1 |
| iqgap2 |
| irag2 |
| irak1 |

| Proteomic biomarkers under study |
|---|
| **Olink proteomic targets** |
| irak4 |
| islr2 |
| ism1 |
| itga11 |
| itga5 |
| itga6 |
| itgam |
| itgav |
| itgb1 |
| itgb1bp1 |
| itgb1bp2 |
| itgb2 |
| itgb5 |
| itgb6 |
| itgb7 |
| itih3 |
| itm2a |
| ivd |
| jam2 |
| jchain |
| jun |
| kazald1 |
| kcnip4 |
| kdr |
| kel |
| kifbp |
| kir2dl3 |
| kir3dl1 |
| kirrel2 |
| kit |
| kitlg |
| klb |
| klk1 |
| klk10 |
| klk11 |
| klk12 |
| klk13 |
| klk14 |
| klk4 |
| klk6 |
| klk8 |
| klrb1 |

| Proteomic biomarkers under study |
|---|
| **Olink proteomic targets** |
| klrd1 |
| krt14 |
| krt18 |
| krt19 |
| krt5 |
| kyat1 |
| kynu |
| l1cam |
| lactb2 |
| lag3 |
| lair1 |
| lair2 |
| lama4 |
| lamp2 |
| lamp3 |
| lap3 |
| lat |
| lat2 |
| layn |
| lbp |
| lbr |
| lcn2 |
| ldlr |
| lefty2 |
| lep |
| lepr |
| lgals1 |
| lgals3 |
| lgals4 |
| lgals7_lgals7b |
| lgals8 |
| lgals9 |
| lgmn |
| lhb |
| lhpp |
| lif |
| lifr |
| lilra2 |
| lilra5 |
| lilrb1 |
| lilrb2 |
| lilrb4 |

| Proteomic biomarkers under study |
|---|
| **Olink proteomic targets** |
| lilrb5 |
| lpcat2 |
| lpl |
| lpo |
| lrig1 |
| lrp1 |
| lrp11 |
| lrpap1 |
| lrrc25 |
| lrrn1 |
| lsm1 |
| lsp1 |
| lta |
| lta4h |
| ltbp2 |
| ltbp3 |
| ltbr |
| lto1 |
| lxn |
| ly6d |
| ly75 |
| ly9 |
| ly96 |
| lyar |
| lyn |
| lypd1 |
| lypd3 |
| lypd8 |
| mad1l1 |
| maea |
| maged1 |
| manf |
| mansc1 |
| map2k6 |
| map3k5 |
| map4k5 |
| mapk9 |
| mapt |
| marco |
| masp1 |
| matn2 |
| matn3 |

| Proteomic biomarkers under study |
|---|
| **Olink proteomic targets** |
| mavs |
| max |
| mb |
| mcam |
| mcfd2 |
| mdga1 |
| mdk |
| med18 |
| megf10 |
| megf9 |
| mep1b |
| mepe |
| mertk |
| mesd |
| met |
| metap1 |
| metap1d |
| metap2 |
| mfap3 |
| mfap5 |
| mfge8 |
| mgll |
| mgmt |
| mia |
| micb_mica |
| mif |
| milr1 |
| mitd1 |
| mln |
| mme |
| mmp1 |
| mmp10 |
| mmp12 |
| mmp13 |
| mmp3 |
| mmp7 |
| mmp8 |
| mmp9 |
| mnda |
| mog |
| mphosph8 |
| mpi |

| **Proteomic biomarkers under study** |
|---|
| **Olink proteomic targets** |
| mpig6b |
| mpo |
| mrpl46 |
| msln |
| msmb |
| msr1 |
| msra |
| mstn |
| mtpn |
| muc13 |
| muc16 |
| mvk |
| myo9b |
| myoc |
| mzb1 |
| mzt1 |
| naaa |
| nadk |
| nampt |
| nbl1 |
| nbn |
| ncam1 |
| ncam2 |
| ncan |
| ncf2 |
| nck2 |
| ncln |
| ncr1 |
| ncs1 |
| ndrg1 |
| ndufs6 |
| nectin2 |
| nectin4 |
| nefl |
| nell1 |
| nell2 |
| nfasc |
| nfatc1 |
| nfatc3 |
| nfkbie |
| ngf |
| nid1 |

| Proteomic biomarkers under study |
|---|
| **Olink proteomic targets** |
| nid2 |
| ninj1 |
| nme3 |
| nmnat1 |
| nomo1 |
| nos1 |
| nos3 |
| notch1 |
| notch3 |
| npdc1 |
| npm1 |
| nppb |
| nppc |
| nptn |
| nptx1 |
| nptxr |
| npy |
| nrcam |
| nrp1 |
| nrp2 |
| nrtn |
| nsfl1c |
| nt5c3a |
| nt5e |
| ntf3 |
| ntf4 |
| ntprobnp |
| ntrk2 |
| ntrk3 |
| nub1 |
| nucb2 |
| nudc |
| nudt2 |
| nudt5 |
| nxph1 |
| obp2b |
| odam |
| ogfr |
| ogn |
| olr1 |
| omd |
| omg |

| Proteomic biomarkers under study |
|---|
| **Olink proteomic targets** |
| optc |
| oscar |
| osm |
| osmr |
| oxt |
| p4hb |
| padi2 |
| padi4 |
| paep |
| pag1 |
| pak4 |
| pam |
| pamr1 |
| pappa |
| park7 |
| parp1 |
| pbld |
| pcdh1 |
| pcdh17 |
| pcolce |
| pcsk9 |
| pdcd1 |
| pdcd1lg2 |
| pdcd5 |
| pdcd6 |
| pdgfa |
| pdgfb |
| pdgfc |
| pdgfra |
| pdgfrb |
| pdlim7 |
| pdp1 |
| pear1 |
| pebp1 |
| pecam1 |
| pfdn2 |
| pfkfb2 |
| pgf |
| pglyrp1 |
| phospho1 |
| pi3 |
| pigr |

| Proteomic biomarkers under study |
|---|
| **Olink proteomic targets** |
| pik3ap1 |
| pik3ip1 |
| pilra |
| pilrb |
| pklr |
| pla2g10 |
| pla2g15 |
| pla2g1b |
| pla2g2a |
| pla2g4a |
| pla2g7 |
| plat |
| plau |
| plaur |
| plin1 |
| plin3 |
| plpbp |
| pltp |
| plxdc1 |
| plxna4 |
| plxnb2 |
| plxnb3 |
| pm20d1 |
| pmvk |
| pnliprp2 |
| pnpt1 |
| podxl |
| podxl2 |
| polr2f |
| pon2 |
| pon3 |
| ppcdc |
| ppib |
| ppm1a |
| ppme1 |
| ppp1r12a |
| ppp1r2 |
| ppp1r9b |
| ppp3r1 |
| ppy |
| pqbp1 |
| prcp |

| Proteomic biomarkers under study |
|---|
| **Olink proteomic targets** |
| prdx1 |
| prdx3 |
| prdx5 |
| prdx6 |
| preb |
| prelp |
| prkab1 |
| prkar1a |
| prkcq |
| prkra |
| prl |
| proc |
| prok1 |
| prss2 |
| prss27 |
| prss8 |
| prtfdc1 |
| prtg |
| prtn3 |
| psg1 |
| psip1 |
| psma1 |
| psmd9 |
| psme1 |
| psme2 |
| psmg3 |
| pspn |
| psrc1 |
| pten |
| ptgds |
| pth1r |
| ptk7 |
| ptn |
| ptpn1 |
| ptpn6 |
| ptprf |
| ptprm |
| ptprn2 |
| ptprs |
| pts |
| ptx3 |
| pvalb |

| **Proteomic biomarkers under study** |
|---|
| **Olink proteomic targets** |
| pvr |
| pxn |
| qdpr |
| qpct |
| rab37 |
| rab6a |
| rab6b |
| rabepk |
| rabgap1l |
| rad23b |
| rangap1 |
| rarres1 |
| rarres2 |
| rasa1 |
| rassf2 |
| rbks |
| rbp2 |
| rbp5 |
| rcor1 |
| reg1a |
| reg1b |
| reg3a |
| reg4 |
| relt |
| ren |
| ret |
| retn |
| rgma |
| rgmb |
| rgs8 |
| rhoc |
| rilp |
| rnase3 |
| rnaset2 |
| rnf41 |
| robo1 |
| robo2 |
| ror1 |
| rp2 |
| rrm2 |
| rrm2b |
| rspo1 |

| Proteomic biomarkers under study |
|---|
| **Olink proteomic targets** |
| rspo3 |
| rtbdn |
| rtn4r |
| ruvbl1 |
| rwdd1 |
| s100a11 |
| s100a12 |
| s100a16 |
| s100a4 |
| s100p |
| samd9l |
| scamp3 |
| scara5 |
| scarb1 |
| scarb2 |
| scarf1 |
| scarf2 |
| scg2 |
| scg3 |
| scgb1a1 |
| scgb3a2 |
| scgn |
| scly |
| scp2 |
| scrn1 |
| sdc1 |
| sdc4 |
| sele |
| selp |
| selplg |
| sema3f |
| sema4c |
| sema4d |
| sema7a |
| septin9 |
| serpina11 |
| serpina12 |
| serpina9 |
| serpinb1 |
| serpinb5 |
| serpinb6 |
| serpinb8 |

| Proteomic biomarkers under study |
|---|
| **Olink proteomic targets** |
| serpinb9 |
| serpine1 |
| sestd1 |
| setmar |
| sez6l |
| sez6l2 |
| sf3b4 |
| sfrp1 |
| sftpa1 |
| sftpa2 |
| sftpd |
| sh2b3 |
| sh2d1a |
| shmt1 |
| siae |
| siglec1 |
| siglec10 |
| siglec15 |
| siglec5 |
| siglec6 |
| siglec7 |
| siglec9 |
| sirpa |
| sirpb1 |
| sirt2 |
| sirt5 |
| sit1 |
| skap1 |
| skap2 |
| slamf1 |
| slamf6 |
| slamf7 |
| slamf8 |
| slc16a1 |
| slc27a4 |
| slc39a14 |
| slc39a5 |
| slit2 |
| slitrk2 |
| slitrk6 |
| smad1 |
| smad5 |

| Proteomic biomarkers under study |
|---|
| **Olink proteomic targets** |
| smarca2 |
| smoc1 |
| smoc2 |
| smpd1 |
| smpdl3a |
| snap23 |
| snap29 |
| sncg |
| snx9 |
| sod1 |
| sod2 |
| sorcs2 |
| sord |
| sort1 |
| sost |
| sparc |
| sparcl1 |
| spink1 |
| spink4 |
| spink5 |
| spink6 |
| spint1 |
| spint2 |
| spock1 |
| spon1 |
| spon2 |
| spp1 |
| spry2 |
| src |
| srp14 |
| srpk2 |
| ssb |
| ssc4d |
| ssc5d |
| st3gal1 |
| st6gal1 |
| stambp |
| stat5b |
| stc1 |
| stc2 |
| stip1 |
| stk11 |

| Proteomic biomarkers under study |
|---|
| **Olink proteomic targets** |
| stk24 |
| stk4 |
| stx16 |
| stx4 |
| stx6 |
| stx8 |
| stxbp3 |
| sugt1 |
| sult1a1 |
| sult2a1 |
| sumf2 |
| susd1 |
| susd2 |
| tacc3 |
| tacstd2 |
| tafa5 |
| tank |
| tarbp2 |
| tbc1d17 |
| tbc1d23 |
| tbc1d5 |
| tbcb |
| tbcc |
| tbl1x |
| tcl1a |
| tcl1b |
| tcn2 |
| tdgf1 |
| tdrkh |
| tek |
| tff1 |
| tff2 |
| tff3 |
| tfpi |
| tfpi2 |
| tfrc |
| tgfa |
| tgfb1 |
| tgfbi |
| tgfbr2 |
| tgfbr3 |
| tgm2 |

| Proteomic biomarkers under study |
|---|
| **Olink proteomic targets** |
| thbd |
| thbs2 |
| thbs4 |
| thop1 |
| thpo |
| thy1 |
| tia1 |
| tie1 |
| tigar |
| timd4 |
| timp1 |
| timp3 |
| timp4 |
| tinagl1 |
| tjap1 |
| tlr3 |
| tmprss15 |
| tmprss5 |
| tmsb10 |
| tnc |
| tnf |
| tnfaip8 |
| tnfrsf10a |
| tnfrsf10b |
| tnfrsf10c |
| tnfrsf11a |
| tnfrsf11b |
| tnfrsf12a |
| tnfrsf13b |
| tnfrsf13c |
| tnfrsf14 |
| tnfrsf19 |
| tnfrsf1a |
| tnfrsf1b |
| tnfrsf21 |
| tnfrsf4 |
| tnfrsf6b |
| tnfrsf8 |
| tnfrsf9 |
| tnfsf10 |
| tnfsf11 |
| tnfsf12 |

| Proteomic biomarkers under study |
|---|
| **Olink proteomic targets** |
| tnfsf13 |
| tnfsf13b |
| tnfsf14 |
| tnni3 |
| tnr |
| tnxb |
| tp53 |
| tp53inp1 |
| tpmt |
| tpp1 |
| tppp3 |
| tpsab1 |
| tpt1 |
| traf2 |
| trem2 |
| treml2 |
| triap1 |
| trim21 |
| trim5 |
| tshb |
| tslp |
| tspan1 |
| tst |
| txlna |
| txndc15 |
| txndc5 |
| txnrd1 |
| tymp |
| tyro3 |
| ubac1 |
| ulbp2 |
| umod |
| uso1 |
| usp8 |
| uxs1 |
| vamp5 |
| vash1 |
| vasn |
| vat1 |
| vcam1 |
| vcan |
| vegfa |

| Proteomic biomarkers under study |
|---|
| **Olink proteomic targets** |
| vegfc |
| vegfd |
| vim |
| vmo1 |
| vnn2 |
| vps37a |
| vps53 |
| vsig4 |
| vsir |
| vstm1 |
| vstm2l |
| vta1 |
| vtcn1 |
| vwa1 |
| vwc2 |
| vwf |
| wars |
| was |
| wasf1 |
| wasf3 |
| wfdc12 |
| wfdc2 |
| wfikkn1 |
| wfikkn2 |
| wif1 |
| wnt9a |
| wwp2 |
| xcl1 |
| xg |
| xpnpep2 |
| xrcc4 |
| yes1 |
| ythdf3 |
| zbtb16 |
| zbtb17 |

**Cox proportional hazards regression analysis of Alzheimer’s disease and endocrine, nutritional, metabolic, and digestive system disorders ICD-10 codes adjusted for APOE4/4 status, principal components 1-5, year of birth, Townsend deprivation index, and sex**

| UKB field corresponding to the ICD_10 code | ICD10_code | Definition of ICD10_code | Hazard Ratio | ci_min | ci_max | P_VAL | N_pairs | n | P_VAL_FDR_CORRECTED | rejected |
|---|---|---|---|---|---|---|---|---|---|---|
| p131654 | K66 | Other Disorders Of Peritoneum | 0.63 | 0.46 | 0.85 | 2.84E-03 | 41 | 4636 | 1.75E-02 | TRUE |
| p131650 | K64 | Haemorrhoids And Perianal Venous Thrombosis | 0.70 | 0.60 | 0.81 | 2.14E-06 | 186 | 20162 | 2.25E-05 | TRUE |
| p131598 | K29 | Gastritis And Duodenitis | 1.23 | 1.12 | 1.35 | 1.42E-05 | 523 | 27914 | 1.24E-04 | TRUE |
| p131582 | K20 | Oesophagitis | 1.28 | 1.10 | 1.49 | 1.55E-03 | 175 | 8660 | 1.08E-02 | TRUE |
| p130820 | E83 | Disorders Of Mineral Metabolism | 1.33 | 1.09 | 1.64 | 5.90E-03 | 94 | 4435 | 3.44E-02 | TRUE |
| p131630 | K52 | Other Non-Infective Gastro-Enteritis And Colitis | 1.36 | 1.20 | 1.55 | 3.04E-06 | 250 | 12984 | 2.90E-05 | TRUE |
| p131560 | K04 | Diseases Of Pulp And Periapical Tissues | 1.44 | 1.11 | 1.87 | 6.67E-03 | 57 | 3365 | 3.68E-02 | TRUE |
| p130828 | E87 | Other Disorders Of Fluid, Electrolyte And Acid-Base Balance | 1.47 | 1.31 | 1.66 | 9.30E-11 | 312 | 11313 | 1.48E-09 | TRUE |
| p130708 | E11 | Non-Insulin-Dependent Diabetes Mellitus | 1.55 | 1.40 | 1.71 | 3.46E-18 | 476 | 19048 | 1.82E-16 | TRUE |
| p131640 | K59 | Other Functional Intestinal Disorders | 1.55 | 1.39 | 1.72 | 4.25E-16 | 394 | 15596 | 1.49E-14 | TRUE |
| p130008 | A04 | Other Bacterial Intestinal Infections | 1.59 | 1.27 | 1.98 | 4.07E-05 | 81 | 3585 | 3.29E-04 | TRUE |
| p130714 | E14 | Unspecified Diabetes Mellitus | 1.60 | 1.39 | 1.83 | 1.94E-11 | 224 | 8733 | 4.08E-10 | TRUE |
| p130826 | E86 | Volume Depletion | 1.71 | 1.45 | 2.02 | 9.89E-11 | 153 | 4911 | 1.48E-09 | TRUE |
| p130770 | E53 | Deficiency Of Other B Group Vitamins | 1.80 | 1.41 | 2.29 | 1.73E-06 | 68 | 1939 | 2.02E-05 | TRUE |
| p130774 | E55 | Vitamin D Deficiency | 1.84 | 1.49 | 2.28 | 1.99E-08 | 87 | 2405 | 2.62E-07 | TRUE |
| p131658 | K70 | Alcoholic Liver Disease | 2.14 | 1.31 | 3.50 | 2.49E-03 | 16 | 955 | 1.64E-02 | TRUE |
| p130718 | E16 | Other Disorders Of Pancreatic Internal Secretion | 2.34 | 1.84 | 2.97 | 3.49E-12 | 69 | 1637 | 9.16E-11 | TRUE |
| p130706 | E10 | Insulin-Dependent Diabetes Mellitus | 3.03 | 2.40 | 3.82 | 9.49E-21 | 73 | 1633 | 9.96E-19 | TRUE |

UKB: UK Biobank
AD: Alzheimer's disease
ci_min: Confidence Interval minimum
ci_max: Confidence Interval maximum
P_VAL: p-value
N_pairs: Number of individuals identified with both ICD-10 code and neurodegenerative disease outcome
n: Number of Individulas Identified with ICD10_code
P_VAL_FDR_CORRECTED: p-value after False Discovery Rate corrected
Model: ICD10 + Apo_E4E4 + p22009_a1 + p22009_a2 + p22009_a3 + p22009_a4 + p22009_a5 + Year_of_birth + Townsend_deprivation_index + sex

**Cox proportional hazards regression analysis of Alzheimer's disease and endocrine, nutritional, metabolic, and digestive system disorders ICD-10 codes adjusted for polygenic risk Z-score excluding APOE, APOE4/4 status, principal components 1-5, year of birth, Townsend deprivation index, and sex**

| UKB field corresponding to the ICD_10 code | ICD10_code | Definition of ICD10_code | Hazard Ratio | ci_min | ci_max | P_VAL | N_pairs | n | P_VAL_FDR_CORRECTED | rejected |
|---|---|---|---|---|---|---|---|---|---|---|
| p131654 | K66 | Other Disorders Of Peritoneum | 0.63 | 0.46 | 0.86 | 3.53E-03 | 41 | 4636 | 2.06E-02 | TRUE |
| p131650 | K64 | Haemorrhoids And Perianal Venous Thrombosis | 0.70 | 0.61 | 0.82 | 3.12E-06 | 186 | 20162 | 3.28E-05 | TRUE |
| p131598 | K29 | Gastritis And Duodenitis | 1.23 | 1.12 | 1.35 | 1.48E-05 | 523 | 27914 | 1.29E-04 | TRUE |
| p131582 | K20 | Oesophagitis | 1.27 | 1.09 | 1.48 | 2.17E-03 | 175 | 8660 | 1.52E-02 | TRUE |
| p131630 | K52 | Other Non-Infective Gastro-Enteritis And Colitis | 1.35 | 1.19 | 1.54 | 4.30E-06 | 250 | 12984 | 4.11E-05 | TRUE |
| p130820 | E83 | Disorders Of Mineral Metabolism | 1.36 | 1.11 | 1.67 | 3.34E-03 | 94 | 4435 | 2.06E-02 | TRUE |
| p131560 | K04 | Diseases Of Pulp And Periapical Tissues | 1.45 | 1.12 | 1.88 | 5.51E-03 | 57 | 3365 | 3.04E-02 | TRUE |
| p130828 | E87 | Other Disorders Of Fluid, Electrolyte And Acid-Base Balance | 1.47 | 1.30 | 1.65 | 1.58E-10 | 312 | 11313 | 2.37E-09 | TRUE |
| p131640 | K59 | Other Functional Intestinal Disorders | 1.55 | 1.39 | 1.72 | 6.59E-16 | 394 | 15596 | 2.31E-14 | TRUE |
| p130708 | E11 | Non-Insulin-Dependent Diabetes Mellitus | 1.55 | 1.40 | 1.71 | 3.00E-18 | 476 | 19048 | 1.58E-16 | TRUE |
| p130714 | E14 | Unspecified Diabetes Mellitus | 1.60 | 1.39 | 1.83 | 1.84E-11 | 224 | 8733 | 3.87E-10 | TRUE |
| p130008 | A04 | Other Bacterial Intestinal Infections | 1.62 | 1.30 | 2.02 | 1.69E-05 | 81 | 3585 | 1.36E-04 | TRUE |
| p130826 | E86 | Volume Depletion | 1.73 | 1.47 | 2.03 | 5.36E-11 | 153 | 4911 | 9.38E-10 | TRUE |
| p130770 | E53 | Deficiency Of Other B Group Vitamins | 1.80 | 1.42 | 2.29 | 1.70E-06 | 68 | 1939 | 1.98E-05 | TRUE |
| p130774 | E55 | Vitamin D Deficiency | 1.84 | 1.48 | 2.27 | 2.50E-08 | 87 | 2405 | 3.29E-07 | TRUE |
| p131658 | K70 | Alcoholic Liver Disease | 2.08 | 1.27 | 3.41 | 3.48E-03 | 16 | 955 | 2.06E-02 | TRUE |
| p130718 | E16 | Other Disorders Of Pancreatic Internal Secretion | 2.34 | 1.84 | 2.97 | 3.14E-12 | 69 | 1637 | 8.24E-11 | TRUE |
| p130706 | E10 | Insulin-Dependent Diabetes Mellitus | 3.09 | 2.45 | 3.90 | 1.88E-21 | 73 | 1633 | 1.97E-19 | TRUE |

PRS: Polygenic risk score
UKB: UK Biobank
AD: Alzheimer's disease
ci_min: Confidence Interval minimum
ci_max: Confidence Interval maximum
P_VAL: p-value
N_pairs: Number of individuals identified with both ICD-10 code and neurodegenerative disease outcome
n: Number of Individulas Identified with ICD10_code
P_VAL_FDR_CORRECTED: p-value after False Discovery Rate corrected
Model: ICD10 + zSCORE_without_apoe + Apo_E4E4 + p22009_a1 + p22009_a2 + p22009_a3 + p22009_a4 + p22009_a5 + Year_of_birth + Townsend_deprivation_index + sex

**Cox proportional hazards regression analysis of Alzheimer's disease and endocrine, nutritional, metabolic, and digestive system disorders ICD-10 codes adjusted for polygenic risk Z-scores excluding APOE, principal components 1-5, year of birth, Townsend deprivation index, and sex**

| UKB field corresponding to the ICD_10 code | ICD10_code | Neurodegenera | Definition of ICD10_code | Hazard Ratio | ci_min | ci_max | P_VAL | N_pairs | n | P_VAL_FDR_CC | rejected |
|---|---|---|---|---|---|---|---|---|---|---|---|
| p131654 | K66 | AD | Other Disorders Of Peritoneum | 0.59 | 0.44 | 0.81 | 8.85E-04 | 41 | 4636 | 6.19E-03 | TRUE |
| p131650 | K64 | AD | Haemorrhoids And Perianal Venous Thrombosis | 0.69 | 0.59 | 0.80 | 6.54E-07 | 186 | 20162 | 7.63E-06 | TRUE |
| p131636 | K57 | AD | Diverticular Disease Of Intestine | 0.89 | 0.81 | 0.97 | 1.01E-02 | 573 | 35207 | 4.80E-02 | TRUE |
| p130814 | E78 | AD | Disorders Of Lipoprotein Metabolism And Other Lipidaemias | 1.18 | 1.09 | 1.27 | 1.63E-05 | 1090 | 53797 | 1.43E-04 | TRUE |
| p131598 | K29 | AD | Gastritis And Duodenitis | 1.20 | 1.09 | 1.32 | 1.61E-04 | 523 | 27914 | 1.20E-03 | TRUE |
| p131582 | K20 | AD | Oesophagitis | 1.26 | 1.08 | 1.47 | 2.85E-03 | 175 | 8660 | 1.66E-02 | TRUE |
| p130820 | E83 | AD | Disorders Of Mineral Metabolism | 1.32 | 1.07 | 1.62 | 8.20E-03 | 94 | 4435 | 4.10E-02 | TRUE |
| p131630 | K52 | AD | Other Non-Infective Gastro-Enteritis And Colitis | 1.36 | 1.20 | 1.55 | 2.50E-06 | 250 | 12984 | 2.52E-05 | TRUE |
| p131560 | K04 | AD | Diseases Of Pulp And Periapical Tissues | 1.44 | 1.11 | 1.87 | 6.61E-03 | 57 | 3365 | 3.65E-02 | TRUE |
| p130708 | E11 | AD | Non-Insulin-Dependent Diabetes Mellitus | 1.48 | 1.34 | 1.63 | 8.67E-15 | 476 | 19048 | 3.04E-13 | TRUE |
| p130828 | E87 | AD | Other Disorders Of Fluid, Electrolyte And Acid-Base Balance | 1.51 | 1.34 | 1.70 | 6.83E-12 | 312 | 11313 | 1.43E-10 | TRUE |
| p130714 | E14 | AD | Unspecified Diabetes Mellitus | 1.53 | 1.33 | 1.75 | 1.11E-09 | 224 | 8733 | 1.58E-08 | TRUE |
| p130008 | A04 | AD | Other Bacterial Intestinal Infections | 1.53 | 1.23 | 1.91 | 1.54E-04 | 81 | 3585 | 1.20E-03 | TRUE |
| p131640 | K59 | AD | Other Functional Intestinal Disorders | 1.55 | 1.39 | 1.72 | 6.02E-16 | 394 | 15596 | 3.16E-14 | TRUE |
| p130770 | E53 | AD | Deficiency Of Other B Group Vitamins | 1.78 | 1.40 | 2.27 | 2.64E-06 | 68 | 1939 | 2.52E-05 | TRUE |
| p130826 | E86 | AD | Volume Depletion | 1.78 | 1.52 | 2.10 | 3.27E-12 | 153 | 4911 | 8.59E-11 | TRUE |
| p130774 | E55 | AD | Vitamin D Deficiency | 1.94 | 1.57 | 2.40 | 1.20E-09 | 87 | 2405 | 1.58E-08 | TRUE |
| p130718 | E16 | AD | Other Disorders Of Pancreatic Internal Secretion | 2.23 | 1.75 | 2.83 | 5.29E-11 | 69 | 1637 | 9.26E-10 | TRUE |
| p130824 | E85 | AD | Amyloidosis | 2.42 | 1.26 | 4.66 | 8.08E-03 | 9 | 182 | 4.10E-02 | TRUE |
| p130706 | E10 | AD | Insulin-Dependent Diabetes Mellitus | 3.13 | 2.48 | 3.95 | 6.06E-22 | 73 | 1633 | 6.36E-20 | TRUE |

PRS: Polygenic risk score
UKB: UK Biobank
AD: Alzheimer's disease
ci_min: Confidence Interval minimum
ci_max: Confidence Interval maximum
P_VAL: p-value
N_pairs: Number of individuals identified with both ICD-10 code and neurodegenerative disease outcome
n: Number of Individulas Identified with ICD10_code
P_VAL_FDR_CORRECTED: p-value after False Discovery Rate corrected
Model: ICD10 + zSCORE_without_apoe + p22009_a1 + p22009_a2 + p22009_a3 + p22009_a4 + p22009_a5 + Year_of_birth + Townsend_deprivation_index + sex

**Cox proportional hazards regression analysis of Alzheimer's disease and endocrine, nutritional, metabolic, and digestive system disorders ICD-10 codes adjusted for polygenic risk Z-score, APOE4/4 status, principal components 1-5, year of birth, Townsend deprivation index, and sex**

| UKB field corresponding to the ICD_10 code | ICD10_code | Definition of ICD10_code | Hazard Ratio | ci_min | ci_max | P_VAL | N_pairs | n | P_VAL_FDR_CORRECTED | rejected |
|---|---|---|---|---|---|---|---|---|---|---|
| p131654 | K66 | Other Disorders Of Peritoneum | 0.63 | 0.46 | 0.86 | 3.39E-03 | 41 | 4636 | 2.02E-02 | TRUE |
| p131650 | K64 | Haemorrhoids And Perianal Venous Thrombosis | 0.70 | 0.61 | 0.82 | 3.13E-06 | 186 | 20162 | 3.28E-05 | TRUE |
| p131598 | K29 | Gastritis And Duodenitis | 1.23 | 1.12 | 1.35 | 1.47E-05 | 523 | 27914 | 1.29E-04 | TRUE |
| p131582 | K20 | Oesophagitis | 1.27 | 1.09 | 1.48 | 2.16E-03 | 175 | 8660 | 1.51E-02 | TRUE |
| p131630 | K52 | Other Non-Infective Gastro-Enteritis And Colitis | 1.35 | 1.19 | 1.54 | 4.28E-06 | 250 | 12984 | 4.08E-05 | TRUE |
| p130820 | E83 | Disorders Of Mineral Metabolism | 1.36 | 1.11 | 1.67 | 3.34E-03 | 94 | 4435 | 2.02E-02 | TRUE |
| p131560 | K04 | Diseases Of Pulp And Periapical Tissues | 1.45 | 1.12 | 1.88 | 5.50E-03 | 57 | 3365 | 3.04E-02 | TRUE |
| p130828 | E87 | Other Disorders Of Fluid, Electrolyte And Acid-Base Balance | 1.47 | 1.30 | 1.65 | 1.56E-10 | 312 | 11313 | 2.34E-09 | TRUE |
| p131640 | K59 | Other Functional Intestinal Disorders | 1.54 | 1.39 | 1.72 | 7.10E-16 | 394 | 15596 | 2.48E-14 | TRUE |
| p130708 | E11 | Non-Insulin-Dependent Diabetes Mellitus | 1.55 | 1.40 | 1.71 | 3.18E-18 | 476 | 19048 | 1.67E-16 | TRUE |
| p130714 | E14 | Unspecified Diabetes Mellitus | 1.60 | 1.39 | 1.83 | 1.82E-11 | 224 | 8733 | 3.83E-10 | TRUE |
| p130008 | A04 | Other Bacterial Intestinal Infections | 1.62 | 1.30 | 2.02 | 1.68E-05 | 81 | 3585 | 1.36E-04 | TRUE |
| p130826 | E86 | Volume Depletion | 1.73 | 1.47 | 2.03 | 5.33E-11 | 153 | 4911 | 9.32E-10 | TRUE |
| p130770 | E53 | Deficiency Of Other B Group Vitamins | 1.80 | 1.42 | 2.29 | 1.69E-06 | 68 | 1939 | 1.97E-05 | TRUE |
| p130774 | E55 | Vitamin D Deficiency | 1.84 | 1.48 | 2.27 | 2.49E-08 | 87 | 2405 | 3.27E-07 | TRUE |
| p131658 | K70 | Alcoholic Liver Disease | 2.08 | 1.27 | 3.41 | 3.47E-03 | 16 | 955 | 2.02E-02 | TRUE |
| p130718 | E16 | Other Disorders Of Pancreatic Internal Secretion | 2.34 | 1.84 | 2.97 | 3.13E-12 | 69 | 1637 | 8.21E-11 | TRUE |
| p130706 | E10 | Insulin-Dependent Diabetes Mellitus | 3.09 | 2.45 | 3.90 | 1.87E-21 | 73 | 1633 | 1.96E-19 | TRUE |

PRS: Polygenic risk score
UKB: UK Biobank
AD: Alzheimer's disease
ci_min: Confidence Interval minimum
ci_max: Confidence Interval maximum
P_VAL: p-value
N_pairs: Number of individuals identified with both ICD-10 code and neurodegenerative disease outcome
n: Number of Individulas Identified with ICD10_code
P_VAL_FDR_CORRECTED: p-value after False Discovery Rate corrected
Model: ICD10 + zSCORE + Apo_E4E4 + p22009_a1 + p22009_a2 + p22009_a3 + p22009_a4 + p22009_a5 + Year_of_birth + Townsend_deprivation_index + sex

**Cox proportional hazards regression analysis of Alzheimer's disease and endocrine, nutritional, metabolic, and digestive system disorders ICD-10 codes adjusted for polygenic risk Z-scores, principal components 1-5, year of birth, Townsend deprivation index, and sex**

| UKB field corresponding to the ICD_10 code | ICD10_code | Definition of ICD10_code | Hazard Ratio | ci_min | ci_max | P_VAL | N_pairs | n | P_VAL_FDR_CORRECTED | rejected |
|---|---|---|---|---|---|---|---|---|---|---|
| p131654 | K66 | Other Disorders Of Peritoneum | 0.63 | 0.46 | 0.86 | 3.26E-03 | 41 | 4636 | 2.01E-02 | TRUE |
| p131650 | K64 | Haemorrhoids And Perianal Venous Thrombosis | 0.70 | 0.61 | 0.82 | 3.18E-06 | 186 | 20162 | 3.34E-05 | TRUE |
| p131598 | K29 | Gastritis And Duodenitis | 1.23 | 1.12 | 1.35 | 1.55E-05 | 523 | 27914 | 1.30E-04 | TRUE |
| p131582 | K20 | Oesophagitis | 1.27 | 1.09 | 1.48 | 2.23E-03 | 175 | 8660 | 1.56E-02 | TRUE |
| p131630 | K52 | Other Non-Infective Gastro-Enteritis And Colitis | 1.35 | 1.19 | 1.54 | 4.28E-06 | 250 | 12984 | 4.08E-05 | TRUE |
| p130820 | E83 | Disorders Of Mineral Metabolism | 1.36 | 1.11 | 1.67 | 3.22E-03 | 94 | 4435 | 2.01E-02 | TRUE |
| p131560 | K04 | Diseases Of Pulp And Periapical Tissues | 1.45 | 1.12 | 1.89 | 5.37E-03 | 57 | 3365 | 2.97E-02 | TRUE |
| p130828 | E87 | Other Disorders Of Fluid, Electrolyte And Acid-Base Balance | 1.47 | 1.30 | 1.65 | 1.54E-10 | 312 | 11313 | 2.31E-09 | TRUE |
| p131640 | K59 | Other Functional Intestinal Disorders | 1.54 | 1.39 | 1.72 | 8.16E-16 | 394 | 15596 | 2.86E-14 | TRUE |
| p130708 | E11 | Non-Insulin-Dependent Diabetes Mellitus | 1.55 | 1.40 | 1.71 | 3.75E-18 | 476 | 19048 | 1.97E-16 | TRUE |
| p130714 | E14 | Unspecified Diabetes Mellitus | 1.60 | 1.39 | 1.83 | 2.02E-11 | 224 | 8733 | 4.23E-10 | TRUE |
| p130008 | A04 | Other Bacterial Intestinal Infections | 1.62 | 1.30 | 2.03 | 1.62E-05 | 81 | 3585 | 1.30E-04 | TRUE |
| p130826 | E86 | Volume Depletion | 1.73 | 1.47 | 2.04 | 4.48E-11 | 153 | 4911 | 7.85E-10 | TRUE |
| p130770 | E53 | Deficiency Of Other B Group Vitamins | 1.80 | 1.41 | 2.29 | 1.71E-06 | 68 | 1939 | 1.99E-05 | TRUE |
| p130774 | E55 | Vitamin D Deficiency | 1.84 | 1.48 | 2.27 | 2.41E-08 | 87 | 2405 | 3.16E-07 | TRUE |
| p131658 | K70 | Alcoholic Liver Disease | 2.07 | 1.27 | 3.39 | 3.75E-03 | 16 | 955 | 2.19E-02 | TRUE |
| p130718 | E16 | Other Disorders Of Pancreatic Internal Secretion | 2.34 | 1.84 | 2.97 | 3.21E-12 | 69 | 1637 | 8.43E-11 | TRUE |
| p130706 | E10 | Insulin-Dependent Diabetes Mellitus | 3.10 | 2.46 | 3.91 | 1.55E-21 | 73 | 1633 | 1.63E-19 | TRUE |

PRS: Polygenic risk score
UKB: UK Biobank
AD: Alzheimer's disease
ci_min: Confidence Interval minimum
ci_max: Confidence Interval maximum
P_VAL: p-value
N_pairs: Number of individuals identified with both ICD-10 code and neurodegenerative disease outcome
n: Number of Individulas Identified with ICD10_code
P_VAL_FDR_CORRECTED: p-value after False Discovery Rate corrected
Model: ICD10 + zSCORE + p22009_a1 + p22009_a2 + p22009_a3 + p22009_a4 + p22009_a5 + Year_of_birth + Townsend_deprivation_index + sex

**Cox proportional hazards regression analysis of Alzheimer's disease and endocrine, nutritional, metabolic, and digestive system disorders ICD-10 codes adjusted for principal components 1-5, year of birth, Townsend deprivation index, and sex**

| UKB field corresponding to the ICD_10 code | ICD10_code | Definition of ICD10_code | Hazard Ratio | ci_min | ci_max | P_VAL | N_pairs | n | P_VAL_FDR_CORRECTED | rejected |
|---|---|---|---|---|---|---|---|---|---|---|
| p131654 | K66 | Other Disorders Of Peritoneum | 0.59 | 0.44 | 0.81 | 8.75E-04 | 41 | 4636 | 5.74E-03 | TRUE |
| p131650 | K64 | Haemorrhoids And Perianal Venous Thrombosis | 0.69 | 0.59 | 0.80 | 6.73E-07 | 186 | 20162 | 7.85E-06 | TRUE |
| p130792 | E66 | Obesity | 0.84 | 0.74 | 0.96 | 9.27E-03 | 260 | 22619 | 4.43E-02 | TRUE |
| p131636 | K57 | Diverticular Disease Of Intestine | 0.89 | 0.81 | 0.97 | 1.08E-02 | 573 | 35207 | 4.94E-02 | TRUE |
| p130814 | E78 | Disorders Of Lipoprotein Metabolism And Other Lipidaemias | 1.18 | 1.09 | 1.27 | 1.68E-05 | 1090 | 53797 | 1.47E-04 | TRUE |
| p131598 | K29 | Gastritis And Duodenitis | 1.20 | 1.09 | 1.32 | 1.59E-04 | 523 | 27914 | 1.28E-03 | TRUE |
| p131582 | K20 | Oesophagitis | 1.28 | 1.10 | 1.49 | 1.68E-03 | 175 | 8660 | 1.03E-02 | TRUE |
| p130820 | E83 | Disorders Of Mineral Metabolism | 1.32 | 1.08 | 1.62 | 8.00E-03 | 94 | 4435 | 4.00E-02 | TRUE |
| p131630 | K52 | Other Non-Infective Gastro-Enteritis And Colitis | 1.37 | 1.20 | 1.56 | 1.91E-06 | 250 | 12984 | 2.00E-05 | TRUE |
| p131560 | K04 | Diseases Of Pulp And Periapical Tissues | 1.44 | 1.11 | 1.87 | 6.17E-03 | 57 | 3365 | 3.24E-02 | TRUE |
| p130708 | E11 | Non-Insulin-Dependent Diabetes Mellitus | 1.47 | 1.33 | 1.62 | 1.47E-14 | 476 | 19048 | 5.16E-13 | TRUE |
| p130828 | E87 | Other Disorders Of Fluid, Electrolyte And Acid-Base Balance | 1.51 | 1.34 | 1.70 | 6.72E-12 | 312 | 11313 | 1.41E-10 | TRUE |
| p130008 | A04 | Other Bacterial Intestinal Infections | 1.51 | 1.21 | 1.89 | 2.31E-04 | 81 | 3585 | 1.73E-03 | TRUE |
| p130714 | E14 | Unspecified Diabetes Mellitus | 1.53 | 1.33 | 1.75 | 1.19E-09 | 224 | 8733 | 1.56E-08 | TRUE |
| p131640 | K59 | Other Functional Intestinal Disorders | 1.55 | 1.40 | 1.73 | 3.41E-16 | 394 | 15596 | 1.79E-14 | TRUE |
| p130770 | E53 | Deficiency Of Other B Group Vitamins | 1.77 | 1.39 | 2.26 | 3.15E-06 | 68 | 1939 | 3.00E-05 | TRUE |
| p130826 | E86 | Volume Depletion | 1.78 | 1.52 | 2.10 | 3.47E-12 | 153 | 4911 | 9.12E-11 | TRUE |
| p130774 | E55 | Vitamin D Deficiency | 1.95 | 1.57 | 2.41 | 9.22E-10 | 87 | 2405 | 1.38E-08 | TRUE |
| p130718 | E16 | Other Disorders Of Pancreatic Internal Secretion | 2.23 | 1.76 | 2.84 | 4.79E-11 | 69 | 1637 | 8.39E-10 | TRUE |
| p130824 | E85 | Amyloidosis | 2.58 | 1.34 | 4.97 | 4.46E-03 | 9 | 182 | 2.46E-02 | TRUE |
| p130706 | E10 | Insulin-Dependent Diabetes Mellitus | 3.09 | 2.45 | 3.90 | 1.80E-21 | 73 | 1633 | 1.89E-19 | TRUE |

UKB: UK Biobank
AD: Alzheimer's disease
ci_min: Confidence Interval minimum
ci_max: Confidence Interval maximum
P_VAL: p-value
N_pairs: Number of individuals identified with both ICD-10 code and neurodegenerative disease outcome
n: Number of Individulas Identified with ICD10_code
P_VAL_FDR_CORRECTED: p-value after False Discovery Rate corrected
Model: ICD10 + p22009_a1 + p22009_a2 + p22009_a3 + p22009_a4 + p22009_a5 + Year_of_birth + Townsend_deprivation_index + sex

**Cox proportional hazards regression analysis of Parkinson's disease and endocrine, nutritional, metabolic, and digestive system disorders ICD-10 codes adjusted for principal components 1-5, year of birth, Townsend deprivation index, and sex**

| UKB field corresponding to the ICD_10 code | ICD10_code | Definition of ICD10_code | Hazard Ratio | ci_min | ci_max | P_VAL | N_pairs | n | P_VAL_FDR_CORRECTED | rejected |
|---|---|---|---|---|---|---|---|---|---|---|
| p131650 | K64 | haemorrhoids and perianal venous thrombosis | 0.59 | 0.50 | 0.71 | 3.25E-09 | 136 | 20006 | 6.36E-08 | TRUE |
| p131654 | K66 | other disorders of peritoneum | 0.62 | 0.45 | 0.87 | 5.59E-03 | 35 | 4612 | 4.21E-02 | TRUE |
| p131636 | K57 | diverticular disease of intestine | 0.69 | 0.62 | 0.77 | 7.26E-11 | 354 | 34801 | 2.37E-09 | TRUE |
| p131648 | K63 | other diseases of intestine | 0.69 | 0.59 | 0.80 | 1.38E-06 | 183 | 19139 | 2.25E-05 | TRUE |
| p130792 | E66 | obesity | 0.82 | 0.71 | 0.94 | 4.91E-03 | 211 | 22453 | 4.21E-02 | TRUE |
| p130708 | E11 | non-insulin-dependent diabetes mellitus | 1.21 | 1.08 | 1.36 | 1.40E-03 | 330 | 18815 | 1.37E-02 | TRUE |
| p131600 | K30 | dyspepsia | 1.34 | 1.13 | 1.59 | 9.03E-04 | 135 | 8811 | 9.83E-03 | TRUE |
| p131640 | K59 | other functional intestinal disorders | 1.56 | 1.38 | 1.76 | 5.23E-13 | 301 | 15431 | 5.12E-11 | TRUE |
| p130714 | E14 | unspecified diabetes mellitus | 1.61 | 1.39 | 1.86 | 2.06E-10 | 197 | 8658 | 5.04E-09 | TRUE |
| p130770 | E53 | deficiency of other b group vitamins | 1.73 | 1.30 | 2.31 | 1.62E-04 | 48 | 1907 | 1.98E-03 | TRUE |
| p130718 | E16 | other disorders of pancreatic internal secretion | 1.82 | 1.35 | 2.46 | 8.06E-05 | 44 | 1602 | 1.13E-03 | TRUE |
| p130706 | E10 | insulin-dependent diabetes mellitus | 2.64 | 2.01 | 3.46 | 2.96E-12 | 53 | 1608 | 1.45E-10 | TRUE |

UKB: UK Biobank
PD: Parkinson's disease
ci_min: Confidence Interval minimum
ci_max: Confidence Interval maximum
P_VAL: p-value
N_pairs: Number of individuals identified with both ICD-10 code and neurodegenerative disease outcome
n: Number of Individulas Identified with ICD10_code
P_VAL_FDR_CORRECTED: p-value after False Discovery Rate corrected
Model: ICD10 + p22009_a1 + p22009_a2 + p22009_a3 + p22009_a4 + p22009_a5 +Year_of_birth + Townsend_deprivation_index + sex

**Cox proportional hazards regression analysis of Parkinson's disease and endocrine, nutritional, metabolic, and digestive system disorders ICD-10 codes adjusted for polygenic risk Z-scores, principal components 1-5, Townsend deprivation index, and sex**

| UKB field corresponding to the ICD_10 code | ICD10_code | Definition of ICD10_code | Hazard Ratio | ci_min | ci_max | P_VAL | N_pairs | n | P_VAL_FDR_CORRECTED | rejected |
|---|---|---|---|---|---|---|---|---|---|---|
| p131650 | K64 | haemorrhoids and perianal venous thrombosis | 0.59 | 0.50 | 0.71 | 3.30E-09 | 136 | 20006 | 6.47E-08 | TRUE |
| p131654 | K66 | other disorders of peritoneum | 0.62 | 0.45 | 0.87 | 5.63E-03 | 35 | 4612 | 4.59E-02 | TRUE |
| p131648 | K63 | other diseases of intestine | 0.69 | 0.59 | 0.80 | 1.12E-06 | 183 | 19139 | 1.83E-05 | TRUE |
| p131636 | K57 | diverticular disease of intestine | 0.69 | 0.62 | 0.77 | 8.13E-11 | 354 | 34801 | 2.66E-09 | TRUE |
| p130792 | E66 | obesity | 0.82 | 0.71 | 0.95 | 6.35E-03 | 211 | 22453 | 4.78E-02 | TRUE |
| p130708 | E11 | non-insulin-dependent diabetes mellitus | 1.21 | 1.08 | 1.36 | 1.04E-03 | 330 | 18815 | 1.12E-02 | TRUE |
| p131600 | K30 | dyspepsia | 1.33 | 1.12 | 1.59 | 1.14E-03 | 135 | 8811 | 1.12E-02 | TRUE |
| p131640 | K59 | other functional intestinal disorders | 1.55 | 1.38 | 1.75 | 8.76E-13 | 301 | 15431 | 7.99E-11 | TRUE |
| p130714 | E14 | unspecified diabetes mellitus | 1.61 | 1.39 | 1.86 | 1.67E-10 | 197 | 8658 | 4.09E-09 | TRUE |
| p130770 | E53 | deficiency of other b group vitamins | 1.73 | 1.30 | 2.30 | 1.70E-04 | 48 | 1907 | 2.09E-03 | TRUE |
| p130718 | E16 | other disorders of pancreatic internal secretion | 1.81 | 1.34 | 2.44 | 9.86E-05 | 44 | 1602 | 1.38E-03 | TRUE |
| p130706 | E10 | insulin-dependent diabetes mellitus | 2.67 | 2.03 | 3.50 | 1.63E-12 | 53 | 1608 | 7.99E-11 | TRUE |

PRS: Polygenic risk score
UKB: UK Biobank
PD: Parkinson's disease
ci_min: Confidence Interval minimum
ci_max: Confidence Interval maximum
P_VAL: p-value
N_pairs: Number of individuals identified with both ICD-10 code and neurodegenerative disease outcome
n: Number of Individulas Identified with ICD10_code
P_VAL_FDR_CORRECTED: p-value after False Discovery Rate corrected
Model: ICD10 + zSCORE + p22009_a1 + p22009_a2 + p22009_a3 + p22009_a4 + p22009_a5 + Year_of_birth + Townsend_deprivation_index + sex

**Cox proportional hazards regression analysis of Parkinson's disease and endocrine, nutritional, metabolic, and digestive system disorders ICD-10 codes adjusted for GBA1 (G_1_155162560 + T_1_155235843) status, principal components 1-5, year of birth, Townsend deprivation index, and sex**

| UKB field corresponding to the ICD_10 code | ICD10_code | Definition of ICD10_code | Hazard Ratio | ci_min | ci_max | P_VAL | N_pairs | n | P_VAL_FDR_CORRECTED | rejected |
|---|---|---|---|---|---|---|---|---|---|---|
| p131650 | K64 | haemorrhoids and perianal venous thrombosis | 0.59 | 0.50 | 0.71 | 3.24E-09 | 136 | 20006 | 6.36E-08 | TRUE |
| p131654 | K66 | other disorders of peritoneum | 0.62 | 0.45 | 0.87 | 5.62E-03 | 35 | 4612 | 4.33E-02 | TRUE |
| p131636 | K57 | diverticular disease of intestine | 0.69 | 0.62 | 0.77 | 7.52E-11 | 354 | 34801 | 2.46E-09 | TRUE |
| p131648 | K63 | other diseases of intestine | 0.69 | 0.59 | 0.80 | 1.38E-06 | 183 | 19139 | 2.25E-05 | TRUE |
| p130792 | E66 | obesity | 0.82 | 0.71 | 0.94 | 5.02E-03 | 211 | 22453 | 4.33E-02 | TRUE |
| p130708 | E11 | non-insulin-dependent diabetes mellitus | 1.21 | 1.07 | 1.36 | 1.58E-03 | 330 | 18815 | 1.55E-02 | TRUE |
| p131600 | K30 | dyspepsia | 1.34 | 1.12 | 1.59 | 1.04E-03 | 135 | 8811 | 1.13E-02 | TRUE |
| p131640 | K59 | other functional intestinal disorders | 1.56 | 1.38 | 1.76 | 4.76E-13 | 301 | 15431 | 4.67E-11 | TRUE |
| p130714 | E14 | unspecified diabetes mellitus | 1.60 | 1.38 | 1.85 | 2.99E-10 | 197 | 8658 | 7.32E-09 | TRUE |
| p130770 | E53 | deficiency of other b group vitamins | 1.73 | 1.30 | 2.30 | 1.70E-04 | 48 | 1907 | 2.08E-03 | TRUE |
| p130718 | E16 | other disorders of pancreatic internal secretion | 1.81 | 1.34 | 2.44 | 9.93E-05 | 44 | 1602 | 1.39E-03 | TRUE |
| p130706 | E10 | insulin-dependent diabetes mellitus | 2.63 | 2.00 | 3.45 | 3.44E-12 | 53 | 1608 | 1.68E-10 | TRUE |

UKB: UK Biobank
PD: Parkinson's disease
ci_min: Confidence Interval minimum
ci_max: Confidence Interval maximum
P_VAL: p-value
N_pairs: Number of individuals identified with both ICD-10 code and neurodegenerative disease outcome
n: Number of Individulas Identified with ICD10_code
P_VAL_FDR_CORRECTED: p-value after False Discovery Rate corrected
Model: ICD10 +G_1_155162560 + T_1_155235843 + p22009_a1 + p22009_a2 + p22009_a3 + p22009_a4 + p22009_a5 + Year_of_birth + Townsend_deprivation_index + sex

**Cox proportional hazards regression analysis of Parkinson's disease and endocrine, nutritional, metabolic, and digestive system disorders ICD-10 codes adjusted for polygenic risk Z-scores, GBA1 (G_1_155162560 + T_1_155235843) status, principal components 1-5, year of birth, Townsend deprivation index, and sex**

| UKB field corresponding to the ICD_10 code | ICD10_code | Definition of ICD10_code | Hazard Ratio | ci_min | ci_max | P_VAL | N_pairs | n | P_VAL_FDR_CORRECTED | rejected |
|---|---|---|---|---|---|---|---|---|---|---|
| p131650 | K64 | haemorrhoids and perianal venous thrombosis | 0.59 | 0.50 | 0.71 | 3.34E-09 | 136 | 20006 | 6.55E-08 | TRUE |
| p131654 | K66 | other disorders of peritoneum | 0.62 | 0.45 | 0.87 | 5.63E-03 | 35 | 4612 | 4.60E-02 | TRUE |
| p131648 | K63 | other diseases of intestine | 0.69 | 0.59 | 0.80 | 1.12E-06 | 183 | 19139 | 1.83E-05 | TRUE |
| p131636 | K57 | diverticular disease of intestine | 0.69 | 0.62 | 0.77 | 8.19E-11 | 354 | 34801 | 2.68E-09 | TRUE |
| p130792 | E66 | obesity | 0.82 | 0.71 | 0.95 | 6.37E-03 | 211 | 22453 | 4.80E-02 | TRUE |
| p130708 | E11 | non-insulin-dependent diabetes mellitus | 1.21 | 1.08 | 1.36 | 1.10E-03 | 330 | 18815 | 1.17E-02 | TRUE |
| p131600 | K30 | dyspepsia | 1.33 | 1.12 | 1.58 | 1.19E-03 | 135 | 8811 | 1.17E-02 | TRUE |
| p131640 | K59 | other functional intestinal disorders | 1.55 | 1.38 | 1.75 | 8.57E-13 | 301 | 15431 | 8.40E-11 | TRUE |
| p130714 | E14 | unspecified diabetes mellitus | 1.61 | 1.39 | 1.86 | 1.91E-10 | 197 | 8658 | 4.68E-09 | TRUE |
| p130770 | E53 | deficiency of other b group vitamins | 1.73 | 1.30 | 2.30 | 1.71E-04 | 48 | 1907 | 2.10E-03 | TRUE |
| p130718 | E16 | other disorders of pancreatic internal secretion | 1.81 | 1.34 | 2.43 | 1.05E-04 | 44 | 1602 | 1.47E-03 | TRUE |
| p130706 | E10 | insulin-dependent diabetes mellitus | 2.66 | 2.03 | 3.50 | 1.78E-12 | 53 | 1608 | 8.70E-11 | TRUE |

PRS: Polygenic risk score
UKB: UK Biobank
PD: Parkinson's disease
ci_min: Confidence Interval minimum
ci_max: Confidence Interval maximum
P_VAL: p-value
N_pairs: Number of individuals identified with both ICD-10 code and neurodegenerative disease outcome
n: Number of Individulas Identified with ICD10_code
P_VAL_FDR_CORRECTED: p-value after False Discovery Rate corrected
Model: ICD10 + zSCORE + G_1_155162560 + T_1_155235843 + p22009_a1 + p22009_a2 + p22009_a3 + p22009_a4 + p22009_a5 + Year_of_birth + Townsend_deprivation_index + sex

**Cox proportional hazards regression analysis of Parkinson's disease and endocrine, nutritional, metabolic, and digestive system disorders ICD-10 codes adjusted for LRRK2 (C_12_40220632 + G_12_40340400) status, principal components 1-5, year of birth, Townsend deprivation index, and sex**

| UKB field corresponding to the ICD_10 code | ICD10_code | Definition of ICD10_code | Hazard Ratio | ci_min | ci_max | P_VAL | N_pairs | n | P_VAL_FDR_CORRECTED | rejected |
|---|---|---|---|---|---|---|---|---|---|---|
| p131650 | K64 | haemorrhoids and perianal venous thrombosis | 0.59 | 0.50 | 0.70 | 2.83E-09 | 136 | 20006 | 5.56E-08 | TRUE |
| p131654 | K66 | other disorders of peritoneum | 0.62 | 0.45 | 0.87 | 5.54E-03 | 35 | 4612 | 4.17E-02 | TRUE |
| p131636 | K57 | diverticular disease of intestine | 0.69 | 0.62 | 0.77 | 7.83E-11 | 354 | 34801 | 2.56E-09 | TRUE |
| p131648 | K63 | other diseases of intestine | 0.69 | 0.59 | 0.80 | 1.40E-06 | 183 | 19139 | 2.29E-05 | TRUE |
| p130792 | E66 | obesity | 0.82 | 0.71 | 0.94 | 4.46E-03 | 211 | 22453 | 3.97E-02 | TRUE |
| p130708 | E11 | non-insulin-dependent diabetes mellitus | 1.21 | 1.07 | 1.36 | 1.49E-03 | 330 | 18815 | 1.46E-02 | TRUE |
| p131600 | K30 | dyspepsia | 1.34 | 1.13 | 1.59 | 9.28E-04 | 135 | 8811 | 1.01E-02 | TRUE |
| p131640 | K59 | other functional intestinal disorders | 1.56 | 1.38 | 1.76 | 5.89E-13 | 301 | 15431 | 5.77E-11 | TRUE |
| p130714 | E14 | unspecified diabetes mellitus | 1.60 | 1.38 | 1.85 | 2.41E-10 | 197 | 8658 | 5.90E-09 | TRUE |
| p130770 | E53 | deficiency of other b group vitamins | 1.73 | 1.30 | 2.31 | 1.63E-04 | 48 | 1907 | 1.99E-03 | TRUE |
| p130718 | E16 | other disorders of pancreatic internal secretion | 1.83 | 1.36 | 2.46 | 7.69E-05 | 44 | 1602 | 1.08E-03 | TRUE |
| p130706 | E10 | insulin-dependent diabetes mellitus | 2.63 | 2.00 | 3.45 | 3.48E-12 | 53 | 1608 | 1.70E-10 | TRUE |

UKB: UK Biobank
PD: Parkinson's disease
ci_min: Confidence Interval minimum
ci_max: Confidence Interval maximum
P_VAL: p-value
N_pairs: Number of individuals identified with both ICD-10 code and neurodegenerative disease outcome
n: Number of Individulas Identified with ICD10_code
P_VAL_FDR_CORRECTED: p-value after False Discovery Rate corrected
Model: ICD10 + C_12_40220632 + G_12_40340400 + p22009_a1 + p22009_a2 + p22009_a3 + p22009_a4 + p22009_a5 + Year_of_birth + Townsend_deprivation_index + sex

**Cox proportional hazards regression analysis of Parkinson's disease and endocrine, nutritional, metabolic, and digestive system disorders ICD-10 codes adjusted for polygenic risk Z-scores, LRRK2 (C_12_40220632 + G_12_40340400) status, principal components 1-5, year of birth, Townsend deprivation index, and sex**

| UKB field corresponding to the ICD_10 code | ICD10_code | Definition of ICD10_code | Hazard Ratio | ci_min | ci_max | P_VAL | N_pairs | n | P_VAL_FDR_CORRECTED | rejected |
|---|---|---|---|---|---|---|---|---|---|---|
| p131650 | K64 | haemorrhoids and perianal venous thrombosis | 0.59 | 0.50 | 0.70 | 3.02E-09 | 136 | 20006 | 5.91E-08 | TRUE |
| p131654 | K66 | other disorders of peritoneum | 0.62 | 0.45 | 0.87 | 5.56E-03 | 35 | 4612 | 4.53E-02 | TRUE |
| p131648 | K63 | other diseases of intestine | 0.69 | 0.59 | 0.80 | 1.08E-06 | 183 | 19139 | 1.77E-05 | TRUE |
| p131636 | K57 | diverticular disease of intestine | 0.69 | 0.62 | 0.77 | 8.22E-11 | 354 | 34801 | 2.69E-09 | TRUE |
| p130792 | E66 | obesity | 0.82 | 0.71 | 0.95 | 6.01E-03 | 211 | 22453 | 4.53E-02 | TRUE |
| p130708 | E11 | non-insulin-dependent diabetes mellitus | 1.21 | 1.08 | 1.36 | 1.07E-03 | 330 | 18815 | 1.13E-02 | TRUE |
| p131600 | K30 | dyspepsia | 1.33 | 1.12 | 1.58 | 1.16E-03 | 135 | 8811 | 1.13E-02 | TRUE |
| p131640 | K59 | other functional intestinal disorders | 1.55 | 1.37 | 1.75 | 9.20E-13 | 301 | 15431 | 8.97E-11 | TRUE |
| p130714 | E14 | unspecified diabetes mellitus | 1.61 | 1.39 | 1.86 | 1.80E-10 | 197 | 8658 | 4.40E-09 | TRUE |
| p130770 | E53 | deficiency of other b group vitamins | 1.73 | 1.30 | 2.31 | 1.66E-04 | 48 | 1907 | 2.03E-03 | TRUE |
| p130718 | E16 | other disorders of pancreatic internal secretion | 1.81 | 1.34 | 2.44 | 9.66E-05 | 44 | 1602 | 1.35E-03 | TRUE |
| p130706 | E10 | insulin-dependent diabetes mellitus | 2.66 | 2.03 | 3.50 | 1.83E-12 | 53 | 1608 | 8.97E-11 | TRUE |

PRS: Polygenic risk score
UKB: UK Biobank
PD: Parkinson's disease
ci_min: Confidence Interval minimum
ci_max: Confidence Interval maximum
P_VAL: p-value
N_pairs: Number of individuals identified with both ICD-10 code and neurodegenerative disease outcome
n: Number of Individulas Identified with ICD10_code
P_VAL_FDR_CORRECTED: p-value after False Discovery Rate corrected
Model: ICD10 + zSCORE  + C_12_40220632 + G_12_40340400 + p22009_a1 + p22009_a2 + p22009_a3 + p22009_a4 + p22009_a5 + Year_of_birth + Townsend_deprivation_index + sex
